# First-time child protection contacts from the prenatal period to age 15 among Aboriginal children born 2006- 2020: a whole-population cohort study in New South Wales, Australia

**DOI:** 10.64898/2026.03.24.26349231

**Authors:** Mark Hanly, BJ Newton, Tasnia Ahmed, Tiah Payne, Madeleine Powell, Kyllie Cripps, Ilan Katz, Rhiannon Pilkington, John Lynch, Paul Gray, Kathleen Falster, the Bring Them Home, Keep Them Home investigators

## Abstract

**Background:** First Nations children are over-represented in child protection systems in Australia and other colonised countries. Here, we apply a prevention and equity lens to the use of child protection data, to inform early opportunities to support Aboriginal children and families at risk of escalating child protection contact, from pregnancy to adolescence.

**Methods:** We followed 15 whole-population cohorts (born 2006-2020) of Aboriginal (n=119,716) and non-Aboriginal (n=1,456,698) children in New South Wales (NSW), Australia, to December 2021, using birth and child protection datasets linked for the NSW Child E-Cohort. In each Aboriginal and non-Aboriginal cohort born 2006 to 2020, we calculated the cumulative incidence (risk) of first-time child protection contacts from the prenatal period up to age 15 years: reports to child protection, screened in reports, investigations, child protection- defined substantiations (of actual harm or risk of significant harm assessed in the investigation), and OOHC placements. Risk differences and relative risks were also calculated.

**Findings:** By birth, 10-15% of Aboriginal children born 2006-2020 had a first report to child protection, with 48-54% by age 5y (2006-2016 births), and 74% by age 15y (2006 births), with similar risks of screened-in reports (e.g. 68% by age 15y). The risk of first-time substantiation was 1- 5% of Aboriginal children by birth, 17-20% by 5y, and 32% by 15y, with higher risks in more contemporary cohorts. By age 1y, 3-4% of Aboriginal children born 2006-2020 had a first OOHC placement, with 7-9% by 5y, and 14% by 15y. The risk differences between Aboriginal and non-Aboriginal children were 23 and 3 percentage points for reports and OOHC by age 1y (2020 births), respectively, increasing as children age.

**Interpretation:** Despite extensive inquiries, calls for prevention and Closing the Gap targets, our study shows the lifetime risk of child protection involvement for Aboriginal families has not improved and inequities persist. These findings support the call for Aboriginal-led approaches and greater investment in early supports for First Nations children and families.

**Research in Context:** *Evidence before this study:* We searched PubMed and Medline for studies on the lifetime risk of child protection contacts among First Nations child populations, published January 2005 to May 2025. Thirteen studies reported various child protection contacts, from the perinatal period through childhood, among birth or synthetic cohorts of First Nations children, born between 1990 and 2018, created from population data sources in jurisdictions in Australia (n=5), the United States(US) (n=6), and Aotearoa/New Zealand (NZ) (n=2) (Table E1). The most recently published study included First Nations children born 2000 to 2013 in Western Australia, which quantified the risk of reports, investigations, substantiations and removals into OOHC, from age 1 to 16 years. By age 1, 12% were reported and 3% were removed into OOHC. By age 16, 52% were reported, and 14% were removed into OOHC. Prior studies of birth or synthetic cohorts of First Nations children born 1990-2018, in the USA, NZ, and South Australia showed similar results. By age 5 years, 16% to 54% for reports, 20% for investigations, 7% to 11% for substantiations and 8% for removals into OOHC. Among the five studies with cohorts followed to 18 years, 42% were reported, 28% to 50% were investigated, 9% to 27% were substantiated, 7% to 16% were removed into OOHC and 0.8% to 3.8% had termination of parental rights.

*Added value of this study:* This is the largest and most contemporary study to quantify the lifetime risk of child protection contact among whole-populations of First Nations children internationally. Among 15 consecutive whole-population cohorts of First Nations children in New South Wales (NSW), Australia, born 2006 to 2020, we reported—for the first time—the full spectrum of child protection contacts, from the prenatal period. By birth, 16% were reported to child protection, 14% were investigated and 5% were substantiated in the most contemporary cohort born 2020. By age 1 year, 2.8% were removed into OOHC. In the oldest cohort born 2006, 74% were reported and 14.4% removed into OOHC by age 15 years. We also reveal the magnitude of the inequity in child protection contacts between First Nations and non-Indigenous children across the lifecourse. For example, among 2006 births, the risk of first-time reports to child protection for Aboriginal and non-Aboriginal children, respectively, was 10.5% versus 1.5% by birth (risk difference (RD), 9 percentage points; risk ratio (RR), 7.0), 53% vs 16% by age five (RD, 38pp; RR, 3.4) and 74% vs 33% by age 15 (RD, 41pp; RR 2.2).

*Implications of all the available evidence:* This study unequivocally shows that the lifetime risk of child protection involvement in the lives of First Nations families has not reduced in more contemporary whole-population cohorts and that inequities persist. This is consistent with evidence from prior studies internationally. It is critical that First Nations-led responses and investment in early family supports must be at the centre of system reform to realise the long-called-for shift toward prevention and to re-dress the pervasive inequities experienced by First Nations children and families in colonised countries such as Australia.

## Introduction

Aboriginal and Torres Strait Islander people are the First People of Australia, with over 60,000 years of history. In 2021, Aboriginal and Torres Strait Islander people were 4% of the total Australian population, and 6% of the population aged 0–18 years. While the young First Nations population is growing, there remains a long-standing national crisis of over- representation in Australian child protection systems—in 2023, Aboriginal and Torres Strait Islander children comprised 43% of the out-of-home care (OOHC) population nationally.^1^ For decades, routine reporting has illustrated the over-representation and pervasive inequities in point-in-time child protection system statistics for First Nations children and families in Australia, as well as other colonised countries such as Aotearoa/New Zealand (NZ), Canada, and the United States (US).^2–4^

While histories of colonisation and dispossession vary internationally, the persistent over- representation of First Nations peoples in child protection systems reflects the enduring impact of colonial structures and policies. These systems have resulted in entrenched disadvantage and state-sanctioned violence toward First Nations children, families, and communities—consequences felt today and across generations.^5–10^ In Australia, the forced removal of thousands of First Nations children from their families and communities throughout much of the twentieth century are known as the ‘Stolen Generations’.^11^ There has been formal acknowledgement of this history, accompanied by ‘Closing the Gap’ policies and targets specifically focused on OOHC.^12^ However, recent Aboriginal-led reviews have highlighted that the extent of contemporary child protection removals were similar to the Stolen Generations.^5–10^ To address this crisis, multiple independent audits and reviews (cf Figure E2) have called for investment in prevention to support the unmet needs of Aboriginal children and families that contributes to child protection system contact.^13^

For this study, we apply a prevention lens to the use of administrative child protection system data, quantifying the risk of child protection system contacts from gestation into adolescence, in whole-populations of Aboriginal and non-Aboriginal children. The goal is to generate evidence to inform the appropriate resourcing of culturally appropriate, supportive prevention responses to families at the earliest time in a child’s life. Previously, several studies of birth cohorts from 1986-2013, in the USA, NZ, and Australia, have shown a high risk of specific child protection contacts at different time points in children’s lives.^14–26^ For example, 7-16% of First Nations children had ≥1 OOHC placement by 18 years. The most contemporary study of First Nations children, in Western Australia, was the first to quantify the risk of multiple child protection contacts from age 1 to 16 years. This study showed half of all First Nations children were reported, and 1 in 7 in OOHC, by age 16, among children born 2000 to 2013.

This study is a step towards realising the ambition of Indigenous Data Governance and Indigenous Data Sovereignty^1^ in generating evidence to inform prevention-oriented responses to Aboriginal families at the earliest possible opportunity in children’s lives. We quantified the risk of all levels of first-time child protection system contacts—from reports to child protection to OOHC—from gestation to age 15 years, among Aboriginal children in NSW, born from 2006 to 2020. Aligned with the national Closing the Gap policy and targets, we also quantified the size of the inequities between First Nations and non-Indigenous children.

## Methods

### Study design

Whole-population cohort study using data linked for the NSW Child E-Cohort Project. This study was undertaken as part of the Bring Them Home, Keep Them Home project,^27,28^ an Aboriginal-led mixed methods study that aims to understand the child protection experiences, outcomes and pathways to restoration^2^ for Aboriginal children in OOHC in NSW. Four of the 11 authors of this research are Aboriginal (BN, TP, KC, PG). From the project’s conception, the research was undertaken in partnership with Aboriginal organisations and stakeholders, including AbSec (the NSW Child, Family and Community Peak Aboriginal Corporation), and three Aboriginal Community Controlled Organisations (ACCOs) in the Illawarra Shoalhaven region. We also engaged with the Aboriginal Health and Medical Research Council (AH&MRC) of NSW and the NSW Department of Communities and Justice (DCJ) State Aboriginal Reference Group.

### Data sources and data linkage

We used NSW DCJ child protection data linked to birth records from the NSW Perinatal Data Collection (PDC), the NSW Registry of Births, Deaths, and Marriages (RBDM), and the NSW Admitted Patient Data Collection (APDC). Probabilistic record linkage was undertaken by the NSW Centre for Health Record Linkage (false positive and false negative rates <0.5%).^29^ We also used NSW population estimates for the Aboriginal population^30^ and total population^31^ by birth year and year of age, compiled by the Australian Bureau of Statistics (ABS).

### Study population

The primary study population included Aboriginal children in NSW for any duration from the prenatal period through childhood. First, we assembled data for an open cohort of all NSW children born from 1 January 2006 to 31 December 2020, including children with NSW birth records and/or child protection records at any time between their estimated date of conception until their birthday in 2021 (Figure E1). Children were included in the Aboriginal child population if the child or any parent was recorded as Aboriginal and/or Torres Strait Islander in any of the birth datasets at birth or in any child protection datasets from the prenatal period until the end of follow-up in 2021 (N=119,716). All other children were included in the non-Aboriginal child population (N=1,456,698). Among Aboriginal children, 103,089 (86.1%) had a NSW birth record and 16,627 (13.9%) were identified solely from the child protection datasets. Among non-Aboriginal children, 1,369,381 (94.0%) had a NSW birth record and 87,317 (6.0%) were identified solely from the child protection data. We divided the Aboriginal and non-Aboriginal study populations into birth-year cohorts, which included children born in NSW and/or children with records in NSW child protection data.

### Outcomes

We examined first-time child protection contacts, including: (i) reports to child protection, (ii) reports screened-in by child protection as “at risk of significant harm”, (iii) investigations, (iv) child protection-defined substantiations (of actual harm or risk of significant harm assessed in the investigation), and (v) removal into OOHC. Note that the threshold for screening in reports was raised from ‘risk of harm’ to ‘risk of significant’ harm in 2010, as part of the Keep Them Safe initiatives introduced after the Wood Royal Commission^13^ (Figure E2). We calculated the year of age at the first occurrence of each outcome using the date recorded in the child protection data, and the date of birth from the birth, or child protection datasets (if no NSW birth record).

### Statistical analysis

For each birth cohort, we calculated the cumulative number of Aboriginal children who experienced each outcome for the first time by each birthday, from estimated conception until the child’s birthday in 2021. It was possible to follow the youngest birth cohort (2020) until their first birthday and the oldest birth cohort (2006) until their 15^th^ birthday. To estimate the cumulative incidence (henceforth ‘risk’) in each birth cohort, the cumulative number of children with each outcome by each birthday was divided by the estimated size of the Aboriginal child population for each birth cohort. The population denominator was derived by counting all unique children recorded in ≥1 of the three birth datasets, or with ≥1 child protection records from gestation to 31 December 2021. The analysis was repeated for the non-Aboriginal child population, with risk differences and risk ratios calculated for select birth cohorts as measures of inequity. Analysis used the *tidyverse* suite of packages^32^ in R version 4.5.2.^33^

Figures E4 and E5 and Tables E5a-e and E6a-e present additional analyses to examine the range of risk estimates when using alternative combinations of linked datasets to define the Aboriginal child population size or the ABS estimated resident population denominator routinely used in government reporting.

### Ethical approval

This study was approved by the NSW Population and Health Services Research Ethics Committee (2020/ETH01265), the University of NSW HREC (2020/ETH01265), and the Aboriginal Health and Medical Research Council (AH&MRC) of NSW Ethics Committee (1688/20). The CHeReL operates under strict data security protocols and implements high level physical security measures. Their security protocols are in accordance with the Australian Government Protective Security Policy Framework, the Population Health Research Network Information Governance Framework, and the NHMRC Code for Responsible Conduct of Research.

## Results

### Aboriginal child population birth-year cohorts

In total, 119,716 NSW Aboriginal children were included in the study, with birth cohort sizes ranging from 7,300 in 2006 to 8,604 in 2020. The estimated population denominator based on the linked birth data sources exceeded the ABS estimated resident population for Aboriginal children by 1,000-2,100 individuals across the 2006-2020 birth cohorts (Figure E3, Table E2a).

### First-time reports to child protection in the Aboriginal child population

The risk of ≥1 report to child protection during gestation was in the range of 10-17% (769- 1360 children) for the 2006 to 2020 birth-year cohorts, increasing to 48-54% by age five across the 2006-2016 birth-year cohorts (Figure 1; Table E3a). By age 15 years, 74% of children (5,410 children) had ≥1 report to child protection among the 2006 birth-year cohort. Following the introduction of the Keep Them Safe Initiatives in 2010 (including changes to mandatory reporting guidelines, introduction of Child Wellbeing Units, and an increase in the risk screening thresholds^34^), the risk of reports to child protection during gestation among children born 2011-2012 decreased to <13% before trending upwards again, reaching 15.8% among children born in 2020.

**Figure 1.**
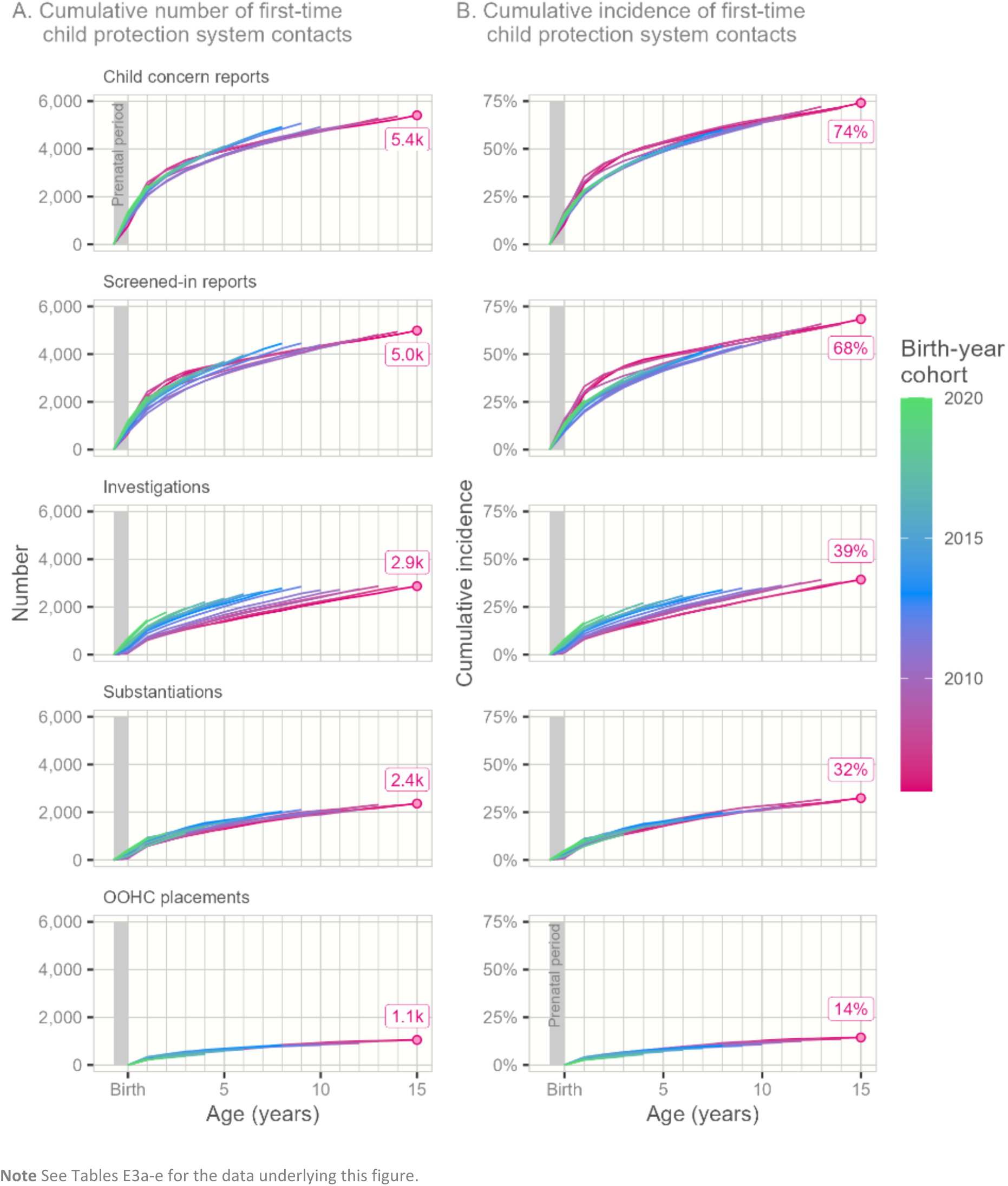
The cumulative number (A) and cumulative incidence (B) of first-time child protection contacts by age for NSW Aboriginal children born 2006–2020, followed from the prenatal period up to age 15 years

### First-time child protection screened-in reports in the Aboriginal child population

The risk of ≥1 screened-in report among Aboriginal children increased from 9-16% during gestation across the 2006 to 2020 birth cohorts, to 41-49% by age five across the 2006 to 2016 birth cohorts (Figure 1; Table E3b). Among children born in 2006, 68% (4,987 children) had ≥1 screened-in report by age 15. Similar to report to the child protection system, the risk of screened-in reports reduced for children born 2011-2012, following the introduction of the Keep Them Safe Initiatives. For children born since 2013, the risk of gestational reports has been trending upwards across consecutive birth cohorts, peaking at 13.9% by age 1 among children born in 2020.

### First-time investigations in the Aboriginal child population

The risk of ≥1 child protection investigation was higher in each consecutive birth cohort from 2006 to 2020, from gestation through childhood (Figure 1; Table E3c). For example, the risk of ≥1 investigation by birth was 1.0% for Aboriginal children born in 2006, compared with 8.0% in the 2020 birth cohort. By age five, the risk of ≥1 investigation was 19% in the 2006 birth cohort, compared with 28% in the 2016 birth cohort. Among the 2006 birth cohort, the risk of ≥1 child protection investigation by age 15 was 39%.

### First-time child protection-defined substantiations in the Aboriginal child population

The risk of ≥1 child protection-defined substantiation by birth increased across consecutive birth cohorts of Aboriginal children, from 0.9% in the 2006 birth cohort to a peak of 4.9% among children born in 2020 (Figure 1; Table E3d). By age five, the risk of ≥1 child protection-defined substantiation ranged from 17-20% across the 2006 to 2016 birth cohorts. Among the 2006 birth cohort, 32% (2,362 children) had ≥1 substantiation by age 15.

### First-time removal into OOHC in the Aboriginal child population

The risk of ≥1 removal into OOHC was 2.6-4.2% by age 1 across the 2006 to 2020 birth cohorts to 7.0-8.7% by age five among the 2006 to 2016 birth cohorts (Figure 1; Table E3e). Among children born in 2006, 14% (1,054 children) had a first-time OOHC placement by age 15. Focussing on the cumulative incidence of Aboriginal children with a first OOHC placement by week of age over the first year of life showed that between 0.4% to 2.1% of children had a first placement by age 2 weeks old and 1.9% to 3.5% had a first placement by age six months (Figure 2).

**Figure 2.**
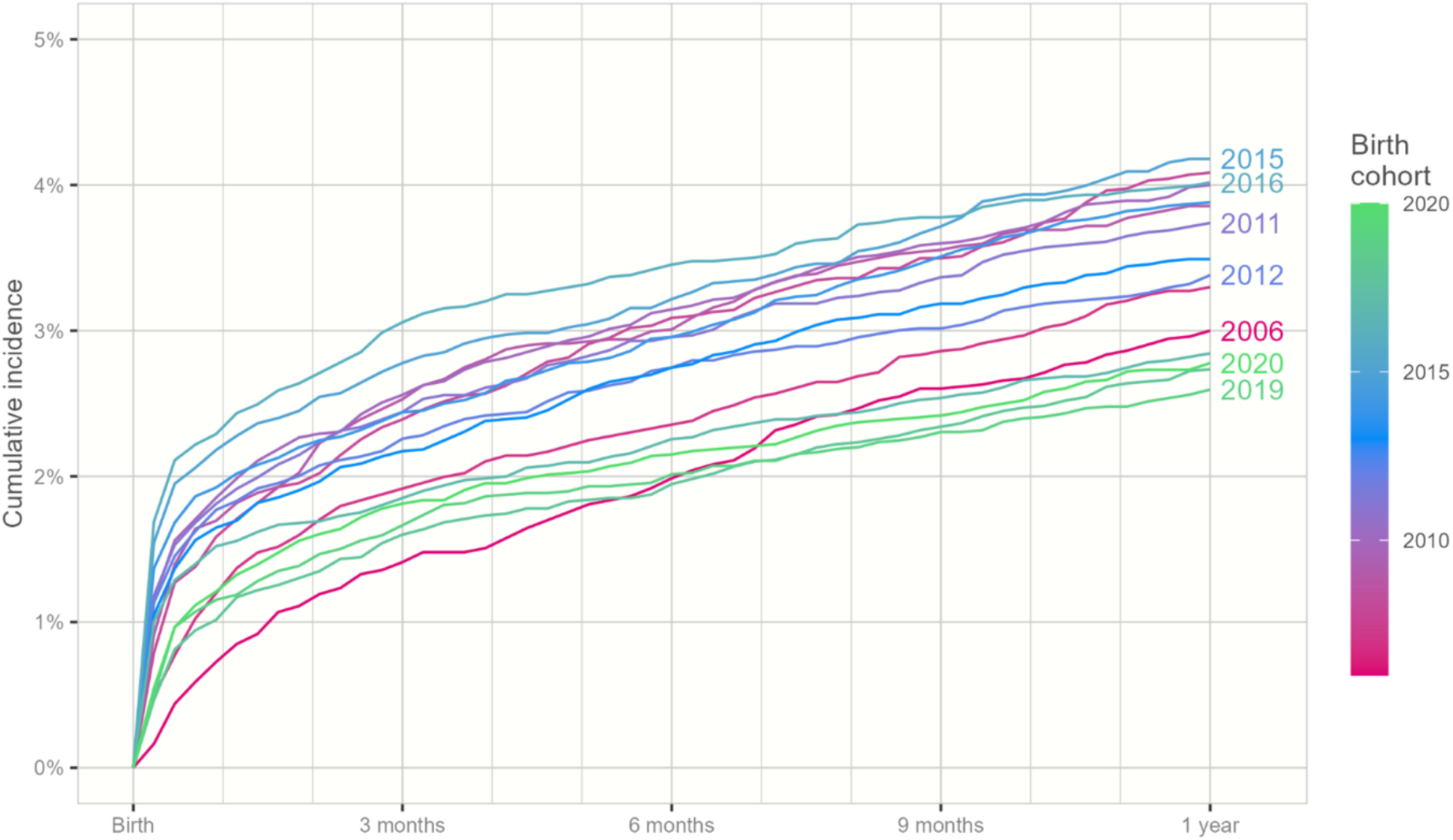
The cumulative incidence of first-time OOHC placements from birth to first birthday for NSW Aboriginal children born 2006–2020

### First-time child protection contacts in Aboriginal and non-Aboriginal child populations

Figure 3 highlights the absolute risk of child protection outcomes for Aboriginal children, alongside non-Aboriginal children, born in 2006, 2011, and 2016 while Figure 4 presents the absolute and relative risks for birth-year cohorts 2006-2020. For example, among 2006 births, the risk of first-time reports to child protection for Aboriginal and non-Aboriginal children, respectively, was 10.5% versus 1.5% by birth (risk difference (RD), 9 percentage points; risk ratio (RR), 7.0), 53% vs 16% by age five (RD, 38pp; RR, 3.4) and 74% vs 33% by age 15 (RD, 41pp; RR 2.2) (Figure 3; Tables E3a and E4a). These large and early gaps persisted in more contemporary cohorts, including the 2011 (RD, 35pp; RR, 3.6 by age five) and 2016 (RD, 35pp; RR, 3.5 by age five) birth-year cohorts.

**Figure 3.**
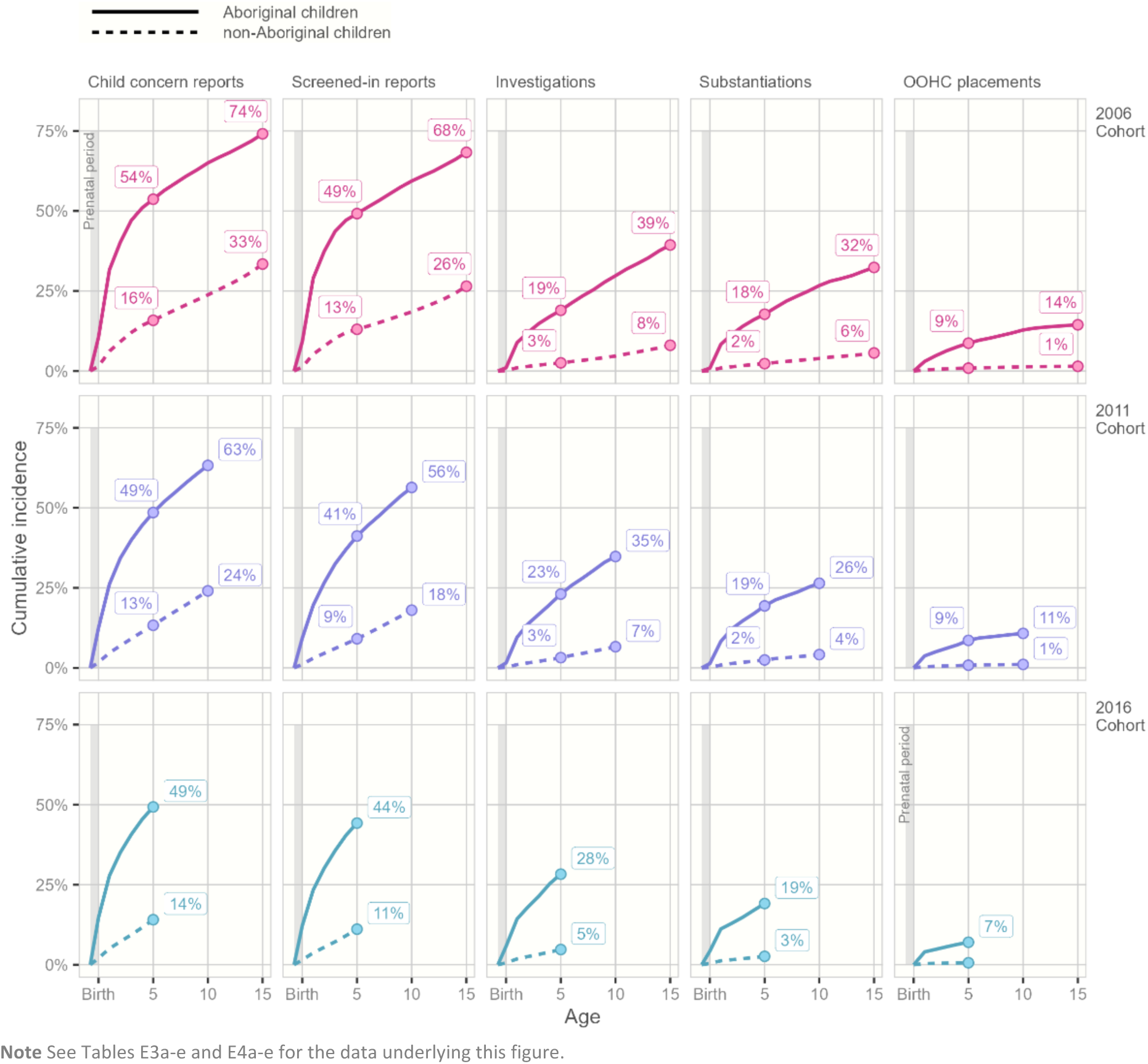
The cumulative incidence of first-time child protection contacts by age for NSW Aboriginal and non-Aboriginal children born 2006, 2011 and 2016, followed from the prenatal period up to age 15 years

**Figure 4.**
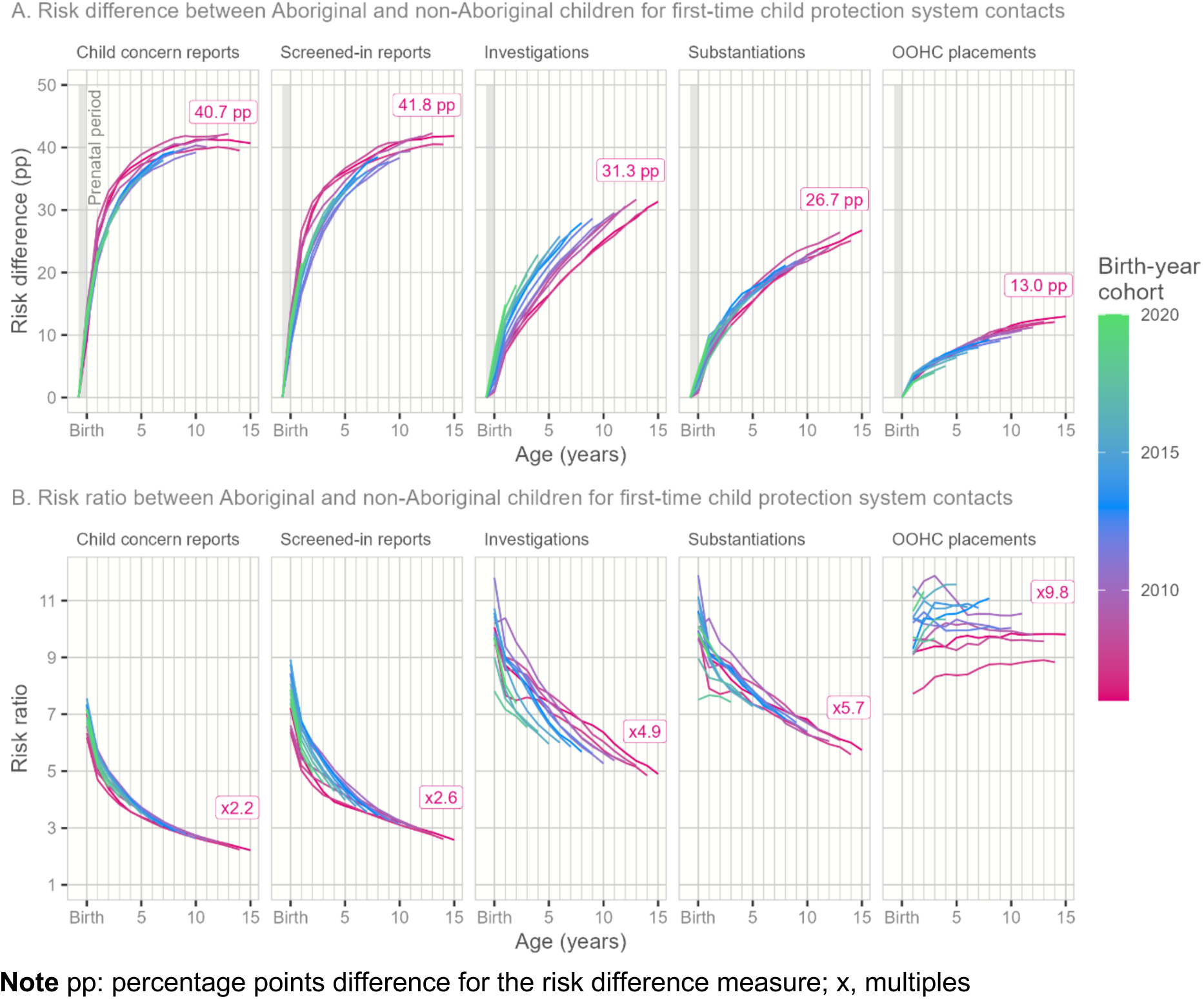
The risk difference (A) and risk ratio (B) of first-time child protection contacts, by year of age, between Aboriginal and non-Aboriginal children born 2006-2020, followed from the prenatal period until age 1 (2018 birth-year cohort) to age 15 (2006 birth-year cohort)

Among 2006 births, the risk of first-time OOHC placements for Aboriginal and non-Aboriginal children by age 1 was 3.0% and 0.33%, respectively (RD, 2.7pp; RR, 9.2), 8.7% vs 0.90% by age five (RD, 7.8pp; RR, 9.7) and 14.4% vs 1.5% by age 15 (RD, 13pp; RR 9.8) (Figure 4).

## Discussion

Our study of almost 120,000 Australian Aboriginal and Torres Strait children, comprising 15 consecutive whole-population birth cohorts from 2006-2020, is the largest and most contemporary study of the lifetime risk of child protection system contact among First Nations children internationally. This Aboriginal-led study has several important findings. First, is the extraordinarily high risk of child protection system contact from the perinatal period to early adolescence for Aboriginal children. By age 15, 3 in 4 Aboriginal children were reported to child protection, 2 in 3 had a screened-in child protection report, 2 in 5 were investigated, 1 in 3 had a child protection-defined substantiation, and 1 in 7 were removed into OOHC, among the 2006 birth cohort. Second, if we are to fulfil public policy goals such as Closing the Gap targets,^12^ it is critical to recognise that Indigenous Data Governance and the approach to analysing data matters. At present, national reporting of child protection data provides the annual ‘system view’ of child protection business. For example, in the 2023/24 financial year, around 3 in 100 Aboriginal children aged 0-17 years had a child protection-defined substantiation and around 1 in 100 were removed into OOHC. In contrast, our child-focused lifetime risk analysis provides actionable evidence to inform prevention responses to better meet the needs of Aboriginal children and families, from pregnancy to adolescence. Third, we show, for the first time, how the absolute inequities in child protection risk between Aboriginal and non-Aboriginal children increase as whole- population cohorts of children age. Finally, the prevention- and community-focused lens we applied to the use of data in this study demonstrates how public health and social policy research, conducted in partnership with Aboriginal organisations and stakeholders, can realise the goals of shared access to data for community, outlined in Priority 4 of the National Agreement on Closing the Gap.^35^

Our study shows the high risk of child protection system contacts for Aboriginal children from early life. For example, by age 1, around 3 in 10 Aboriginal children were reported to child protection, while around 5 in 10 children were first reported by age 5, in the most contemporary birth cohorts followed to these ages. Although there is variation in legislation, policy and practice across jurisdictions, the lifetime risks of child protection contacts were also high among Aboriginal children born 1998-2003 in South Australia (56% by age 17),^26^ Western Australia (52% by age 16, birth cohorts 2000-2013)^18^ and the Northern Territory (46% by age 10, born 1999-2006).^19^ This consistently high risk of child protection contact across Australian jurisdictions reflects the result of pervasive systemic racism and the ‘over-surveillance’ of and intervention in the lives of Aboriginal families and communities.^5,9,27,28^ The well-documented and disproportionate burden of health and social disadvantage experience by Aboriginal children and families are often constructed as ‘risk’ rather than unmet support needs. Our findings highlight the opportunity to respond to such perceived ‘risk’ differently, from the time when family health and social disadvantage is first identified. For example, ante- and post-natal health system contacts are an opportunity to initiate culturally appropriate services that better support the health and social needs of Aboriginal families at risk of child protection intervention.

Our study reveals the secular trends in the lifetime risk of statutory child protection system responses to NSW Aboriginal families across 15 cohorts born from 2006 to 2020. The risk of first-time child protection investigations was higher and earlier in more contemporary birth cohorts of Aboriginal children. For example, by age five, the risk of investigation was 28% in Aboriginal children born in 2016, compared with 19% in the 2006 cohort. Comparable risks of investigations have previously been reported among Aboriginal children in South Australia (e.g. >25% in 2010-2017 births)^26^ and Western Australia (e.g. 25% in 2010-2011 births), with similar increases in risk in more contemporary cohorts.^18^ In our study, 3-4% Aboriginal children were first removed into OOHC by age 1, 7-9% by age 5, and 14% children by age 15, among the most contemporary cohorts to be followed to each age. Comparable risks of removal into OOHC have been reported in South Australia (7-9% by age five),^26^ Western Australia (7.5% by age 5),^18^ and the Northern Territory (7-8% by age ten).^19^ If Australian governments are to fulfil their commitment to Closing the Gap Target 12 by 2031 (a 45 percent reduction in the overrepresentation of Aboriginal children in OOHC since 2019),^12^ then urgent investment is needed in the long-called for Aboriginal-led prevention responses, across the family preservation to restoration continuum,^27,28^ to support families affected by intergenerational disadvantage, state violence and trauma.^5,7,8,36^

For the first time, we demonstrate how the inequities in all levels of child protection system contact between Aboriginal and non-Aboriginal children emerged prenatally, increased in absolute size as whole population cohorts of children aged, and remained similar in magnitude across birth cohorts. While national reporting of child protection statistics routinely shows the higher rates of child protection contact among Aboriginal compared with non-Aboriginal children aged 0-17 years in a financial year period, our lifecourse view of risk provides the actionable intelligence needed to underpin prevention across the lives of children and families. While the specific risk estimates varied, the size of the inequities were large regardless of the data sources used to define the Aboriginal child population denominator. Similar to previous research,^37,38^ we had higher enumeration of Aboriginal children when using more linked datasets for children and parents. When we defined Aboriginality solely from child protection data, the cumulative number of Aboriginal children with child protection contacts was lower, and the size of inequities smaller, especially for reports and screened-in reports. These findings have direct relevance to Closing the Gap target reporting^12^ and the evaluation of prevention initiatives, with likely under-estimation of the number of child protection contacts for Aboriginal children, and the size of ‘the gap’, when using child protection data alone.

### Strengths and limitations

This Aboriginal-led research, conducted by public health and social policy researchers in partnership with Aboriginal organisations and stakeholders, is an international exemplar of actioning the principles of Indigenous Data Governance and Indigenous Data Sovereignty to generate community- and prevention-relevant evidence to inform system reform. In this largest and most contemporary study of the lifetime risk of child protection contact among First Nations children internationally, we provide a model of co-creating prevention-focused evidence to inform investment in culturally appropriate, supportive responses to better meet the health and social needs of Aboriginal families at the earliest opportunity in children’s lives. We also illustrate the potential to better enumerate Aboriginal children for routine child protection and Closing the Gap reporting^12^ by using linked, whole-population datasets from birth through childhood.

A challenge of using administrative datasets is that Aboriginal identification for individuals can vary across data sources and time.^37,38^ Guided by Aboriginal partners and stakeholders, children were included in the Aboriginal population in our study if Aboriginal and/or Torres Strait Islander was recorded for any child and/or parent in the linked datasets. Another challenge in estimating population denominators is the lack of data on the number of individuals who enter or exit the population in each Australian jurisdiction annually. Routine government reporting use estimates of the resident population derived from 5-yearly census data, which provides relatively stable numbers of Aboriginal children across birth cohorts.^30^ In contrast, birth datasets suggest a growing young Aboriginal population in more contemporary cohorts. For this reason, we examined alternative approaches to estimating the NSW Aboriginal child population using linked births and/or child protection datasets, revealing the range of risk estimates generated using different data sources and methods.

## Conclusion

Despite more than 60 inquiries into child protection and related systems, including several prominent Aboriginal-led reviews,^5–8^ and the long-called for shift toward prevention, our study unequivocally shows that the lifetime risk of child protection involvement in the lives of Aboriginal families has not reduced in more contemporary whole-population cohorts. Aboriginal-led responses and investment in early supports for First Nations children and families must be at the centre of system reform due to the persistently high risk of child protection contact for Aboriginal children evident in in this study.^5–8,27^ The accessibility of universal and targeted prevention can be increased by transferring resources, funding, and decision-making powers to Aboriginal families, communities, and organisations. This includes investing in family support services across the spectrum of health, wellbeing and safety needs, designed and delivered by Aboriginal Controlled Community Organisations^7^ and involving families in all decisions regarding their children, at every point of child protection intervention.^28^

## Data Availability

All data produced in the present work are contained in the manuscript and supplemental tables

## Acknowledgements

The NSW Child E-Cohort Project has receiving funding support from a National Health and Medical Research Council (NHMRC) Clinical Trials and Cohort Studies Grant (1187489) and a UNSW Medicine Neuroscience, Mental Health and Addictions Theme and SPHERE Clinical Academic Group seed funding grant. The Bring Them Home, Keep Them Home Project was funded by an ARC Discovery Indigenous grant (194187). This study also received funding through a pilot grant from the Research Excellence in Aboriginal Child and Adolescent Health (REACH) Centre for Research Excellence (1135273).

We thank the children and families of New South Wales (NSW) whose data are included in this study. We thank the NSW Centre for Health Record Linkage for managing and conducting the data linkage for the NSW Child E-Cohort Project (led by K.F.). We also thank the NSW Ministry of Health; NSW Registry of Births, Deaths and Marriages; and NSW Department of Communities and Justice (DCJ) for use of the other data sources in this study. We acknowledge and sincerely thank the research partner organisations for the Bring Them Home, Keep Them Home project, including Waminda—South Coast Women’s Health and Wellbeing Aboriginal Corporation, South Coast Medical Service Aboriginal Corporation, Illawarra Aboriginal Corporation, and the NSW Child, Family and Community Peak Aboriginal Corporation (AbSec). We also acknowledge and sincerely thank the families and the many practitioners across NSW who participated in the Bring Them Home, Keep Them Home research, which informed this study. We thank the Aboriginal Health and Medical Research Council of NSW and the DCJ Aboriginal Reference Group for their input and feedback on this research. We thank the Families and Community Services Insights, Analysis and Research team, Department of Communities and Justice, for their review and feedback on the manuscript. The findings and views reported in this study are those of the authors and should not be attributed to any agency or government department.

## Results

## Main Results

First-time child protection contacts from the prenatal period to age 15 among Aboriginal children born 2006-2020: a whole-population cohort study in New South Wales, Australia

## Online Supplemental Material

## Appendix A. Additional analyses: comparison of results when using different datasets to define the Aboriginal child population

Estimates of the cumulative number of Aboriginal children experiencing first-time child protection contacts are sensitive to the datasets used to code Aboriginal identity. Relying solely on the identity recorded in child protection datasets reduces the number of children identified as Aboriginal and accordingly the cumulative number of contacts over time is lower (Figure E4). This difference is most pronounced for child protection reports and screened-in reports, where least is known about the children and families involved. For example, using the child protection and birth datasets to code Aboriginal identity results in a cumulative number of 5,410 first-time child protection reports by age 15 among the 2006 birth cohort whereas relying solely on the child protection datasets reduces this figure to 4,357 (Figure E4; compare Tables E3a and E5a). The disparity is much smaller when looking at later outcomes such as first-time OOHC placement. The cumulative number of children born in 2006 experiencing first-time OOHC placement by age 15 is 1,054 when CPS and births datasets are used to determine Aboriginal identity, compared to 1,013 when only CPS datasets are used (Figure E4; compare Tables E3e and E5e).

In addition to the datasets used to determine Aboriginal identity, the outcomes for the cumulative incidence and Aboriginal to non-Aboriginal gap are sensitive to the source of data used to estimate the Aboriginal and non-Aboriginal population denominator. Figure E5 presents the cumulative incidence of first-time CPS contacts for Aboriginal and non-Aboriginal children born in 2006 based on two possible combinations of numerator and denominator source data: (i) the numerator based solely on Aboriginality in the CPS datasets and the denominator based on the ABS Estimated Residential population, and (ii) the numerator based on Aboriginality in the CPS and NSW births datasets and the denominator based on the NSW born population and CPS contacts). The former option reflects what would be available using unlinked child protection data while the latter option is what we used for the main analysis. Under the first definition, the cumulative incidence of first-time reports by age 15 is 72% for Aboriginal children and 40% for non-Aboriginal children, compared to 74% and 33% respectively in the main analysis. The difference between the two approaches is less pronounced in the more serious types of child protection contact.

---

**Figure E1.**
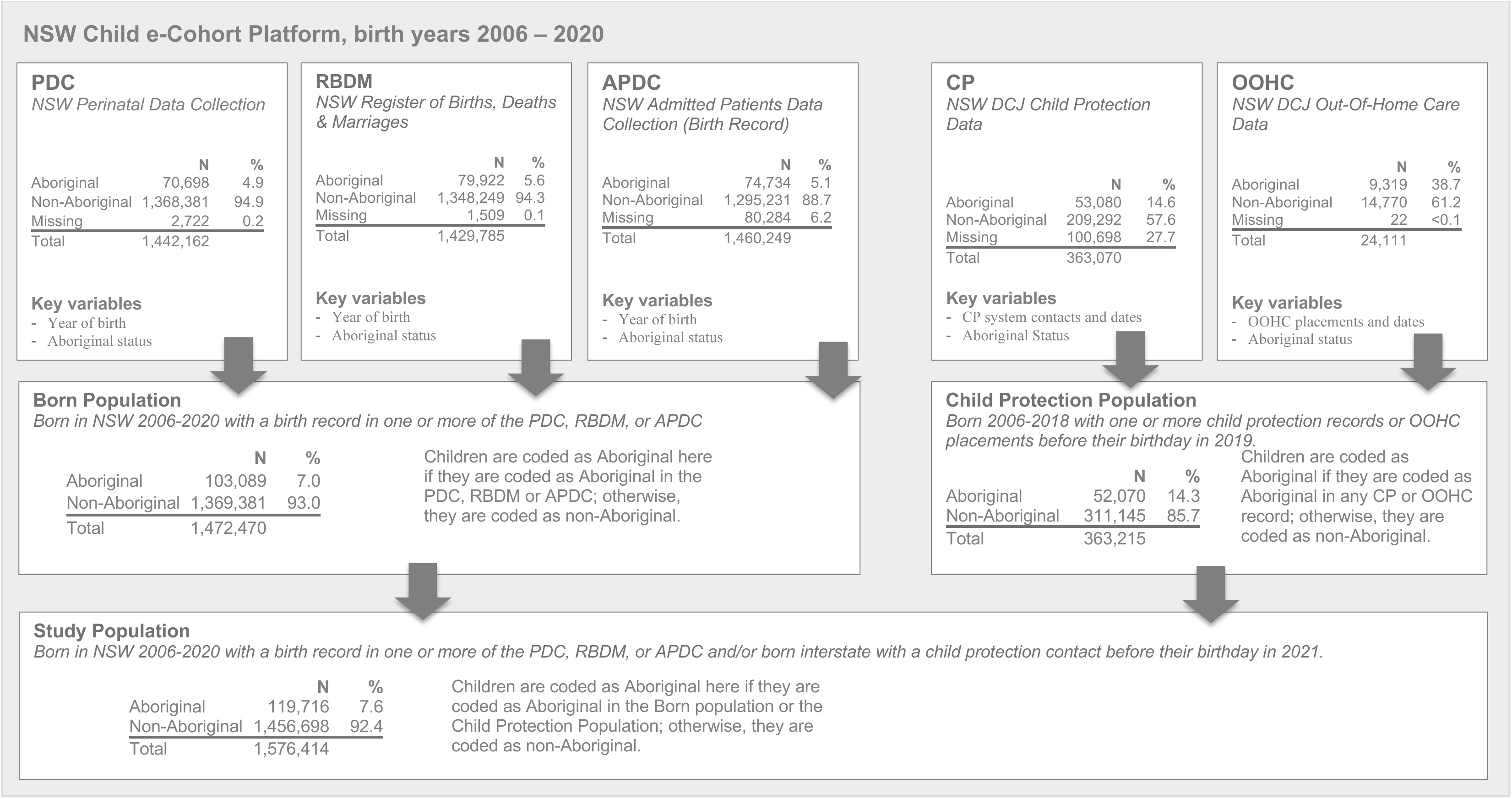
Flowchart defining the study population from birth and child protection datasets available in the NSW Child E-Cohort Platform

**Figure E2.**
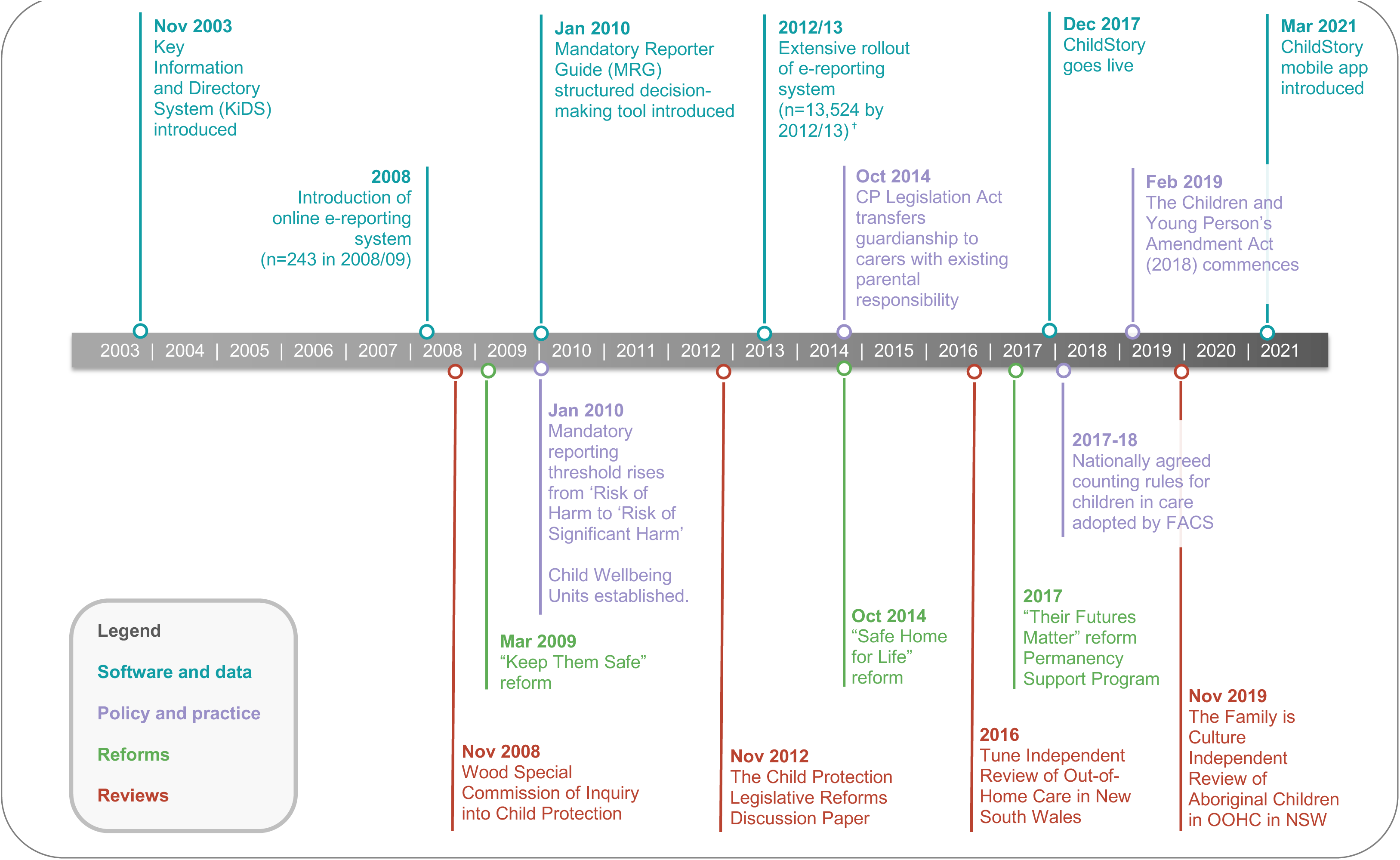
Timeline of major New South Wales Child Protection System, software, policy, reforms, and reviews 2003 – 2021

**Figure E3.**
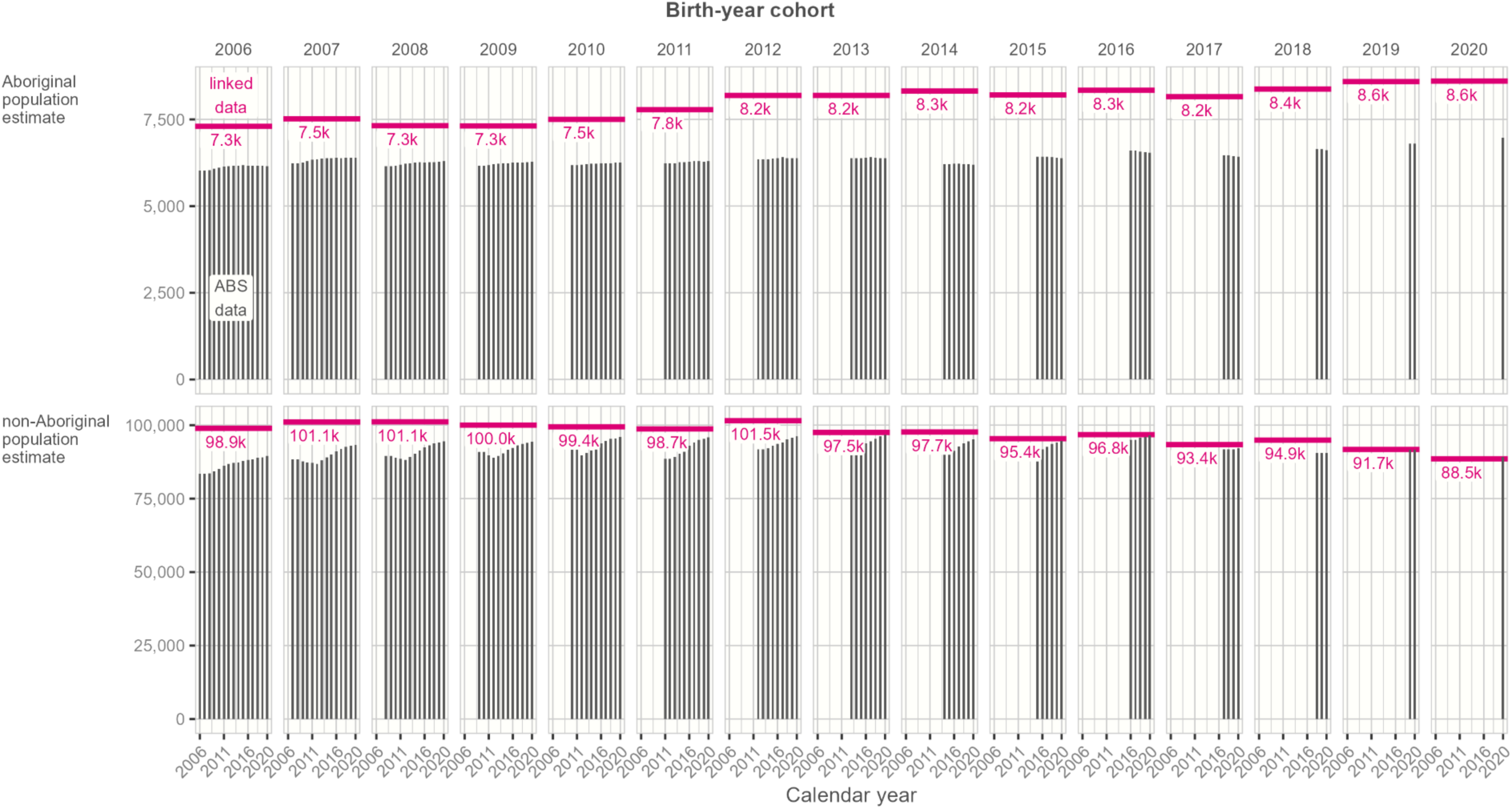
Population denominator estimates by cohort year (2006-2020) over time for Aboriginal and non-Aboriginal children in NSW based on (i) Australian Bureau of Statistics data^1^ and linked birth and child protection datasets from the NSW Child e-Cohort data platform^2^ ^1^ Aboriginal population estimates from Australian Bureau of Statistics. Estimates and Projections, Aboriginal and Torres Strait Islander Australians, 2001 to 2026 (Catalogue No. 3238.0). 2014. Accessed January 16, 2025. https://www.abs.gov.au/AUSSTATS/abs@.nsf/DetailsPage/3238.02001%20to%202026 and total population estimates from Australian Bureau of Statistics. Australian Demographic Statistics (Catalogue No. 3101.0). 2019. Accessed January 16, 2025. https://www.abs.gov.au/ausstats/abs@.nsf/exnote/3101.0 ^2^ Linked data estimates include all individuals with one or more linked records in the NSW Perinatal Data Collection (PDC), the NSW Admitted Patients Data Collection (APDC), the NSW Registry of Births Deaths and Marriages (RBDM) birth records, the Department of Communities and Justice (DCJ) Child Protection records and Out-of-home Care (OOHC) records.

**Figure E4.**
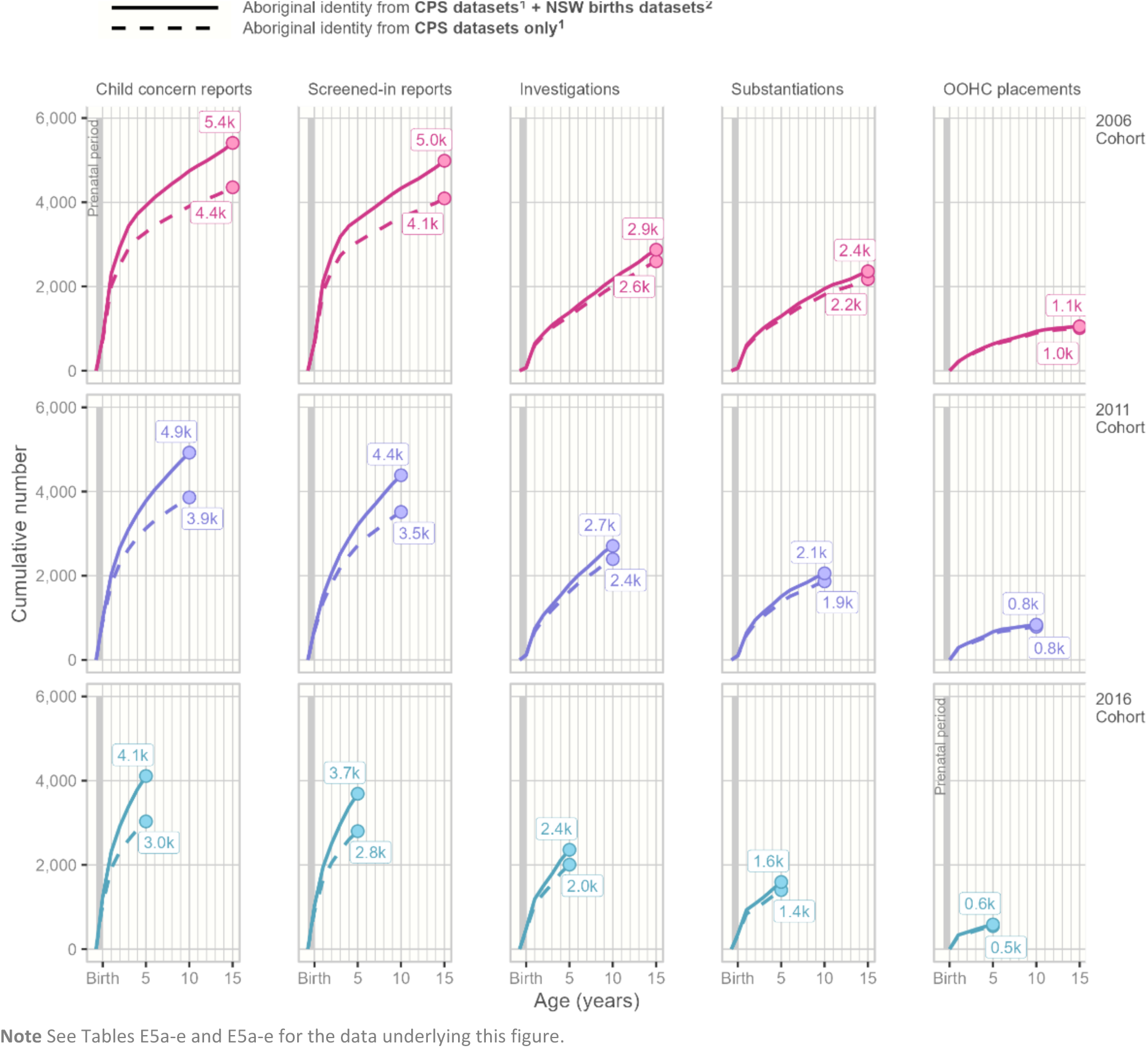
Cumulative number of first-time child protection system contacts for Aboriginal children living in NSW and born in 2006, 2011, and 2016, by data sources used to code a child’s Aboriginality ^1^ Children are coded as Aboriginal if (i) they or their parents are recorded as Aboriginal in any birth dataset, including the NSW Perinatal Data Collection (PDC), the NSW Registry of Births Deaths and Marriages (RBDM) birth records, or the NSW Admitted Patients Data Collection (APDC) birth record, or (ii) if the child is recorded as Aboriginal in the Department of Communities and Justice (DCJ) Child Protection or Out-of-home Care (OOHC) records from the prenatal period until 2021. ^2^ Children are coded as Aboriginal if they are recorded as Aboriginal in the Department of Communities and Justice (DCJ) Child Protection or Out-of-home Care (OOHC) records from the prenatal period until 2021.

**Figure E5.**
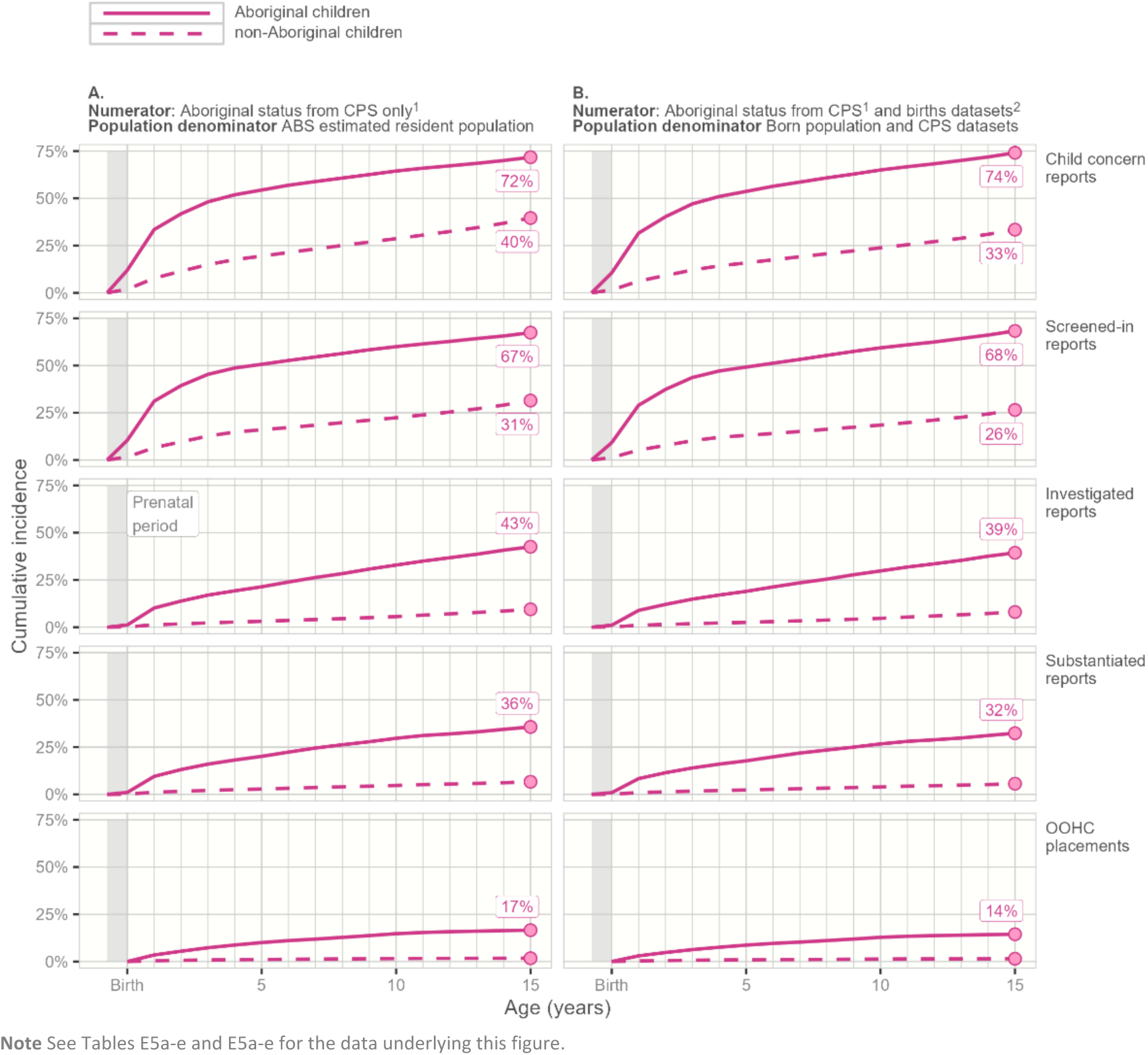
Cumulative incidence of first-time child protection system contacts for Aboriginal children living in NSW and born in 2006, by data sources used to code a child’s Aboriginality and data sources used to estimate the population denominator ^1^ Children are coded as Aboriginal if they are recorded as Aboriginal in the Department of Communities and Justice (DCJ) Child Protection or Out-of-home Care (OOHC) records from the prenatal period until 2021. ^2^ Children are coded as Aboriginal if (i) they or their parents are recorded as Aboriginal in any birth dataset, including the NSW Perinatal Data Collection (PDC), the NSW Registry of Births Deaths and Marriages (RBDM) birth records, or the NSW Admitted Patients Data Collection (APDC) birth record, or (ii) if the child is recorded as Aboriginal in the Department of Communities and Justice (DCJ) Child Protection or Out-of-home Care (OOHC) records from the prenatal period until 2021. ^3^ Population denominator based on Australian Bureau of Statistics population estimates (See Figure E3 and Tables E2a-E2b) ^4^ Population denominator based on NSW child e-Cohort data platform (See Figure E3 and Tables E2a-E2b)

**Table E1.**
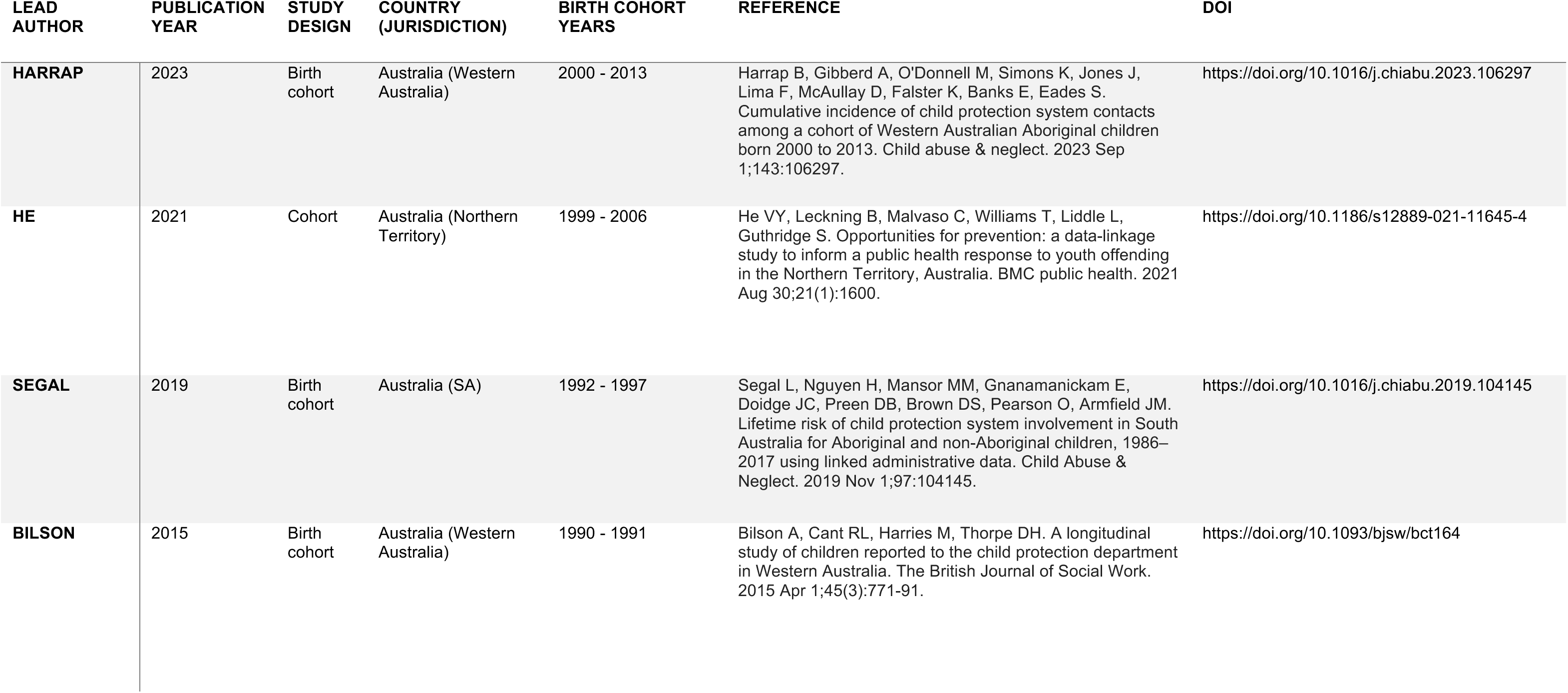
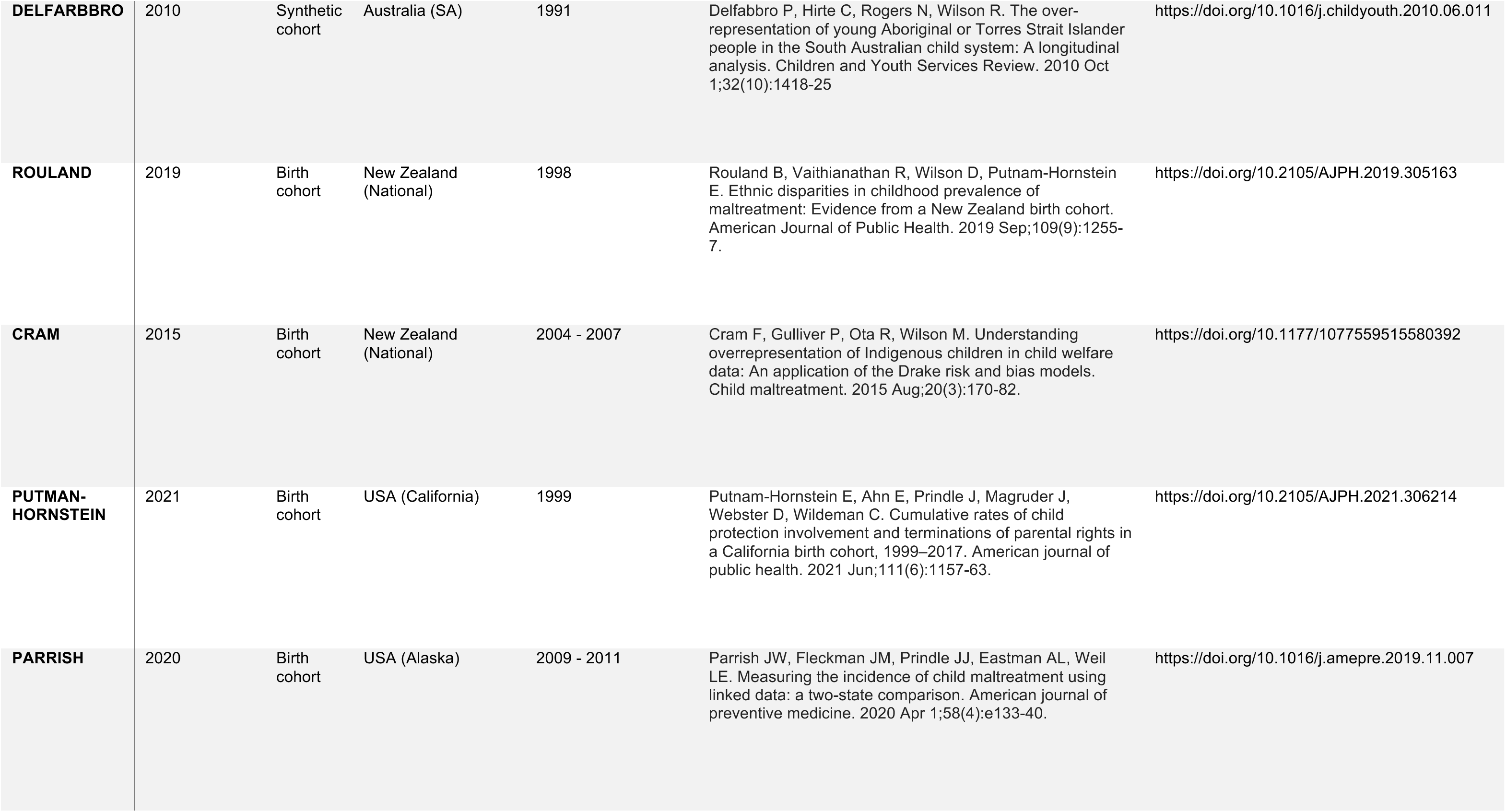
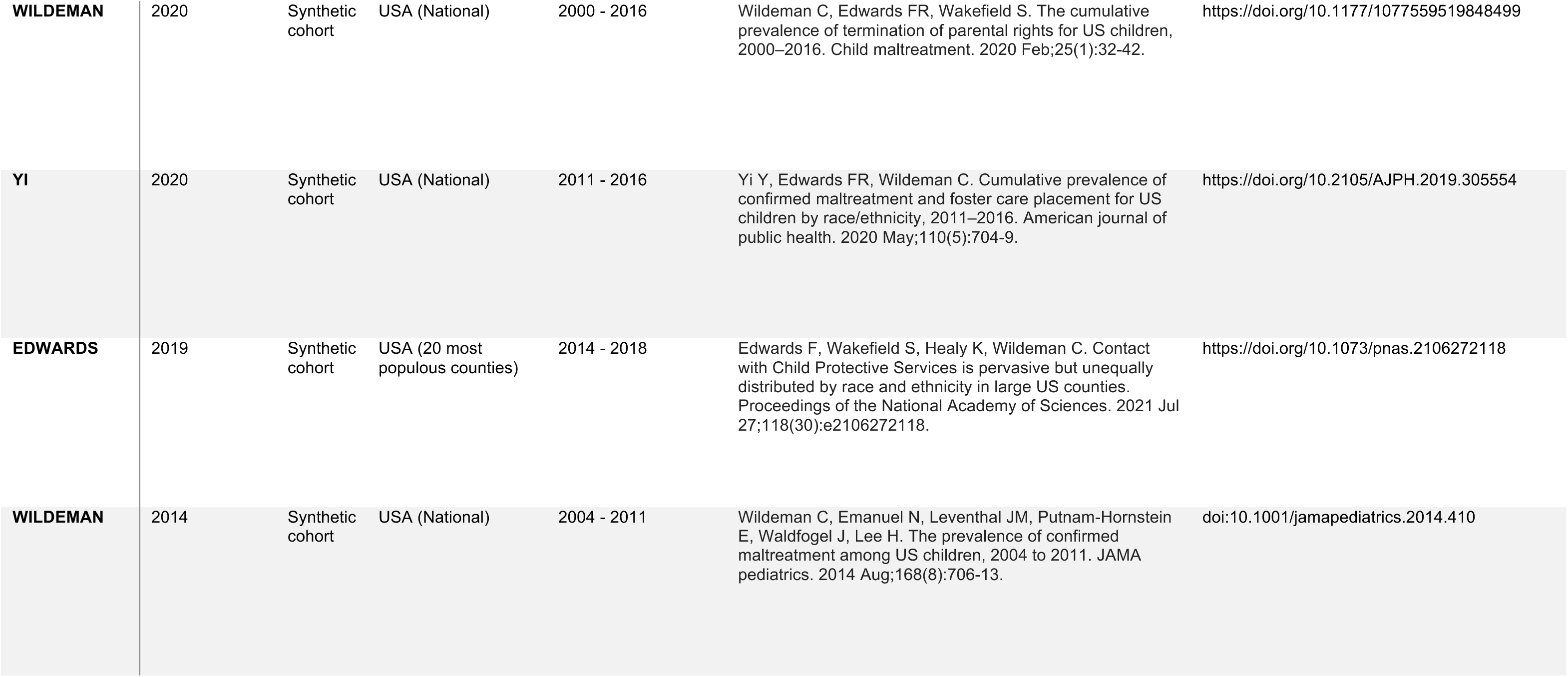
Systematic Review Results: Details of 13 studies on the lifetime risk of child protection contacts among First Nations child populations, published January 2005 to May 2025.

**Table E2a.** Estimated population denominator for Aboriginal children living in NSW (2006- 2018) based on (i) Australian Bureau of Statistics (ABS) estimates^1^ and (ii) Linked birth and child protection datasets from the NSW Child e-Cohort platform^2^.

| Birth-year cohort | Denominator source | Calendar year |  |  |  |  |  |  |  |  |  |  |  |  |  |  |  |
| --- | --- | --- | --- | --- | --- | --- | --- | --- | --- | --- | --- | --- | --- | --- | --- | --- | --- |
|  |  | 2006 | 2007 | 2008 | 2009 | 2010 | 2011 | 2012 | 2013 | 2014 | 2015 | 2016 | 2017 | 2018 | 2019 | 2020 | 2021 |
| 2006 | ABS data | 6,024 | 6,024 | 6,038 | 6,074 | 6,110 | 6,136 | 6,157 | 6,172 | 6,170 | 6,174 | 6,164 | 6,160 | 6,167 | 6,167 | 6,156 | 6,140 |
|  | Linked data | 7,300 | 7,300 | 7,300 | 7,300 | 7,300 | 7,300 | 7,300 | 7,300 | 7,300 | 7,300 | 7,300 | 7,300 | 7,300 | 7,300 | 7,300 | 7,300 |
| 2007 | ABS data |  | 6,243 | 6,243 | 6,257 | 6,294 | 6,332 | 6,359 | 6,370 | 6,387 | 6,387 | 6,392 | 6,380 | 6,390 | 6,398 | 6,396 | 6,384 |
|  | Linked data |  | 7,517 | 7,517 | 7,517 | 7,517 | 7,517 | 7,517 | 7,517 | 7,517 | 7,517 | 7,517 | 7,517 | 7,517 | 7,517 | 7,517 | 7,517 |
| 2008 | ABS data |  |  | 6,147 | 6,147 | 6,159 | 6,192 | 6,226 | 6,243 | 6,250 | 6,263 | 6,259 | 6,262 | 6,266 | 6,280 | 6,289 | 6,290 |
|  | Linked data |  |  | 7,320 | 7,320 | 7,320 | 7,320 | 7,320 | 7,320 | 7,320 | 7,320 | 7,320 | 7,320 | 7,320 | 7,320 | 7,320 | 7,320 |
| 2009 | ABS data |  |  |  | 6,161 | 6,161 | 6,173 | 6,206 | 6,219 | 6,235 | 6,241 | 6,251 | 6,246 | 6,258 | 6,262 | 6,277 | 6,286 |
|  | Linked data |  |  |  | 7,314 | 7,314 | 7,314 | 7,314 | 7,314 | 7,314 | 7,314 | 7,314 | 7,314 | 7,314 | 7,314 | 7,314 | 7,314 |
| 2010 | ABS data |  |  |  |  | 6,182 | 6,182 | 6,193 | 6,205 | 6,217 | 6,230 | 6,234 | 6,245 | 6,240 | 6,252 | 6,257 | 6,272 |
|  | Linked data |  |  |  |  | 7,502 | 7,502 | 7,502 | 7,502 | 7,502 | 7,502 | 7,502 | 7,502 | 7,502 | 7,502 | 7,502 | 7,502 |
| 2011 | ABS data |  |  |  |  |  | 6,240 | 6,240 | 6,245 | 6,260 | 6,273 | 6,288 | 6,293 | 6,294 | 6,288 | 6,298 | 6,302 |
|  | Linked data |  |  |  |  |  | 7,782 | 7,782 | 7,782 | 7,782 | 7,782 | 7,782 | 7,782 | 7,782 | 7,782 | 7,782 | 7,782 |
| 2012 | ABS data |  |  |  |  |  |  | 6,348 | 6,348 | 6,353 | 6,371 | 6,388 | 6,404 | 6,387 | 6,386 | 6,379 | 6,388 |
|  | Linked data |  |  |  |  |  |  | 8,192 | 8,192 | 8,192 | 8,192 | 8,192 | 8,192 | 8,192 | 8,192 | 8,192 | 8,192 |
| 2013 | ABS data |  |  |  |  |  |  |  | 6,376 | 6,376 | 6,381 | 6,396 | 6,411 | 6,402 | 6,386 | 6,387 | 6,382 |
|  | Linked data |  |  |  |  |  |  |  | 8,193 | 8,193 | 8,193 | 8,193 | 8,193 | 8,193 | 8,193 | 8,193 | 8,193 |
| 2014 | ABS data |  |  |  |  |  |  |  |  | 6,212 | 6,212 | 6,217 | 6,230 | 6,214 | 6,206 | 6,193 | 6,195 |
|  | Linked data |  |  |  |  |  |  |  |  | 8,321 | 8,321 | 8,321 | 8,321 | 8,321 | 8,321 | 8,321 | 8,321 |
| 2015 | ABS data |  |  |  |  |  |  |  |  |  | 6,423 | 6,423 | 6,426 | 6,409 | 6,391 | 6,382 | 6,367 |
|  | Linked data |  |  |  |  |  |  |  |  |  | 8,208 | 8,208 | 8,208 | 8,208 | 8,208 | 8,208 | 8,208 |
| 2016 | ABS data |  |  |  |  |  |  |  |  |  |  | 6,597 | 6,597 | 6,574 | 6,554 | 6,534 | 6,521 |
|  | Linked data |  |  |  |  |  |  |  |  |  |  | 8,340 | 8,340 | 8,340 | 8,340 | 8,340 | 8,340 |
| 2017 | ABS data |  |  |  |  |  |  |  |  |  |  |  | 6,467 | 6,467 | 6,447 | 6,431 | 6,413 |
|  | Linked data |  |  |  |  |  |  |  |  |  |  |  | 8,157 | 8,157 | 8,157 | 8,157 | 8,157 |
| 2018 | ABS data |  |  |  |  |  |  |  |  |  |  |  |  | 6,637 | 6,637 | 6,615 | 6,599 |
|  | Linked data |  |  |  |  |  |  |  |  |  |  |  |  | 8,374 | 8,374 | 8,374 | 8,374 |
| 2019 | ABS data |  |  |  |  |  |  |  |  |  |  |  |  |  | 6,802 | 6,802 | 6,780 |
|  | Linked data |  |  |  |  |  |  |  |  |  |  |  |  |  | 8,592 | 8,592 | 8,592 |
| 2020 | ABS data |  |  |  |  |  |  |  |  |  |  |  |  |  |  | 6,970 | 6,970 |
|  | Linked data |  |  |  |  |  |  |  |  |  |  |  |  |  |  | 8,604 | 8,604 |

<sup>1</sup> Aboriginal population estimates from Australian Bureau of Statistics. Estimates and Projections, Aboriginal and Torres Strait Islander Australians, 2001 to 2026 (Catalogue No. 3238.0). 2014. Accessed January 16, 2025.
https://www.abs.gov.au/AUSSTATS/abs@.nsf/DetailsPage/3238.02001%20to%202026 and total population estimates from Australian Bureau of Statistics. Australian Demographic Statistics (Catalogue No. 3101.0). 2019. Accessed January 16, 2025.
https://www.abs.gov.au/ausstats/abs@.nsf/exnote/3101.0
<sup>2</sup> Linked data estimates include all individuals with one or more linked records in the NSW Perinatal Data Collection (PDC), the NSW Admitted Patients Data Collection (APDC), the NSW Registry of Births Deaths and Marriages (RBDM) birth records, the Department of Communities and Justice (DCJ) Child Protection records and Out-of-home Care (OOHC) records.

**Table E2b.** Estimated population denominator for non-Aboriginal children living in NSW (2006-2021) based on (i) Australian Bureau of Statistics (ABS) estimates^1^ and (ii) Linked birth and child protection datasets from the NSW Child e-Cohort platform^2^.

| Birth-year cohort | Denominator source | Calendar year |  |  |  |  |  |  |  |  |  |  |  |  |  |  |  |
| --- | --- | --- | --- | --- | --- | --- | --- | --- | --- | --- | --- | --- | --- | --- | --- | --- | --- |
|  |  | 2006 | 2007 | 2008 | 2009 | 2010 | 2011 | 2012 | 2013 | 2014 | 2015 | 2016 | 2017 | 2018 | 2019 | 2020 | 2021 |
| 2006 | ABS data | 83,477 | 83,477 | 83,534 | 84,248 | 85,174 | 86,143 | 86,873 | 87,096 | 87,382 | 87,770 | 88,164 | 88,352 | 88,844 | 89,079 | 89,477 | 89,931 |
|  | Linked data | 98,906 | 98,906 | 98,906 | 98,906 | 98,906 | 98,906 | 98,906 | 98,906 | 98,906 | 98,906 | 98,906 | 98,906 | 98,906 | 98,906 | 98,906 | 98,906 |
| 2007 | ABS data |  | 88,407 | 88,407 | 87,721 | 87,371 | 87,142 | 86,858 | 87,978 | 89,096 | 90,150 | 91,095 | 91,999 | 92,671 | 92,881 | 93,235 | 93,512 |
|  | Linked data |  | 101,088 | 101,088 | 101,088 | 101,088 | 101,088 | 101,088 | 101,088 | 101,088 | 101,088 | 101,088 | 101,088 | 101,088 | 101,088 | 101,088 | 101,088 |
| 2008 | ABS data |  |  | 89,495 | 89,495 | 88,906 | 88,574 | 88,141 | 89,159 | 90,254 | 91,431 | 92,413 | 93,175 | 93,879 | 94,173 | 94,531 | 94,924 |
|  | Linked data |  |  | 101,134 | 101,134 | 101,134 | 101,134 | 101,134 | 101,134 | 101,134 | 101,134 | 101,134 | 101,134 | 101,134 | 101,134 | 101,134 | 101,134 |
| 2009 | ABS data |  |  |  | 90,996 | 90,996 | 89,778 | 88,863 | 89,583 | 90,388 | 91,518 | 92,407 | 93,071 | 93,690 | 93,970 | 94,391 | 94,715 |
|  | Linked data |  |  |  | 100,031 | 100,031 | 100,031 | 100,031 | 100,031 | 100,031 | 100,031 | 100,031 | 100,031 | 100,031 | 100,031 | 100,031 | 100,031 |
| 2010 | ABS data |  |  |  |  | 91,589 | 91,589 | 89,741 | 90,627 | 91,514 | 92,642 | 93,775 | 94,647 | 95,294 | 95,530 | 96,011 | 96,393 |
|  | Linked data |  |  |  |  | 99,421 | 99,421 | 99,421 | 99,421 | 99,421 | 99,421 | 99,421 | 99,421 | 99,421 | 99,421 | 99,421 | 99,421 |
| 2011 | ABS data |  |  |  |  |  | 88,578 | 88,578 | 89,135 | 90,284 | 91,686 | 92,961 | 94,137 | 94,975 | 95,377 | 95,819 | 96,187 |
|  | Linked data |  |  |  |  |  | 98,730 | 98,730 | 98,730 | 98,730 | 98,730 | 98,730 | 98,730 | 98,730 | 98,730 | 98,730 | 98,730 |
| 2012 | ABS data |  |  |  |  |  |  | 91,829 | 91,829 | 92,070 | 92,881 | 93,614 | 94,119 | 95,186 | 95,618 | 96,140 | 96,579 |
|  | Linked data |  |  |  |  |  |  | 101,515 | 101,515 | 101,515 | 101,515 | 101,515 | 101,515 | 101,515 | 101,515 | 101,515 | 101,515 |
| 2013 | ABS data |  |  |  |  |  |  |  | 92,851 | 92,851 | 93,177 | 93,823 | 94,411 | 95,407 | 96,211 | 96,639 | 97,047 |
|  | Linked data |  |  |  |  |  |  |  | 97,510 | 97,510 | 97,510 | 97,510 | 97,510 | 97,510 | 97,510 | 97,510 | 97,510 |
| 2014 | ABS data |  |  |  |  |  |  |  |  | 90,186 | 90,186 | 91,314 | 92,844 | 93,883 | 94,513 | 95,219 | 95,790 |
|  | Linked data |  |  |  |  |  |  |  |  | 97,680 | 97,680 | 97,680 | 97,680 | 97,680 | 97,680 | 97,680 | 97,680 |
| 2015 | ABS data |  |  |  |  |  |  |  |  |  | 91,689 | 91,689 | 92,682 | 93,593 | 94,071 | 94,745 | 95,438 |
|  | Linked data |  |  |  |  |  |  |  |  |  | 95,389 | 95,389 | 95,389 | 95,389 | 95,389 | 95,389 | 95,389 |
| 2016 | ABS data |  |  |  |  |  |  |  |  |  |  | 95,007 | 95,007 | 95,649 | 96,017 | 96,484 | 96,995 |
|  | Linked data |  |  |  |  |  |  |  |  |  |  | 96,774 | 96,774 | 96,774 | 96,774 | 96,774 | 96,774 |
| 2017 | ABS data |  |  |  |  |  |  |  |  |  |  |  | 91,728 | 91,728 | 91,699 | 92,094 | 92,391 |
|  | Linked data |  |  |  |  |  |  |  |  |  |  |  | 93,376 | 93,376 | 93,376 | 93,376 | 93,376 |
| 2018 | ABS data |  |  |  |  |  |  |  |  |  |  |  |  | 90,598 | 90,598 | 90,553 | 90,668 |
|  | Linked data |  |  |  |  |  |  |  |  |  |  |  |  | 94,892 | 94,892 | 94,892 | 94,892 |
| 2019 | ABS data |  |  |  |  |  |  |  |  |  |  |  |  |  | 91,634 | 91,634 | 91,473 |
|  | Linked data |  |  |  |  |  |  |  |  |  |  |  |  |  | 91,731 | 91,731 | 91,731 |
| 2020 | ABS data |  |  |  |  |  |  |  |  |  |  |  |  |  |  | 89,290 | 89,290 |
|  | Linked data |  |  |  |  |  |  |  |  |  |  |  |  |  |  | 88,521 | 88,521 |

<sup>1</sup> Aboriginal population estimates from Australian Bureau of Statistics. Estimates and Projections, Aboriginal and Torres Strait Islander Australians, 2001 to 2026 (Catalogue No. 3238.0). 2014. Accessed January 16, 2025. https://www.abs.gov.au/AUSSTATS/abs@.nsf/DetailsPage/3238.02001%20to%202026 and total population estimates from Australian Bureau of Statistics. Australian Demographic Statistics (Catalogue No. 3101.0). 2019. Accessed January 16, 2025. https://www.abs.gov.au/ausstats/abs@.nsf/exnote/3101.0
<sup>2</sup> Linked data estimates include all individuals with one or more linked records in the NSW Perinatal Data Collection (PDC), the NSW Admitted Patients Data Collection (APDC), the NSW Registry of Births Deaths and Marriages (RBDM) birth records, the Department of Communities and Justice (DCJ) Child Protection records and Out-of-home Care (OOHC) records.

**Table E3a.** Number, cumulative number and cumulative incidence of first-time child protection reports by year of age for Aboriginal children born 2006-2020.

| Birth-year cohort |  | Age |  |  |  |  |  |  |  |  |  |  |  |  |  |  |  |
| --- | --- | --- | --- | --- | --- | --- | --- | --- | --- | --- | --- | --- | --- | --- | --- | --- | --- |
|  |  | Birth | 1 | 2 | 3 | 4 | 5 | 6 | 7 | 8 | 9 | 10 | 11 | 12 | 13 | 14 | 15 |
| 2006 | N | 769 | 1,538 | 633 | 496 | 286 | 196 | 198 | 162 | 163 | 146 | 159 | 125 | 114 | 130 | 133 | 162 |
|  | Cumulative N | 769 | 2,307 | 2,940 | 3,436 | 3,722 | 3,918 | 4,116 | 4,278 | 4,441 | 4,587 | 4,746 | 4,871 | 4,985 | 5,115 | 5,248 | 5,410 |
|  | Cumulative % | 10.5 | 31.6 | 40.3 | 47.1 | 51.0 | 53.7 | 56.4 | 58.6 | 60.8 | 62.8 | 65.0 | 66.7 | 68.3 | 70.1 | 71.9 | 74.1 |
| 2007 | N | 973 | 1,488 | 691 | 381 | 227 | 223 | 168 | 176 | 169 | 146 | 154 | 140 | 143 | 135 | 151 |  |
|  | Cumulative N | 973 | 2,461 | 3,152 | 3,533 | 3,760 | 3,983 | 4,151 | 4,327 | 4,496 | 4,642 | 4,796 | 4,936 | 5,079 | 5,214 | 5,365 |  |
|  | Cumulative % | 12.9 | 32.7 | 41.9 | 47.0 | 50.0 | 53.0 | 55.2 | 57.6 | 59.8 | 61.8 | 63.8 | 65.7 | 67.6 | 69.4 | 71.4 |  |
| 2008 | N | 1,121 | 1,474 | 515 | 299 | 294 | 238 | 208 | 195 | 180 | 161 | 125 | 135 | 153 | 184 |  |  |
|  | Cumulative N | 1,121 | 2,595 | 3,110 | 3,409 | 3,703 | 3,941 | 4,149 | 4,344 | 4,524 | 4,685 | 4,810 | 4,945 | 5,098 | 5,282 |  |  |
|  | Cumulative % | 15.3 | 35.5 | 42.5 | 46.6 | 50.6 | 53.8 | 56.7 | 59.3 | 61.8 | 64.0 | 65.7 | 67.6 | 69.6 | 72.2 |  |  |
| 2009 | N | 1,238 | 1,175 | 433 | 350 | 249 | 284 | 258 | 222 | 200 | 142 | 171 | 173 | 168 |  |  |  |
|  | Cumulative N | 1,238 | 2,413 | 2,846 | 3,196 | 3,445 | 3,729 | 3,987 | 4,209 | 4,409 | 4,551 | 4,722 | 4,895 | 5,063 |  |  |  |
|  | Cumulative % | 16.9 | 33.0 | 38.9 | 43.7 | 47.1 | 51.0 | 54.5 | 57.5 | 60.3 | 62.2 | 64.6 | 66.9 | 69.2 |  |  |  |
| 2010 | N | 1,089 | 1,002 | 539 | 442 | 336 | 302 | 253 | 235 | 192 | 191 | 207 | 157 |  |  |  |  |
|  | Cumulative N | 1,089 | 2,091 | 2,630 | 3,072 | 3,408 | 3,710 | 3,963 | 4,198 | 4,390 | 4,581 | 4,788 | 4,945 |  |  |  |  |
|  | Cumulative % | 14.5 | 27.9 | 35.1 | 40.9 | 45.4 | 49.5 | 52.8 | 56.0 | 58.5 | 61.1 | 63.8 | 65.9 |  |  |  |  |
| 2011 | N | 970 | 1,066 | 634 | 438 | 360 | 308 | 265 | 225 | 232 | 218 | 206 |  |  |  |  |  |
|  | Cumulative N | 970 | 2,036 | 2,670 | 3,108 | 3,468 | 3,776 | 4,041 | 4,266 | 4,498 | 4,716 | 4,922 |  |  |  |  |  |
|  | Cumulative % | 12.5 | 26.2 | 34.3 | 39.9 | 44.6 | 48.5 | 51.9 | 54.8 | 57.8 | 60.6 | 63.2 |  |  |  |  |  |
| 2012 | N | 1,040 | 1,154 | 656 | 471 | 385 | 317 | 285 | 286 | 244 | 236 |  |  |  |  |  |  |
|  | Cumulative N | 1,040 | 2,194 | 2,850 | 3,321 | 3,706 | 4,023 | 4,308 | 4,594 | 4,838 | 5,074 |  |  |  |  |  |  |
|  | Cumulative % | 12.7 | 26.8 | 34.8 | 40.5 | 45.2 | 49.1 | 52.6 | 56.1 | 59.1 | 61.9 |  |  |  |  |  |  |
| 2013 | N | 1,134 | 1,150 | 611 | 486 | 393 | 304 | 318 | 294 | 236 |  |  |  |  |  |  |  |
|  | Cumulative N | 1,134 | 2,284 | 2,895 | 3,381 | 3,774 | 4,078 | 4,396 | 4,690 | 4,926 |  |  |  |  |  |  |  |
|  | Cumulative % | 13.8 | 27.9 | 35.3 | 41.3 | 46.1 | 49.8 | 53.7 | 57.2 | 60.1 |  |  |  |  |  |  |  |
| 2014 | N | 1,168 | 1,148 | 625 | 491 | 322 | 312 | 327 | 301 |  |  |  |  |  |  |  |  |
|  | Cumulative N | 1,168 | 2,316 | 2,941 | 3,432 | 3,754 | 4,066 | 4,393 | 4,694 |  |  |  |  |  |  |  |  |
|  | Cumulative % | 14.0 | 27.8 | 35.3 | 41.2 | 45.1 | 48.9 | 52.8 | 56.4 |  |  |  |  |  |  |  |  |
| 2015 | N | 1,183 | 1,146 | 571 | 443 | 380 | 341 | 334 |  |  |  |  |  |  |  |  |  |
|  | Cumulative N | 1,183 | 2,329 | 2,900 | 3,343 | 3,723 | 4,064 | 4,398 |  |  |  |  |  |  |  |  |  |
|  | Cumulative % | 14.4 | 28.4 | 35.3 | 40.7 | 45.4 | 49.5 | 53.6 |  |  |  |  |  |  |  |  |  |
| 2016 | N | 1,227 | 1,092 | 608 | 467 | 397 | 320 |  |  |  |  |  |  |  |  |  |  |
|  | Cumulative N | 1,227 | 2,319 | 2,927 | 3,394 | 3,791 | 4,111 |  |  |  |  |  |  |  |  |  |  |
|  | Cumulative % | 14.7 | 27.8 | 35.1 | 40.7 | 45.5 | 49.3 |  |  |  |  |  |  |  |  |  |  |
| 2017 | N | 1,252 | 1,048 | 564 | 487 | 383 |  |  |  |  |  |  |  |  |  |  |  |
|  | Cumulative N | 1,252 | 2,300 | 2,864 | 3,351 | 3,734 |  |  |  |  |  |  |  |  |  |  |  |
|  | Cumulative % | 15.3 | 28.2 | 35.1 | 41.1 | 45.8 |  |  |  |  |  |  |  |  |  |  |  |
| 2018 | N | 1,130 | 1,187 | 636 | 432 |  |  |  |  |  |  |  |  |  |  |  |  |
|  | Cumulative N | 1,130 | 2,317 | 2,953 | 3,385 |  |  |  |  |  |  |  |  |  |  |  |  |
|  | Cumulative % | 13.5 | 27.7 | 35.3 | 40.4 |  |  |  |  |  |  |  |  |  |  |  |  |
| 2019 | N | 1,264 | 1,129 | 562 |  |  |  |  |  |  |  |  |  |  |  |  |  |
|  | Cumulative N | 1,264 | 2,393 | 2,955 |  |  |  |  |  |  |  |  |  |  |  |  |  |
|  | Cumulative % | 14.7 | 27.9 | 34.4 |  |  |  |  |  |  |  |  |  |  |  |  |  |
| 2020 | N | 1,360 | 1,126 |  |  |  |  |  |  |  |  |  |  |  |  |  |  |
|  | Cumulative N | 1,360 | 2,486 |  |  |  |  |  |  |  |  |  |  |  |  |  |  |
|  | Cumulative % | 15.8 | 28.9 |  |  |  |  |  |  |  |  |  |  |  |  |  |  |

**Table E3b.** Number, cumulative number and cumulative incidence of first-time child protection ROSH reports by year of age for Aboriginal children born 2006-2020.

| Birth-year cohort |  | Age |  |  |  |  |  |  |  |  |  |  |  |  |  |  |  |
| --- | --- | --- | --- | --- | --- | --- | --- | --- | --- | --- | --- | --- | --- | --- | --- | --- | --- |
|  |  | Birth | 1 | 2 | 3 | 4 | 5 | 6 | 7 | 8 | 9 | 10 | 11 | 12 | 13 | 14 | 15 |
| 2006 | N | 670 | 1,446 | 611 | 460 | 253 | 150 | 150 | 146 | 156 | 154 | 137 | 116 | 110 | 134 | 134 | 160 |
|  | Cumulative N | 670 | 2,116 | 2,727 | 3,187 | 3,440 | 3,590 | 3,740 | 3,886 | 4,042 | 4,196 | 4,333 | 4,449 | 4,559 | 4,693 | 4,827 | 4,987 |
|  | Cumulative % | 9.2 | 29.0 | 37.4 | 43.7 | 47.1 | 49.2 | 51.2 | 53.2 | 55.4 | 57.5 | 59.4 | 60.9 | 62.5 | 64.3 | 66.1 | 68.3 |
| 2007 | N | 934 | 1,337 | 643 | 355 | 187 | 173 | 143 | 158 | 141 | 142 | 138 | 140 | 140 | 152 | 154 |  |
|  | Cumulative N | 934 | 2,271 | 2,914 | 3,269 | 3,456 | 3,629 | 3,772 | 3,930 | 4,071 | 4,213 | 4,351 | 4,491 | 4,631 | 4,783 | 4,937 |  |
|  | Cumulative % | 12.4 | 30.2 | 38.8 | 43.5 | 46.0 | 48.3 | 50.2 | 52.3 | 54.2 | 56.0 | 57.9 | 59.7 | 61.6 | 63.6 | 65.7 |  |
| 2008 | N | 1,068 | 1,351 | 466 | 234 | 218 | 197 | 174 | 176 | 185 | 152 | 118 | 150 | 141 | 187 |  |  |
|  | Cumulative N | 1,068 | 2,419 | 2,885 | 3,119 | 3,337 | 3,534 | 3,708 | 3,884 | 4,069 | 4,221 | 4,339 | 4,489 | 4,630 | 4,817 |  |  |
|  | Cumulative % | 14.6 | 33.0 | 39.4 | 42.6 | 45.6 | 48.3 | 50.7 | 53.1 | 55.6 | 57.7 | 59.3 | 61.3 | 63.3 | 65.8 |  |  |
| 2009 | N | 1,169 | 1,029 | 336 | 292 | 221 | 254 | 225 | 203 | 203 | 159 | 180 | 187 | 164 |  |  |  |
|  | Cumulative N | 1,169 | 2,198 | 2,534 | 2,826 | 3,047 | 3,301 | 3,526 | 3,729 | 3,932 | 4,091 | 4,271 | 4,458 | 4,622 |  |  |  |
|  | Cumulative % | 16.0 | 30.1 | 34.6 | 38.6 | 41.7 | 45.1 | 48.2 | 51.0 | 53.8 | 55.9 | 58.4 | 61.0 | 63.2 |  |  |  |
| 2010 | N | 962 | 789 | 418 | 377 | 337 | 277 | 250 | 215 | 207 | 209 | 211 | 165 |  |  |  |  |
|  | Cumulative N | 962 | 1,751 | 2,169 | 2,546 | 2,883 | 3,160 | 3,410 | 3,625 | 3,832 | 4,041 | 4,252 | 4,417 |  |  |  |  |
|  | Cumulative % | 12.8 | 23.3 | 28.9 | 33.9 | 38.4 | 42.1 | 45.5 | 48.3 | 51.1 | 53.9 | 56.7 | 58.9 |  |  |  |  |
| 2011 | N | 723 | 801 | 543 | 461 | 353 | 324 | 258 | 230 | 247 | 238 | 208 |  |  |  |  |  |
|  | Cumulative N | 723 | 1,524 | 2,067 | 2,528 | 2,881 | 3,205 | 3,463 | 3,693 | 3,940 | 4,178 | 4,386 |  |  |  |  |  |
|  | Cumulative % | 9.3 | 19.6 | 26.6 | 32.5 | 37.0 | 41.2 | 44.5 | 47.5 | 50.6 | 53.7 | 56.4 |  |  |  |  |  |
| 2012 | N | 767 | 895 | 607 | 453 | 360 | 308 | 281 | 299 | 270 | 216 |  |  |  |  |  |  |
|  | Cumulative N | 767 | 1,662 | 2,269 | 2,722 | 3,082 | 3,390 | 3,671 | 3,970 | 4,240 | 4,456 |  |  |  |  |  |  |
|  | Cumulative % | 9.4 | 20.3 | 27.7 | 33.2 | 37.6 | 41.4 | 44.8 | 48.5 | 51.8 | 54.4 |  |  |  |  |  |  |
| 2013 | N | 864 | 1,006 | 550 | 455 | 369 | 326 | 324 | 312 | 243 |  |  |  |  |  |  |  |
|  | Cumulative N | 864 | 1,870 | 2,420 | 2,875 | 3,244 | 3,570 | 3,894 | 4,206 | 4,449 |  |  |  |  |  |  |  |
|  | Cumulative % | 10.5 | 22.8 | 29.5 | 35.1 | 39.6 | 43.6 | 47.5 | 51.3 | 54.3 |  |  |  |  |  |  |  |
| 2014 | N | 926 | 922 | 612 | 460 | 335 | 323 | 321 | 286 |  |  |  |  |  |  |  |  |
|  | Cumulative N | 926 | 1,848 | 2,460 | 2,920 | 3,255 | 3,578 | 3,899 | 4,185 |  |  |  |  |  |  |  |  |
|  | Cumulative % | 11.1 | 22.2 | 29.6 | 35.1 | 39.1 | 43.0 | 46.9 | 50.3 |  |  |  |  |  |  |  |  |
| 2015 | N | 957 | 968 | 550 | 409 | 372 | 356 | 337 |  |  |  |  |  |  |  |  |  |
|  | Cumulative N | 957 | 1,925 | 2,475 | 2,884 | 3,256 | 3,612 | 3,949 |  |  |  |  |  |  |  |  |  |
|  | Cumulative % | 11.7 | 23.5 | 30.2 | 35.1 | 39.7 | 44.0 | 48.1 |  |  |  |  |  |  |  |  |  |
| 2016 | N | 1,017 | 932 | 564 | 460 | 402 | 315 |  |  |  |  |  |  |  |  |  |  |
|  | Cumulative N | 1,017 | 1,949 | 2,513 | 2,973 | 3,375 | 3,690 |  |  |  |  |  |  |  |  |  |  |
|  | Cumulative % | 12.2 | 23.4 | 30.1 | 35.6 | 40.5 | 44.2 |  |  |  |  |  |  |  |  |  |  |
| 2017 | N | 1,050 | 950 | 554 | 465 | 373 |  |  |  |  |  |  |  |  |  |  |  |
|  | Cumulative N | 1,050 | 2,000 | 2,554 | 3,019 | 3,392 |  |  |  |  |  |  |  |  |  |  |  |
|  | Cumulative % | 12.9 | 24.5 | 31.3 | 37.0 | 41.6 |  |  |  |  |  |  |  |  |  |  |  |
| 2018 | N | 973 | 1,044 | 596 | 445 |  |  |  |  |  |  |  |  |  |  |  |  |
|  | Cumulative N | 973 | 2,017 | 2,613 | 3,058 |  |  |  |  |  |  |  |  |  |  |  |  |
|  | Cumulative % | 11.6 | 24.1 | 31.2 | 36.5 |  |  |  |  |  |  |  |  |  |  |  |  |
| 2019 | N | 1,109 | 1,023 | 542 |  |  |  |  |  |  |  |  |  |  |  |  |  |
|  | Cumulative N | 1,109 | 2,132 | 2,674 |  |  |  |  |  |  |  |  |  |  |  |  |  |
|  | Cumulative % | 12.9 | 24.8 | 31.1 |  |  |  |  |  |  |  |  |  |  |  |  |  |
| 2020 | N | 1,198 | 1,037 |  |  |  |  |  |  |  |  |  |  |  |  |  |  |
|  | Cumulative N | 1,198 | 2,235 |  |  |  |  |  |  |  |  |  |  |  |  |  |  |
|  | Cumulative % | 13.9 | 26.0 |  |  |  |  |  |  |  |  |  |  |  |  |  |  |

**Table E3c.** Number, cumulative number and cumulative incidence of first-time child protection investigations by year of age for Aboriginal children born 2006-2020.

| Birth-year cohort |  | Age |  |  |  |  |  |  |  |  |  |  |  |  |  |  |  |
| --- | --- | --- | --- | --- | --- | --- | --- | --- | --- | --- | --- | --- | --- | --- | --- | --- | --- |
|  |  | Birth | 1 | 2 | 3 | 4 | 5 | 6 | 7 | 8 | 9 | 10 | 11 | 12 | 13 | 14 | 15 |
| 2006 | N | 75 | 573 | 232 | 204 | 158 | 142 | 170 | 160 | 142 | 171 | 146 | 151 | 124 | 133 | 161 | 132 |
|  | Cumulative N | 75 | 648 | 880 | 1,084 | 1,242 | 1,384 | 1,554 | 1,714 | 1,856 | 2,027 | 2,173 | 2,324 | 2,448 | 2,581 | 2,742 | 2,874 |
|  | Cumulative % | 1.0 | 8.9 | 12.1 | 14.8 | 17.0 | 19.0 | 21.3 | 23.5 | 25.4 | 27.8 | 29.8 | 31.8 | 33.5 | 35.4 | 37.6 | 39.4 |
| 2007 | N | 83 | 529 | 241 | 207 | 169 | 200 | 195 | 161 | 146 | 163 | 149 | 141 | 166 | 166 | 132 |  |
|  | Cumulative N | 83 | 612 | 853 | 1,060 | 1,229 | 1,429 | 1,624 | 1,785 | 1,931 | 2,094 | 2,243 | 2,384 | 2,550 | 2,716 | 2,848 |  |
|  | Cumulative % | 1.1 | 8.1 | 11.3 | 14.1 | 16.3 | 19.0 | 21.6 | 23.7 | 25.7 | 27.9 | 29.8 | 31.7 | 33.9 | 36.1 | 37.9 |  |
| 2008 | N | 78 | 631 | 249 | 178 | 219 | 229 | 188 | 173 | 173 | 162 | 156 | 154 | 140 | 141 |  |  |
|  | Cumulative N | 78 | 709 | 958 | 1,136 | 1,355 | 1,584 | 1,772 | 1,945 | 2,118 | 2,280 | 2,436 | 2,590 | 2,730 | 2,871 |  |  |
|  | Cumulative % | 1.1 | 9.7 | 13.1 | 15.5 | 18.5 | 21.6 | 24.2 | 26.6 | 28.9 | 31.1 | 33.3 | 35.4 | 37.3 | 39.2 |  |  |
| 2009 | N | 107 | 536 | 211 | 258 | 222 | 192 | 210 | 176 | 159 | 169 | 165 | 180 | 169 |  |  |  |
|  | Cumulative N | 107 | 643 | 854 | 1,112 | 1,334 | 1,526 | 1,736 | 1,912 | 2,071 | 2,240 | 2,405 | 2,585 | 2,754 |  |  |  |
|  | Cumulative % | 1.5 | 8.8 | 11.7 | 15.2 | 18.2 | 20.9 | 23.7 | 26.1 | 28.3 | 30.6 | 32.9 | 35.3 | 37.7 |  |  |  |
| 2010 | N | 100 | 548 | 298 | 274 | 232 | 221 | 196 | 179 | 159 | 184 | 188 | 144 |  |  |  |  |
|  | Cumulative N | 100 | 648 | 946 | 1,220 | 1,452 | 1,673 | 1,869 | 2,048 | 2,207 | 2,391 | 2,579 | 2,723 |  |  |  |  |
|  | Cumulative % | 1.3 | 8.6 | 12.6 | 16.3 | 19.4 | 22.3 | 24.9 | 27.3 | 29.4 | 31.9 | 34.4 | 36.3 |  |  |  |  |
| 2011 | N | 121 | 623 | 324 | 238 | 236 | 252 | 214 | 175 | 189 | 195 | 140 |  |  |  |  |  |
|  | Cumulative N | 121 | 744 | 1,068 | 1,306 | 1,542 | 1,794 | 2,008 | 2,183 | 2,372 | 2,567 | 2,707 |  |  |  |  |  |
|  | Cumulative % | 1.6 | 9.6 | 13.7 | 16.8 | 19.8 | 23.1 | 25.8 | 28.1 | 30.5 | 33.0 | 34.8 |  |  |  |  |  |
| 2012 | N | 219 | 676 | 333 | 292 | 252 | 250 | 235 | 222 | 194 | 182 |  |  |  |  |  |  |
|  | Cumulative N | 219 | 895 | 1,228 | 1,520 | 1,772 | 2,022 | 2,257 | 2,479 | 2,673 | 2,855 |  |  |  |  |  |  |
|  | Cumulative % | 2.7 | 10.9 | 15.0 | 18.6 | 21.6 | 24.7 | 27.6 | 30.3 | 32.6 | 34.9 |  |  |  |  |  |  |
| 2013 | N | 299 | 707 | 348 | 309 | 264 | 207 | 214 | 254 | 178 |  |  |  |  |  |  |  |
|  | Cumulative N | 299 | 1,006 | 1,354 | 1,663 | 1,927 | 2,134 | 2,348 | 2,602 | 2,780 |  |  |  |  |  |  |  |
|  | Cumulative % | 3.6 | 12.3 | 16.5 | 20.3 | 23.5 | 26.0 | 28.7 | 31.8 | 33.9 |  |  |  |  |  |  |  |
| 2014 | N | 350 | 690 | 396 | 313 | 235 | 226 | 252 | 182 |  |  |  |  |  |  |  |  |
|  | Cumulative N | 350 | 1,040 | 1,436 | 1,749 | 1,984 | 2,210 | 2,462 | 2,644 |  |  |  |  |  |  |  |  |
|  | Cumulative % | 4.2 | 12.5 | 17.3 | 21.0 | 23.8 | 26.6 | 29.6 | 31.8 |  |  |  |  |  |  |  |  |
| 2015 | N | 396 | 736 | 356 | 273 | 261 | 278 | 243 |  |  |  |  |  |  |  |  |  |
|  | Cumulative N | 396 | 1,132 | 1,488 | 1,761 | 2,022 | 2,300 | 2,543 |  |  |  |  |  |  |  |  |  |
|  | Cumulative % | 4.8 | 13.8 | 18.1 | 21.5 | 24.6 | 28.0 | 31.0 |  |  |  |  |  |  |  |  |  |
| 2016 | N | 481 | 712 | 312 | 284 | 326 | 244 |  |  |  |  |  |  |  |  |  |  |
|  | Cumulative N | 481 | 1,193 | 1,505 | 1,789 | 2,115 | 2,359 |  |  |  |  |  |  |  |  |  |  |
|  | Cumulative % | 5.8 | 14.3 | 18.0 | 21.5 | 25.4 | 28.3 |  |  |  |  |  |  |  |  |  |  |
| 2017 | N | 493 | 698 | 381 | 349 | 291 |  |  |  |  |  |  |  |  |  |  |  |
|  | Cumulative N | 493 | 1,191 | 1,572 | 1,921 | 2,212 |  |  |  |  |  |  |  |  |  |  |  |
|  | Cumulative % | 6.0 | 14.6 | 19.3 | 23.6 | 27.1 |  |  |  |  |  |  |  |  |  |  |  |
| 2018 | N | 415 | 770 | 438 | 326 |  |  |  |  |  |  |  |  |  |  |  |  |
|  | Cumulative N | 415 | 1,185 | 1,623 | 1,949 |  |  |  |  |  |  |  |  |  |  |  |  |
|  | Cumulative % | 5.0 | 14.2 | 19.4 | 23.3 |  |  |  |  |  |  |  |  |  |  |  |  |
| 2019 | N | 591 | 804 | 393 |  |  |  |  |  |  |  |  |  |  |  |  |  |
|  | Cumulative N | 591 | 1,395 | 1,788 |  |  |  |  |  |  |  |  |  |  |  |  |  |
|  | Cumulative % | 6.9 | 16.2 | 20.8 |  |  |  |  |  |  |  |  |  |  |  |  |  |
| 2020 | N | 692 | 764 |  |  |  |  |  |  |  |  |  |  |  |  |  |  |
|  | Cumulative N | 692 | 1,456 |  |  |  |  |  |  |  |  |  |  |  |  |  |  |
|  | Cumulative % | 8.0 | 16.9 |  |  |  |  |  |  |  |  |  |  |  |  |  |  |

**Table E3d.**
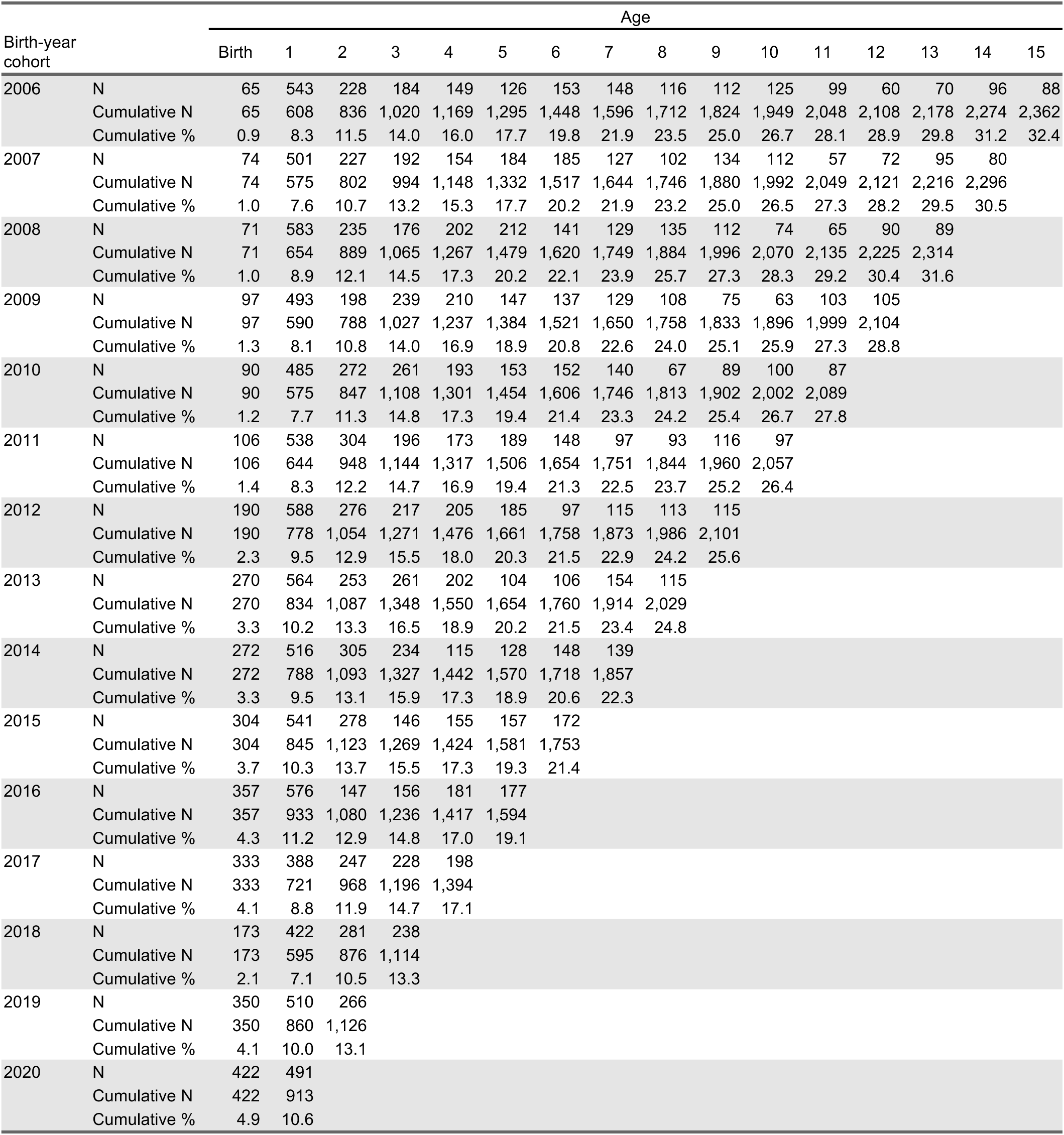
Number, cumulative number and cumulative incidence of first-time child protection service-defined substantiations by year of age for Aboriginal children born 2006- 2020.

| Birth-year cohort |  | Age |  |  |  |  |  |  |  |  |  |  |  |  |  |  |  |
| --- | --- | --- | --- | --- | --- | --- | --- | --- | --- | --- | --- | --- | --- | --- | --- | --- | --- |
|  |  | Birth | 1 | 2 | 3 | 4 | 5 | 6 | 7 | 8 | 9 | 10 | 11 | 12 | 13 | 14 | 15 |
| 2006 | N | 65 | 543 | 228 | 184 | 149 | 126 | 153 | 148 | 116 | 112 | 125 | 99 | 60 | 70 | 96 | 88 |
|  | Cumulative N | 65 | 608 | 836 | 1,020 | 1,169 | 1,295 | 1,448 | 1,596 | 1,712 | 1,824 | 1,949 | 2,048 | 2,108 | 2,178 | 2,274 | 2,362 |
|  | Cumulative % | 0.9 | 8.3 | 11.5 | 14.0 | 16.0 | 17.7 | 19.8 | 21.9 | 23.5 | 25.0 | 26.7 | 28.1 | 28.9 | 29.8 | 31.2 | 32.4 |
| 2007 | N | 74 | 501 | 227 | 192 | 154 | 184 | 185 | 127 | 102 | 134 | 112 | 57 | 72 | 95 | 80 |  |
|  | Cumulative N | 74 | 575 | 802 | 994 | 1,148 | 1,332 | 1,517 | 1,644 | 1,746 | 1,880 | 1,992 | 2,049 | 2,121 | 2,216 | 2,296 |  |
|  | Cumulative % | 1.0 | 7.6 | 10.7 | 13.2 | 15.3 | 17.7 | 20.2 | 21.9 | 23.2 | 25.0 | 26.5 | 27.3 | 28.2 | 29.5 | 30.5 |  |
| 2008 | N | 71 | 583 | 235 | 176 | 202 | 212 | 141 | 129 | 135 | 112 | 74 | 65 | 90 | 89 |  |  |
|  | Cumulative N | 71 | 654 | 889 | 1,065 | 1,267 | 1,479 | 1,620 | 1,749 | 1,884 | 1,996 | 2,070 | 2,135 | 2,225 | 2,314 |  |  |
|  | Cumulative % | 1.0 | 8.9 | 12.1 | 14.5 | 17.3 | 20.2 | 22.1 | 23.9 | 25.7 | 27.3 | 28.3 | 29.2 | 30.4 | 31.6 |  |  |
| 2009 | N | 97 | 493 | 198 | 239 | 210 | 147 | 137 | 129 | 108 | 75 | 63 | 103 | 105 |  |  |  |
|  | Cumulative N | 97 | 590 | 788 | 1,027 | 1,237 | 1,384 | 1,521 | 1,650 | 1,758 | 1,833 | 1,896 | 1,999 | 2,104 |  |  |  |
|  | Cumulative % | 1.3 | 8.1 | 10.8 | 14.0 | 16.9 | 18.9 | 20.8 | 22.6 | 24.0 | 25.1 | 25.9 | 27.3 | 28.8 |  |  |  |
| 2010 | N | 90 | 485 | 272 | 261 | 193 | 153 | 152 | 140 | 67 | 89 | 100 | 87 |  |  |  |  |
|  | Cumulative N | 90 | 575 | 847 | 1,108 | 1,301 | 1,454 | 1,606 | 1,746 | 1,813 | 1,902 | 2,002 | 2,089 |  |  |  |  |
|  | Cumulative % | 1.2 | 7.7 | 11.3 | 14.8 | 17.3 | 19.4 | 21.4 | 23.3 | 24.2 | 25.4 | 26.7 | 27.8 |  |  |  |  |
| 2011 | N | 106 | 538 | 304 | 196 | 173 | 189 | 148 | 97 | 93 | 116 | 97 |  |  |  |  |  |
|  | Cumulative N | 106 | 644 | 948 | 1,144 | 1,317 | 1,506 | 1,654 | 1,751 | 1,844 | 1,960 | 2,057 |  |  |  |  |  |
|  | Cumulative % | 1.4 | 8.3 | 12.2 | 14.7 | 16.9 | 19.4 | 21.3 | 22.5 | 23.7 | 25.2 | 26.4 |  |  |  |  |  |
| 2012 | N | 190 | 588 | 276 | 217 | 205 | 185 | 97 | 115 | 113 | 115 |  |  |  |  |  |  |
|  | Cumulative N | 190 | 778 | 1,054 | 1,271 | 1,476 | 1,661 | 1,758 | 1,873 | 1,986 | 2,101 |  |  |  |  |  |  |
|  | Cumulative % | 2.3 | 9.5 | 12.9 | 15.5 | 18.0 | 20.3 | 21.5 | 22.9 | 24.2 | 25.6 |  |  |  |  |  |  |
| 2013 | N | 270 | 564 | 253 | 261 | 202 | 104 | 106 | 154 | 115 |  |  |  |  |  |  |  |
|  | Cumulative N | 270 | 834 | 1,087 | 1,348 | 1,550 | 1,654 | 1,760 | 1,914 | 2,029 |  |  |  |  |  |  |  |
|  | Cumulative % | 3.3 | 10.2 | 13.3 | 16.5 | 18.9 | 20.2 | 21.5 | 23.4 | 24.8 |  |  |  |  |  |  |  |
| 2014 | N | 272 | 516 | 305 | 234 | 115 | 128 | 148 | 139 |  |  |  |  |  |  |  |  |
|  | Cumulative N | 272 | 788 | 1,093 | 1,327 | 1,442 | 1,570 | 1,718 | 1,857 |  |  |  |  |  |  |  |  |
|  | Cumulative % | 3.3 | 9.5 | 13.1 | 15.9 | 17.3 | 18.9 | 20.6 | 22.3 |  |  |  |  |  |  |  |  |
| 2015 | N | 304 | 541 | 278 | 146 | 155 | 157 | 172 |  |  |  |  |  |  |  |  |  |
|  | Cumulative N | 304 | 845 | 1,123 | 1,269 | 1,424 | 1,581 | 1,753 |  |  |  |  |  |  |  |  |  |
|  | Cumulative % | 3.7 | 10.3 | 13.7 | 15.5 | 17.3 | 19.3 | 21.4 |  |  |  |  |  |  |  |  |  |
| 2016 | N | 357 | 576 | 147 | 156 | 181 | 177 |  |  |  |  |  |  |  |  |  |  |
|  | Cumulative N | 357 | 933 | 1,080 | 1,236 | 1,417 | 1,594 |  |  |  |  |  |  |  |  |  |  |
|  | Cumulative % | 4.3 | 11.2 | 12.9 | 14.8 | 17.0 | 19.1 |  |  |  |  |  |  |  |  |  |  |
| 2017 | N | 333 | 388 | 247 | 228 | 198 |  |  |  |  |  |  |  |  |  |  |  |
|  | Cumulative N | 333 | 721 | 968 | 1,196 | 1,394 |  |  |  |  |  |  |  |  |  |  |  |
|  | Cumulative % | 4.1 | 8.8 | 11.9 | 14.7 | 17.1 |  |  |  |  |  |  |  |  |  |  |  |
| 2018 | N | 173 | 422 | 281 | 238 |  |  |  |  |  |  |  |  |  |  |  |  |
|  | Cumulative N | 173 | 595 | 876 | 1,114 |  |  |  |  |  |  |  |  |  |  |  |  |
|  | Cumulative % | 2.1 | 7.1 | 10.5 | 13.3 |  |  |  |  |  |  |  |  |  |  |  |  |
| 2019 | N | 350 | 510 | 266 |  |  |  |  |  |  |  |  |  |  |  |  |  |
|  | Cumulative N | 350 | 860 | 1,126 |  |  |  |  |  |  |  |  |  |  |  |  |  |
|  | Cumulative % | 4.1 | 10.0 | 13.1 |  |  |  |  |  |  |  |  |  |  |  |  |  |
| 2020 | N | 422 | 491 |  |  |  |  |  |  |  |  |  |  |  |  |  |  |
|  | Cumulative N | 422 | 913 |  |  |  |  |  |  |  |  |  |  |  |  |  |  |
|  | Cumulative % | 4.9 | 10.6 |  |  |  |  |  |  |  |  |  |  |  |  |  |  |

**Table E3e.** Number, cumulative number and cumulative incidence of first-time OOHC placements by year of age for Aboriginal children born 2006-2020.

| Birth-year cohort |  | Age |  |  |  |  |  |  |  |  |  |  |  |  |  |  |  |
| --- | --- | --- | --- | --- | --- | --- | --- | --- | --- | --- | --- | --- | --- | --- | --- | --- | --- |
|  |  | Birth | 1 | 2 | 3 | 4 | 5 | 6 | 7 | 8 | 9 | 10 | 11 | 12 | 13 | 14 | 15 |
| 2006 | N | 0 | 219 | 129 | 113 | 92 | 82 | 67 | 50 | 58 | 59 | 67 | 40 | 25 | 19 | 18 | 16 |
|  | Cumulative N | 0 | 219 | 348 | 461 | 553 | 635 | 702 | 752 | 810 | 869 | 936 | 976 | 1,001 | 1,020 | 1,038 | 1,054 |
|  | Cumulative % | 0.0 | 3.0 | 4.8 | 6.3 | 7.6 | 8.7 | 9.6 | 10.3 | 11.1 | 11.9 | 12.8 | 13.4 | 13.7 | 14.0 | 14.2 | 14.4 |
| 2007 | N | 0 | 248 | 121 | 97 | 81 | 60 | 64 | 69 | 77 | 71 | 41 | 27 | 28 | 23 | 17 |  |
|  | Cumulative N | 0 | 248 | 369 | 466 | 547 | 607 | 671 | 740 | 817 | 888 | 929 | 956 | 984 | 1,007 | 1,024 |  |
|  | Cumulative % | 0.0 | 3.3 | 4.9 | 6.2 | 7.3 | 8.1 | 8.9 | 9.8 | 10.9 | 11.8 | 12.4 | 12.7 | 13.1 | 13.4 | 13.6 |  |
| 2008 | N | 0 | 299 | 112 | 82 | 68 | 66 | 82 | 60 | 81 | 43 | 25 | 29 | 21 | 25 |  |  |
|  | Cumulative N | 0 | 299 | 411 | 493 | 561 | 627 | 709 | 769 | 850 | 893 | 918 | 947 | 968 | 993 |  |  |
|  | Cumulative % | 0.0 | 4.1 | 5.6 | 6.7 | 7.7 | 8.6 | 9.7 | 10.5 | 11.6 | 12.2 | 12.5 | 12.9 | 13.2 | 13.6 |  |  |
| 2009 | N | 0 | 282 | 105 | 90 | 85 | 67 | 74 | 68 | 42 | 31 | 20 | 27 | 23 |  |  |  |
|  | Cumulative N | 0 | 282 | 387 | 477 | 562 | 629 | 703 | 771 | 813 | 844 | 864 | 891 | 914 |  |  |  |
|  | Cumulative % | 0.0 | 3.9 | 5.3 | 6.5 | 7.7 | 8.6 | 9.6 | 10.5 | 11.1 | 11.5 | 11.8 | 12.2 | 12.5 |  |  |  |
| 2010 | N | 0 | 300 | 102 | 98 | 69 | 75 | 67 | 52 | 37 | 35 | 25 | 31 |  |  |  |  |
|  | Cumulative N | 0 | 300 | 402 | 500 | 569 | 644 | 711 | 763 | 800 | 835 | 860 | 891 |  |  |  |  |
|  | Cumulative % | 0.0 | 4.0 | 5.4 | 6.7 | 7.6 | 8.6 | 9.5 | 10.2 | 10.7 | 11.1 | 11.5 | 11.9 |  |  |  |  |
| 2011 | N | 0 | 291 | 99 | 86 | 84 | 108 | 59 | 26 | 30 | 29 | 26 |  |  |  |  |  |
|  | Cumulative N | 0 | 291 | 390 | 476 | 560 | 668 | 727 | 753 | 783 | 812 | 838 |  |  |  |  |  |
|  | Cumulative % | 0.0 | 3.7 | 5.0 | 6.1 | 7.2 | 8.6 | 9.3 | 9.7 | 10.1 | 10.4 | 10.8 |  |  |  |  |  |
| 2012 | N | 0 | 277 | 125 | 107 | 99 | 65 | 45 | 37 | 38 | 31 |  |  |  |  |  |  |
|  | Cumulative N | 0 | 277 | 402 | 509 | 608 | 673 | 718 | 755 | 793 | 824 |  |  |  |  |  |  |
|  | Cumulative % | 0.0 | 3.4 | 4.9 | 6.2 | 7.4 | 8.2 | 8.8 | 9.2 | 9.7 | 10.1 |  |  |  |  |  |  |
| 2013 | N | 0 | 286 | 142 | 120 | 88 | 54 | 51 | 52 | 40 |  |  |  |  |  |  |  |
|  | Cumulative N | 0 | 286 | 428 | 548 | 636 | 690 | 741 | 793 | 833 |  |  |  |  |  |  |  |
|  | Cumulative % | 0.0 | 3.5 | 5.2 | 6.7 | 7.8 | 8.4 | 9.0 | 9.7 | 10.2 |  |  |  |  |  |  |  |
| 2014 | N | 0 | 323 | 134 | 111 | 46 | 38 | 52 | 29 |  |  |  |  |  |  |  |  |
|  | Cumulative N | 0 | 323 | 457 | 568 | 614 | 652 | 704 | 733 |  |  |  |  |  |  |  |  |
|  | Cumulative % | 0.0 | 3.9 | 5.5 | 6.8 | 7.4 | 7.8 | 8.5 | 8.8 |  |  |  |  |  |  |  |  |
| 2015 | N | 0 | 343 | 114 | 55 | 52 | 45 | 44 |  |  |  |  |  |  |  |  |  |
|  | Cumulative N | 0 | 343 | 457 | 512 | 564 | 609 | 653 |  |  |  |  |  |  |  |  |  |
|  | Cumulative % | 0.0 | 4.2 | 5.6 | 6.2 | 6.9 | 7.4 | 8.0 |  |  |  |  |  |  |  |  |  |
| 2016 | N | 0 | 335 | 65 | 67 | 63 | 55 |  |  |  |  |  |  |  |  |  |  |
|  | Cumulative N | 0 | 335 | 400 | 467 | 530 | 585 |  |  |  |  |  |  |  |  |  |  |
|  | Cumulative % | 0.0 | 4.0 | 4.8 | 5.6 | 6.4 | 7.0 |  |  |  |  |  |  |  |  |  |  |
| 2017 | N | 0 | 232 | 92 | 67 | 66 |  |  |  |  |  |  |  |  |  |  |  |
|  | Cumulative N | 0 | 232 | 324 | 391 | 457 |  |  |  |  |  |  |  |  |  |  |  |
|  | Cumulative % | 0.0 | 2.8 | 4.0 | 4.8 | 5.6 |  |  |  |  |  |  |  |  |  |  |  |
| 2018 | N | 0 | 229 | 76 | 72 |  |  |  |  |  |  |  |  |  |  |  |  |
|  | Cumulative N | 0 | 229 | 305 | 377 |  |  |  |  |  |  |  |  |  |  |  |  |
|  | Cumulative % | 0.0 | 2.7 | 3.6 | 4.5 |  |  |  |  |  |  |  |  |  |  |  |  |
| 2019 | N | 0 | 223 | 96 |  |  |  |  |  |  |  |  |  |  |  |  |  |
|  | Cumulative N | 0 | 223 | 319 |  |  |  |  |  |  |  |  |  |  |  |  |  |
|  | Cumulative % | 0.0 | 2.6 | 3.7 |  |  |  |  |  |  |  |  |  |  |  |  |  |
| 2020 | N | 0 | 239 |  |  |  |  |  |  |  |  |  |  |  |  |  |  |
|  | Cumulative N | 0 | 239 |  |  |  |  |  |  |  |  |  |  |  |  |  |  |
|  | Cumulative % | 0.0 | 2.8 |  |  |  |  |  |  |  |  |  |  |  |  |  |  |

**Table E4a.** Number, cumulative number and cumulative incidence of first-time child protection reports by year of age for non-Aboriginal children born 2006-2020.

| Birth-year cohort |  | Age |  |  |  |  |  |  |  |  |  |  |  |  |  |  |  |
| --- | --- | --- | --- | --- | --- | --- | --- | --- | --- | --- | --- | --- | --- | --- | --- | --- | --- |
|  |  | Birth | 1 | 2 | 3 | 4 | 5 | 6 | 7 | 8 | 9 | 10 | 11 | 12 | 13 | 14 | 15 |
| 2006 | N | 1,484 | 4,635 | 2,971 | 2,829 | 2,198 | 1,551 | 1,627 | 1,579 | 1,561 | 1,534 | 1,563 | 1,635 | 1,643 | 1,747 | 2,077 | 2,424 |
|  | Cumulative N | 1,484 | 6,119 | 9,090 | 11,919 | 14,117 | 15,668 | 17,295 | 18,874 | 20,435 | 21,969 | 23,532 | 25,167 | 26,810 | 28,557 | 30,634 | 33,058 |
|  | Cumulative % | 1.5 | 6.2 | 9.2 | 12.1 | 14.3 | 15.8 | 17.5 | 19.1 | 20.7 | 22.2 | 23.8 | 25.4 | 27.1 | 28.9 | 31.0 | 33.4 |
| 2007 | N | 2,113 | 4,938 | 3,025 | 2,341 | 1,718 | 1,741 | 1,635 | 1,786 | 1,652 | 1,739 | 1,668 | 1,764 | 1,665 | 2,017 | 2,409 |  |
|  | Cumulative N | 2,113 | 7,051 | 10,076 | 12,417 | 14,135 | 15,876 | 17,511 | 19,297 | 20,949 | 22,688 | 24,356 | 26,120 | 27,785 | 29,802 | 32,211 |  |
|  | Cumulative % | 2.1 | 7.0 | 10.0 | 12.3 | 14.0 | 15.7 | 17.3 | 19.1 | 20.7 | 22.4 | 24.1 | 25.8 | 27.5 | 29.5 | 31.9 |  |
| 2008 | N | 2,442 | 4,818 | 2,350 | 1,826 | 1,871 | 1,822 | 1,877 | 1,828 | 1,738 | 1,851 | 1,876 | 1,810 | 1,971 | 2,242 |  |  |
|  | Cumulative N | 2,442 | 7,260 | 9,610 | 11,436 | 13,307 | 15,129 | 17,006 | 18,834 | 20,572 | 22,423 | 24,299 | 26,109 | 28,080 | 30,322 |  |  |
|  | Cumulative % | 2.4 | 7.2 | 9.5 | 11.3 | 13.2 | 15.0 | 16.8 | 18.6 | 20.3 | 22.2 | 24.0 | 25.8 | 27.8 | 30.0 |  |  |
| 2009 | N | 2,569 | 3,842 | 1,898 | 1,904 | 1,884 | 1,930 | 1,980 | 1,881 | 1,874 | 1,999 | 1,793 | 2,064 | 2,036 |  |  |  |
|  | Cumulative N | 2,569 | 6,411 | 8,309 | 10,213 | 12,097 | 14,027 | 16,007 | 17,888 | 19,762 | 21,761 | 23,554 | 25,618 | 27,654 |  |  |  |
|  | Cumulative % | 2.6 | 6.4 | 8.3 | 10.2 | 12.1 | 14.0 | 16.0 | 17.9 | 19.8 | 21.8 | 23.5 | 25.6 | 27.6 |  |  |  |
| 2010 | N | 1,969 | 2,861 | 2,120 | 2,017 | 2,130 | 2,085 | 1,948 | 2,065 | 2,110 | 1,901 | 2,144 | 2,343 |  |  |  |  |
|  | Cumulative N | 1,969 | 4,830 | 6,950 | 8,967 | 11,097 | 13,182 | 15,130 | 17,195 | 19,305 | 21,206 | 23,350 | 25,693 |  |  |  |  |
|  | Cumulative % | 2.0 | 4.9 | 7.0 | 9.0 | 11.2 | 13.3 | 15.2 | 17.3 | 19.4 | 21.3 | 23.5 | 25.8 |  |  |  |  |
| 2011 | N | 1,690 | 2,956 | 2,190 | 2,160 | 2,123 | 2,014 | 2,170 | 2,100 | 2,033 | 2,160 | 2,157 |  |  |  |  |  |
|  | Cumulative N | 1,690 | 4,646 | 6,836 | 8,996 | 11,119 | 13,133 | 15,303 | 17,403 | 19,436 | 21,596 | 23,753 |  |  |  |  |  |
|  | Cumulative % | 1.7 | 4.7 | 6.9 | 9.1 | 11.3 | 13.3 | 15.5 | 17.6 | 19.7 | 21.9 | 24.1 |  |  |  |  |  |
| 2012 | N | 1,764 | 3,076 | 2,253 | 2,146 | 2,091 | 2,146 | 2,285 | 2,183 | 2,208 | 2,284 |  |  |  |  |  |  |
|  | Cumulative N | 1,764 | 4,840 | 7,093 | 9,239 | 11,330 | 13,476 | 15,761 | 17,944 | 20,152 | 22,436 |  |  |  |  |  |  |
|  | Cumulative % | 1.7 | 4.8 | 7.0 | 9.1 | 11.2 | 13.3 | 15.5 | 17.7 | 19.9 | 22.1 |  |  |  |  |  |  |
| 2013 | N | 1,836 | 3,009 | 2,176 | 2,086 | 2,139 | 2,242 | 2,205 | 2,239 | 2,297 |  |  |  |  |  |  |  |
|  | Cumulative N | 1,836 | 4,845 | 7,021 | 9,107 | 11,246 | 13,488 | 15,693 | 17,932 | 20,229 |  |  |  |  |  |  |  |
|  | Cumulative % | 1.9 | 5.0 | 7.2 | 9.3 | 11.5 | 13.8 | 16.1 | 18.4 | 20.7 |  |  |  |  |  |  |  |
| 2014 | N | 1,904 | 2,990 | 2,133 | 2,051 | 2,113 | 2,057 | 2,376 | 2,402 |  |  |  |  |  |  |  |  |
|  | Cumulative N | 1,904 | 4,894 | 7,027 | 9,078 | 11,191 | 13,248 | 15,624 | 18,026 |  |  |  |  |  |  |  |  |
|  | Cumulative % | 1.9 | 5.0 | 7.2 | 9.3 | 11.5 | 13.6 | 16.0 | 18.5 |  |  |  |  |  |  |  |  |
| 2015 | N | 1,819 | 2,977 | 2,156 | 2,080 | 1,998 | 2,224 | 2,449 |  |  |  |  |  |  |  |  |  |
|  | Cumulative N | 1,819 | 4,796 | 6,952 | 9,032 | 11,030 | 13,254 | 15,703 |  |  |  |  |  |  |  |  |  |
|  | Cumulative % | 1.9 | 5.0 | 7.3 | 9.5 | 11.6 | 13.9 | 16.5 |  |  |  |  |  |  |  |  |  |
| 2016 | N | 1,991 | 2,889 | 2,123 | 2,047 | 2,220 | 2,344 |  |  |  |  |  |  |  |  |  |  |
|  | Cumulative N | 1,991 | 4,880 | 7,003 | 9,050 | 11,270 | 13,614 |  |  |  |  |  |  |  |  |  |  |
|  | Cumulative % | 2.1 | 5.0 | 7.2 | 9.4 | 11.6 | 14.1 |  |  |  |  |  |  |  |  |  |  |
| 2017 | N | 2,007 | 2,882 | 2,109 | 2,085 | 2,213 |  |  |  |  |  |  |  |  |  |  |  |
|  | Cumulative N | 2,007 | 4,889 | 6,998 | 9,083 | 11,296 |  |  |  |  |  |  |  |  |  |  |  |
|  | Cumulative % | 2.1 | 5.2 | 7.5 | 9.7 | 12.1 |  |  |  |  |  |  |  |  |  |  |  |
| 2018 | N | 1,869 | 3,132 | 2,268 | 2,131 |  |  |  |  |  |  |  |  |  |  |  |  |
|  | Cumulative N | 1,869 | 5,001 | 7,269 | 9,400 |  |  |  |  |  |  |  |  |  |  |  |  |
|  | Cumulative % | 2.0 | 5.3 | 7.7 | 9.9 |  |  |  |  |  |  |  |  |  |  |  |  |
| 2019 | N | 2,010 | 2,938 | 2,158 |  |  |  |  |  |  |  |  |  |  |  |  |  |
|  | Cumulative N | 2,010 | 4,948 | 7,106 |  |  |  |  |  |  |  |  |  |  |  |  |  |
|  | Cumulative % | 2.2 | 5.4 | 7.7 |  |  |  |  |  |  |  |  |  |  |  |  |  |
| 2020 | N | 1,951 | 2,895 |  |  |  |  |  |  |  |  |  |  |  |  |  |  |
|  | Cumulative N | 1,951 | 4,846 |  |  |  |  |  |  |  |  |  |  |  |  |  |  |
|  | Cumulative % | 2.2 | 5.5 |  |  |  |  |  |  |  |  |  |  |  |  |  |  |

**Table E4b.** Number, cumulative number and cumulative incidence of first-time child protection ROSH reports by year of age for non-Aboriginal children born 2006-2020.

| Birth-year cohort |  | Age |  |  |  |  |  |  |  |  |  |  |  |  |  |  |  |
| --- | --- | --- | --- | --- | --- | --- | --- | --- | --- | --- | --- | --- | --- | --- | --- | --- | --- |
|  |  | Birth | 1 | 2 | 3 | 4 | 5 | 6 | 7 | 8 | 9 | 10 | 11 | 12 | 13 | 14 | 15 |
| 2006 | N | 1,258 | 3,933 | 2,460 | 2,451 | 1,785 | 1,009 | 984 | 1,057 | 1,134 | 1,090 | 1,117 | 1,274 | 1,353 | 1,447 | 1,747 | 2,098 |
|  | Cumulative N | 1,258 | 5,191 | 7,651 | 10,102 | 11,887 | 12,896 | 13,880 | 14,937 | 16,071 | 17,161 | 18,278 | 19,552 | 20,905 | 22,352 | 24,099 | 26,197 |
|  | Cumulative % | 1.3 | 5.2 | 7.7 | 10.2 | 12.0 | 13.0 | 14.0 | 15.1 | 16.2 | 17.4 | 18.5 | 19.8 | 21.1 | 22.6 | 24.4 | 26.5 |
| 2007 | N | 1,969 | 4,109 | 2,608 | 1,881 | 1,077 | 1,062 | 1,064 | 1,237 | 1,192 | 1,289 | 1,339 | 1,397 | 1,442 | 1,704 | 2,095 |  |
|  | Cumulative N | 1,969 | 6,078 | 8,686 | 10,567 | 11,644 | 12,706 | 13,770 | 15,007 | 16,199 | 17,488 | 18,827 | 20,224 | 21,666 | 23,370 | 25,465 |  |
|  | Cumulative % | 1.9 | 6.0 | 8.6 | 10.5 | 11.5 | 12.6 | 13.6 | 14.8 | 16.0 | 17.3 | 18.6 | 20.0 | 21.4 | 23.1 | 25.2 |  |
| 2008 | N | 2,275 | 4,156 | 1,877 | 1,125 | 1,138 | 1,164 | 1,330 | 1,298 | 1,304 | 1,452 | 1,519 | 1,546 | 1,671 | 1,932 |  |  |
|  | Cumulative N | 2,275 | 6,431 | 8,308 | 9,433 | 10,571 | 11,735 | 13,065 | 14,363 | 15,667 | 17,119 | 18,638 | 20,184 | 21,855 | 23,787 |  |  |
|  | Cumulative % | 2.2 | 6.4 | 8.2 | 9.3 | 10.5 | 11.6 | 12.9 | 14.2 | 15.5 | 16.9 | 18.4 | 20.0 | 21.6 | 23.5 |  |  |
| 2009 | N | 2,419 | 3,118 | 1,215 | 1,161 | 1,203 | 1,334 | 1,383 | 1,398 | 1,470 | 1,636 | 1,529 | 1,790 | 1,746 |  |  |  |
|  | Cumulative N | 2,419 | 5,537 | 6,752 | 7,913 | 9,116 | 10,450 | 11,833 | 13,231 | 14,701 | 16,337 | 17,866 | 19,656 | 21,402 |  |  |  |
|  | Cumulative % | 2.4 | 5.5 | 6.7 | 7.9 | 9.1 | 10.4 | 11.8 | 13.2 | 14.7 | 16.3 | 17.9 | 19.6 | 21.4 |  |  |  |
| 2010 | N | 1,624 | 1,854 | 1,293 | 1,300 | 1,499 | 1,474 | 1,472 | 1,631 | 1,694 | 1,621 | 1,900 | 2,000 |  |  |  |  |
|  | Cumulative N | 1,624 | 3,478 | 4,771 | 6,071 | 7,570 | 9,044 | 10,516 | 12,147 | 13,841 | 15,462 | 17,362 | 19,362 |  |  |  |  |
|  | Cumulative % | 1.6 | 3.5 | 4.8 | 6.1 | 7.6 | 9.1 | 10.6 | 12.2 | 13.9 | 15.6 | 17.5 | 19.5 |  |  |  |  |
| 2011 | N | 1,088 | 1,893 | 1,497 | 1,508 | 1,474 | 1,517 | 1,666 | 1,716 | 1,787 | 1,806 | 1,847 |  |  |  |  |  |
|  | Cumulative N | 1,088 | 2,981 | 4,478 | 5,986 | 7,460 | 8,977 | 10,643 | 12,359 | 14,146 | 15,952 | 17,799 |  |  |  |  |  |
|  | Cumulative % | 1.1 | 3.0 | 4.5 | 6.1 | 7.6 | 9.1 | 10.8 | 12.5 | 14.3 | 16.2 | 18.0 |  |  |  |  |  |
| 2012 | N | 1,126 | 1,936 | 1,619 | 1,534 | 1,562 | 1,652 | 1,801 | 1,822 | 1,915 | 1,966 |  |  |  |  |  |  |
|  | Cumulative N | 1,126 | 3,062 | 4,681 | 6,215 | 7,777 | 9,429 | 11,230 | 13,052 | 14,967 | 16,933 |  |  |  |  |  |  |
|  | Cumulative % | 1.1 | 3.0 | 4.6 | 6.1 | 7.7 | 9.3 | 11.1 | 12.9 | 14.7 | 16.7 |  |  |  |  |  |  |
| 2013 | N | 1,174 | 2,118 | 1,553 | 1,496 | 1,605 | 1,730 | 1,875 | 1,921 | 1,951 |  |  |  |  |  |  |  |
|  | Cumulative N | 1,174 | 3,292 | 4,845 | 6,341 | 7,946 | 9,676 | 11,551 | 13,472 | 15,423 |  |  |  |  |  |  |  |
|  | Cumulative % | 1.2 | 3.4 | 5.0 | 6.5 | 8.1 | 9.9 | 11.8 | 13.8 | 15.8 |  |  |  |  |  |  |  |
| 2014 | N | 1,298 | 2,029 | 1,551 | 1,593 | 1,622 | 1,694 | 2,009 | 2,013 |  |  |  |  |  |  |  |  |
|  | Cumulative N | 1,298 | 3,327 | 4,878 | 6,471 | 8,093 | 9,787 | 11,796 | 13,809 |  |  |  |  |  |  |  |  |
|  | Cumulative % | 1.3 | 3.4 | 5.0 | 6.6 | 8.3 | 10.0 | 12.1 | 14.1 |  |  |  |  |  |  |  |  |
| 2015 | N | 1,247 | 2,172 | 1,640 | 1,580 | 1,652 | 1,864 | 2,069 |  |  |  |  |  |  |  |  |  |
|  | Cumulative N | 1,247 | 3,419 | 5,059 | 6,639 | 8,291 | 10,155 | 12,224 |  |  |  |  |  |  |  |  |  |
|  | Cumulative % | 1.3 | 3.6 | 5.3 | 7.0 | 8.7 | 10.6 | 12.8 |  |  |  |  |  |  |  |  |  |
| 2016 | N | 1,461 | 2,130 | 1,673 | 1,670 | 1,852 | 1,968 |  |  |  |  |  |  |  |  |  |  |
|  | Cumulative N | 1,461 | 3,591 | 5,264 | 6,934 | 8,786 | 10,754 |  |  |  |  |  |  |  |  |  |  |
|  | Cumulative % | 1.5 | 3.7 | 5.4 | 7.2 | 9.1 | 11.1 |  |  |  |  |  |  |  |  |  |  |
| 2017 | N | 1,551 | 2,204 | 1,723 | 1,690 | 1,883 |  |  |  |  |  |  |  |  |  |  |  |
|  | Cumulative N | 1,551 | 3,755 | 5,478 | 7,168 | 9,051 |  |  |  |  |  |  |  |  |  |  |  |
|  | Cumulative % | 1.7 | 4.0 | 5.9 | 7.7 | 9.7 |  |  |  |  |  |  |  |  |  |  |  |
| 2018 | N | 1,461 | 2,447 | 1,882 | 1,773 |  |  |  |  |  |  |  |  |  |  |  |  |
|  | Cumulative N | 1,461 | 3,908 | 5,790 | 7,563 |  |  |  |  |  |  |  |  |  |  |  |  |
|  | Cumulative % | 1.5 | 4.1 | 6.1 | 8.0 |  |  |  |  |  |  |  |  |  |  |  |  |
| 2019 | N | 1,604 | 2,409 | 1,779 |  |  |  |  |  |  |  |  |  |  |  |  |  |
|  | Cumulative N | 1,604 | 4,013 | 5,792 |  |  |  |  |  |  |  |  |  |  |  |  |  |
|  | Cumulative % | 1.7 | 4.4 | 6.3 |  |  |  |  |  |  |  |  |  |  |  |  |  |
| 2020 | N | 1,568 | 2,377 |  |  |  |  |  |  |  |  |  |  |  |  |  |  |
|  | Cumulative N | 1,568 | 3,945 |  |  |  |  |  |  |  |  |  |  |  |  |  |  |
|  | Cumulative % | 1.8 | 4.5 |  |  |  |  |  |  |  |  |  |  |  |  |  |  |

**Table E4c.** Number, cumulative number and cumulative incidence of first-time child protections investigation by year of age for non-Aboriginal children born 2006-2020.

| Birth-year cohort |  | Age |  |  |  |  |  |  |  |  |  |  |  |  |  |  |  |
| --- | --- | --- | --- | --- | --- | --- | --- | --- | --- | --- | --- | --- | --- | --- | --- | --- | --- |
|  |  | Birth | 1 | 2 | 3 | 4 | 5 | 6 | 7 | 8 | 9 | 10 | 11 | 12 | 13 | 14 | 15 |
| 2006 | N | 101 | 905 | 417 | 408 | 371 | 317 | 408 | 383 | 407 | 435 | 475 | 653 | 593 | 649 | 633 | 793 |
|  | Cumulative N | 101 | 1,006 | 1,423 | 1,831 | 2,202 | 2,519 | 2,927 | 3,310 | 3,717 | 4,152 | 4,627 | 5,280 | 5,873 | 6,522 | 7,155 | 7,948 |
|  | Cumulative % | 0.1 | 1.0 | 1.4 | 1.9 | 2.2 | 2.5 | 3.0 | 3.3 | 3.8 | 4.2 | 4.7 | 5.3 | 5.9 | 6.6 | 7.2 | 8.0 |
| 2007 | N | 115 | 959 | 458 | 345 | 326 | 510 | 460 | 444 | 437 | 574 | 636 | 589 | 625 | 617 | 815 |  |
|  | Cumulative N | 115 | 1,074 | 1,532 | 1,877 | 2,203 | 2,713 | 3,173 | 3,617 | 4,054 | 4,628 | 5,264 | 5,853 | 6,478 | 7,095 | 7,910 |  |
|  | Cumulative % | 0.1 | 1.1 | 1.5 | 1.9 | 2.2 | 2.7 | 3.1 | 3.6 | 4.0 | 4.6 | 5.2 | 5.8 | 6.4 | 7.0 | 7.8 |  |
| 2008 | N | 114 | 973 | 407 | 343 | 521 | 473 | 461 | 493 | 547 | 674 | 570 | 692 | 687 | 682 |  |  |
|  | Cumulative N | 114 | 1,087 | 1,494 | 1,837 | 2,358 | 2,831 | 3,292 | 3,785 | 4,332 | 5,006 | 5,576 | 6,268 | 6,955 | 7,637 |  |  |
|  | Cumulative % | 0.1 | 1.1 | 1.5 | 1.8 | 2.3 | 2.8 | 3.3 | 3.7 | 4.3 | 4.9 | 5.5 | 6.2 | 6.9 | 7.6 |  |  |
| 2009 | N | 149 | 879 | 338 | 449 | 505 | 486 | 532 | 602 | 658 | 625 | 598 | 616 | 647 |  |  |  |
|  | Cumulative N | 149 | 1,028 | 1,366 | 1,815 | 2,320 | 2,806 | 3,338 | 3,940 | 4,598 | 5,223 | 5,821 | 6,437 | 7,084 |  |  |  |
|  | Cumulative % | 0.1 | 1.0 | 1.4 | 1.8 | 2.3 | 2.8 | 3.3 | 3.9 | 4.6 | 5.2 | 5.8 | 6.4 | 7.1 |  |  |  |
| 2010 | N | 130 | 697 | 488 | 490 | 537 | 546 | 567 | 658 | 620 | 634 | 699 | 656 |  |  |  |  |
|  | Cumulative N | 130 | 827 | 1,315 | 1,805 | 2,342 | 2,888 | 3,455 | 4,113 | 4,733 | 5,367 | 6,066 | 6,722 |  |  |  |  |
|  | Cumulative % | 0.1 | 0.8 | 1.3 | 1.8 | 2.4 | 2.9 | 3.5 | 4.1 | 4.8 | 5.4 | 6.1 | 6.8 |  |  |  |  |
| 2011 | N | 130 | 877 | 531 | 490 | 541 | 588 | 702 | 616 | 665 | 693 | 686 |  |  |  |  |  |
|  | Cumulative N | 130 | 1,007 | 1,538 | 2,028 | 2,569 | 3,157 | 3,859 | 4,475 | 5,140 | 5,833 | 6,519 |  |  |  |  |  |
|  | Cumulative % | 0.1 | 1.0 | 1.6 | 2.1 | 2.6 | 3.2 | 3.9 | 4.5 | 5.2 | 5.9 | 6.6 |  |  |  |  |  |
| 2012 | N | 274 | 958 | 527 | 514 | 604 | 682 | 677 | 672 | 684 | 717 |  |  |  |  |  |  |
|  | Cumulative N | 274 | 1,232 | 1,759 | 2,273 | 2,877 | 3,559 | 4,236 | 4,908 | 5,592 | 6,309 |  |  |  |  |  |  |
|  | Cumulative % | 0.3 | 1.2 | 1.7 | 2.2 | 2.8 | 3.5 | 4.2 | 4.8 | 5.5 | 6.2 |  |  |  |  |  |  |
| 2013 | N | 337 | 994 | 569 | 565 | 662 | 654 | 662 | 696 | 690 |  |  |  |  |  |  |  |
|  | Cumulative N | 337 | 1,331 | 1,900 | 2,465 | 3,127 | 3,781 | 4,443 | 5,139 | 5,829 |  |  |  |  |  |  |  |
|  | Cumulative % | 0.3 | 1.4 | 1.9 | 2.5 | 3.2 | 3.9 | 4.6 | 5.3 | 6.0 |  |  |  |  |  |  |  |
| 2014 | N | 393 | 977 | 603 | 641 | 615 | 676 | 745 | 653 |  |  |  |  |  |  |  |  |
|  | Cumulative N | 393 | 1,370 | 1,973 | 2,614 | 3,229 | 3,905 | 4,650 | 5,303 |  |  |  |  |  |  |  |  |
|  | Cumulative % | 0.4 | 1.4 | 2.0 | 2.7 | 3.3 | 4.0 | 4.8 | 5.4 |  |  |  |  |  |  |  |  |
| 2015 | N | 429 | 1,082 | 717 | 645 | 662 | 701 | 691 |  |  |  |  |  |  |  |  |  |
|  | Cumulative N | 429 | 1,511 | 2,228 | 2,873 | 3,535 | 4,236 | 4,927 |  |  |  |  |  |  |  |  |  |
|  | Cumulative % | 0.4 | 1.6 | 2.3 | 3.0 | 3.7 | 4.4 | 5.2 |  |  |  |  |  |  |  |  |  |
| 2016 | N | 569 | 1,190 | 712 | 652 | 759 | 715 |  |  |  |  |  |  |  |  |  |  |
|  | Cumulative N | 569 | 1,759 | 2,471 | 3,123 | 3,882 | 4,597 |  |  |  |  |  |  |  |  |  |  |
|  | Cumulative % | 0.6 | 1.8 | 2.6 | 3.2 | 4.0 | 4.8 |  |  |  |  |  |  |  |  |  |  |
| 2017 | N | 627 | 1,179 | 727 | 738 | 697 |  |  |  |  |  |  |  |  |  |  |  |
|  | Cumulative N | 627 | 1,806 | 2,533 | 3,271 | 3,968 |  |  |  |  |  |  |  |  |  |  |  |
|  | Cumulative % | 0.7 | 1.9 | 2.7 | 3.5 | 4.2 |  |  |  |  |  |  |  |  |  |  |  |
| 2018 | N | 602 | 1,270 | 784 | 721 |  |  |  |  |  |  |  |  |  |  |  |  |
|  | Cumulative N | 602 | 1,872 | 2,656 | 3,377 |  |  |  |  |  |  |  |  |  |  |  |  |
|  | Cumulative % | 0.6 | 2.0 | 2.8 | 3.6 |  |  |  |  |  |  |  |  |  |  |  |  |
| 2019 | N | 653 | 1,197 | 736 |  |  |  |  |  |  |  |  |  |  |  |  |  |
|  | Cumulative N | 653 | 1,850 | 2,586 |  |  |  |  |  |  |  |  |  |  |  |  |  |
|  | Cumulative % | 0.7 | 2.0 | 2.8 |  |  |  |  |  |  |  |  |  |  |  |  |  |
| 2020 | N | 734 | 1,126 |  |  |  |  |  |  |  |  |  |  |  |  |  |  |
|  | Cumulative N | 734 | 1,860 |  |  |  |  |  |  |  |  |  |  |  |  |  |  |
|  | Cumulative % | 0.8 | 2.1 |  |  |  |  |  |  |  |  |  |  |  |  |  |  |

**Table E4d.** Number, cumulative number and cumulative incidence of first-time child protections service-defined substantiation by year of age for non-Aboriginal children born 2006-2020.

| Birth-year cohort |  | Age |  |  |  |  |  |  |  |  |  |  |  |  |  |  |  |
| --- | --- | --- | --- | --- | --- | --- | --- | --- | --- | --- | --- | --- | --- | --- | --- | --- | --- |
|  |  | Birth | 1 | 2 | 3 | 4 | 5 | 6 | 7 | 8 | 9 | 10 | 11 | 12 | 13 | 14 | 15 |
| 2006 | N | 83 | 837 | 380 | 379 | 329 | 275 | 351 | 337 | 294 | 299 | 306 | 421 | 236 | 290 | 317 | 442 |
|  | Cumulative N | 83 | 920 | 1,300 | 1,679 | 2,008 | 2,283 | 2,634 | 2,971 | 3,265 | 3,564 | 3,870 | 4,291 | 4,527 | 4,817 | 5,134 | 5,576 |
|  | Cumulative % | 0.08 | 0.93 | 1.31 | 1.70 | 2.03 | 2.31 | 2.66 | 3.00 | 3.30 | 3.60 | 3.91 | 4.34 | 4.58 | 4.87 | 5.19 | 5.64 |
| 2007 | N | 100 | 878 | 421 | 312 | 280 | 444 | 395 | 342 | 299 | 361 | 413 | 260 | 291 | 303 | 430 |  |
|  | Cumulative N | 100 | 978 | 1,399 | 1,711 | 1,991 | 2,435 | 2,830 | 3,172 | 3,471 | 3,832 | 4,245 | 4,505 | 4,796 | 5,099 | 5,529 |  |
|  | Cumulative % | 0.1 | 1.0 | 1.4 | 1.7 | 2.0 | 2.4 | 2.8 | 3.1 | 3.4 | 3.8 | 4.2 | 4.5 | 4.7 | 5.0 | 5.5 |  |
| 2008 | N | 102 | 887 | 375 | 312 | 436 | 412 | 328 | 321 | 387 | 425 | 257 | 334 | 320 | 358 |  |  |
|  | Cumulative N | 102 | 989 | 1,364 | 1,676 | 2,112 | 2,524 | 2,852 | 3,173 | 3,560 | 3,985 | 4,242 | 4,576 | 4,896 | 5,254 |  |  |
|  | Cumulative % | 0.1 | 1.0 | 1.3 | 1.7 | 2.1 | 2.5 | 2.8 | 3.1 | 3.5 | 3.9 | 4.2 | 4.5 | 4.8 | 5.2 |  |  |
| 2009 | N | 137 | 797 | 298 | 408 | 433 | 349 | 354 | 387 | 437 | 270 | 266 | 293 | 326 |  |  |  |
|  | Cumulative N | 137 | 934 | 1,232 | 1,640 | 2,073 | 2,422 | 2,776 | 3,163 | 3,600 | 3,870 | 4,136 | 4,429 | 4,755 |  |  |  |
|  | Cumulative % | 0.1 | 0.9 | 1.2 | 1.6 | 2.1 | 2.4 | 2.8 | 3.2 | 3.6 | 3.9 | 4.1 | 4.4 | 4.8 |  |  |  |
| 2010 | N | 119 | 616 | 443 | 422 | 380 | 354 | 369 | 398 | 266 | 285 | 315 | 321 |  |  |  |  |
|  | Cumulative N | 119 | 735 | 1,178 | 1,600 | 1,980 | 2,334 | 2,703 | 3,101 | 3,367 | 3,652 | 3,967 | 4,288 |  |  |  |  |
|  | Cumulative % | 0.1 | 0.7 | 1.2 | 1.6 | 2.0 | 2.3 | 2.7 | 3.1 | 3.4 | 3.7 | 4.0 | 4.3 |  |  |  |  |
| 2011 | N | 113 | 759 | 472 | 367 | 357 | 364 | 449 | 256 | 287 | 325 | 340 |  |  |  |  |  |
|  | Cumulative N | 113 | 872 | 1,344 | 1,711 | 2,068 | 2,432 | 2,881 | 3,137 | 3,424 | 3,749 | 4,089 |  |  |  |  |  |
|  | Cumulative % | 0.1 | 0.9 | 1.4 | 1.7 | 2.1 | 2.5 | 2.9 | 3.2 | 3.5 | 3.8 | 4.1 |  |  |  |  |  |
| 2012 | N | 237 | 826 | 416 | 335 | 405 | 407 | 293 | 301 | 301 | 362 |  |  |  |  |  |  |
|  | Cumulative N | 237 | 1,063 | 1,479 | 1,814 | 2,219 | 2,626 | 2,919 | 3,220 | 3,521 | 3,883 |  |  |  |  |  |  |
|  | Cumulative % | 0.2 | 1.0 | 1.5 | 1.8 | 2.2 | 2.6 | 2.9 | 3.2 | 3.5 | 3.8 |  |  |  |  |  |  |
| 2013 | N | 302 | 781 | 386 | 394 | 431 | 259 | 314 | 328 | 340 |  |  |  |  |  |  |  |
|  | Cumulative N | 302 | 1,083 | 1,469 | 1,863 | 2,294 | 2,553 | 2,867 | 3,195 | 3,535 |  |  |  |  |  |  |  |
|  | Cumulative % | 0.3 | 1.1 | 1.5 | 1.9 | 2.4 | 2.6 | 2.9 | 3.3 | 3.6 |  |  |  |  |  |  |  |
| 2014 | N | 287 | 694 | 454 | 400 | 240 | 284 | 375 | 325 |  |  |  |  |  |  |  |  |
|  | Cumulative N | 287 | 981 | 1,435 | 1,835 | 2,075 | 2,359 | 2,734 | 3,059 |  |  |  |  |  |  |  |  |
|  | Cumulative % | 0.3 | 1.0 | 1.5 | 1.9 | 2.1 | 2.4 | 2.8 | 3.1 |  |  |  |  |  |  |  |  |
| 2015 | N | 317 | 785 | 484 | 279 | 311 | 325 | 347 |  |  |  |  |  |  |  |  |  |
|  | Cumulative N | 317 | 1,102 | 1,586 | 1,865 | 2,176 | 2,501 | 2,848 |  |  |  |  |  |  |  |  |  |
|  | Cumulative % | 0.3 | 1.2 | 1.7 | 2.0 | 2.3 | 2.6 | 3.0 |  |  |  |  |  |  |  |  |  |
| 2016 | N | 393 | 802 | 316 | 324 | 338 | 369 |  |  |  |  |  |  |  |  |  |  |
|  | Cumulative N | 393 | 1,195 | 1,511 | 1,835 | 2,173 | 2,542 |  |  |  |  |  |  |  |  |  |  |
|  | Cumulative % | 0.4 | 1.2 | 1.6 | 1.9 | 2.2 | 2.6 |  |  |  |  |  |  |  |  |  |  |
| 2017 | N | 425 | 571 | 359 | 352 | 375 |  |  |  |  |  |  |  |  |  |  |  |
|  | Cumulative N | 425 | 996 | 1,355 | 1,707 | 2,082 |  |  |  |  |  |  |  |  |  |  |  |
|  | Cumulative % | 0.5 | 1.1 | 1.5 | 1.8 | 2.2 |  |  |  |  |  |  |  |  |  |  |  |
| 2018 | N | 261 | 618 | 423 | 397 |  |  |  |  |  |  |  |  |  |  |  |  |
|  | Cumulative N | 261 | 879 | 1,302 | 1,699 |  |  |  |  |  |  |  |  |  |  |  |  |
|  | Cumulative % | 0.3 | 0.9 | 1.4 | 1.8 |  |  |  |  |  |  |  |  |  |  |  |  |
| 2019 | N | 370 | 595 | 396 |  |  |  |  |  |  |  |  |  |  |  |  |  |
|  | Cumulative N | 370 | 965 | 1,361 |  |  |  |  |  |  |  |  |  |  |  |  |  |
|  | Cumulative % | 0.4 | 1.1 | 1.5 |  |  |  |  |  |  |  |  |  |  |  |  |  |
| 2020 | N | 441 | 600 |  |  |  |  |  |  |  |  |  |  |  |  |  |  |
|  | Cumulative N | 441 | 1,041 |  |  |  |  |  |  |  |  |  |  |  |  |  |  |
|  | Cumulative % | 0.5 | 1.2 |  |  |  |  |  |  |  |  |  |  |  |  |  |  |

**Table E4e.** Number, cumulative number and cumulative incidence of first-time OOHC placements by year of age for non-Aboriginal children born 2006-2020.

| Birth-year cohort |  | Age |  |  |  |  |  |  |  |  |  |  |  |  |  |  |  |
| --- | --- | --- | --- | --- | --- | --- | --- | --- | --- | --- | --- | --- | --- | --- | --- | --- | --- |
|  |  | Birth | 1 | 2 | 3 | 4 | 5 | 6 | 7 | 8 | 9 | 10 | 11 | 12 | 13 | 14 | 15 |
| 2006 | N | 0 | 323 | 185 | 157 | 134 | 88 | 87 | 78 | 74 | 87 | 75 | 63 | 31 | 25 | 25 | 24 |
|  | Cumulative N | 0 | 323 | 508 | 665 | 799 | 887 | 974 | 1,052 | 1,126 | 1,213 | 1,288 | 1,351 | 1,382 | 1,407 | 1,432 | 1,456 |
|  | Cumulative % | 0.0 | 0.3 | 0.5 | 0.7 | 0.8 | 0.9 | 1.0 | 1.1 | 1.1 | 1.2 | 1.3 | 1.4 | 1.4 | 1.4 | 1.4 | 1.5 |
| 2007 | N | 0 | 432 | 193 | 129 | 121 | 100 | 99 | 83 | 98 | 106 | 66 | 30 | 34 | 30 | 38 |  |
|  | Cumulative N | 0 | 432 | 625 | 754 | 875 | 975 | 1,074 | 1,157 | 1,255 | 1,361 | 1,427 | 1,457 | 1,491 | 1,521 | 1,559 |  |
|  | Cumulative % | 0.0 | 0.4 | 0.6 | 0.7 | 0.9 | 1.0 | 1.1 | 1.1 | 1.2 | 1.3 | 1.4 | 1.4 | 1.5 | 1.5 | 1.5 |  |
| 2008 | N | 0 | 430 | 159 | 122 | 117 | 108 | 97 | 92 | 85 | 63 | 48 | 40 | 34 | 39 |  |  |
|  | Cumulative N | 0 | 430 | 589 | 711 | 828 | 936 | 1,033 | 1,125 | 1,210 | 1,273 | 1,321 | 1,361 | 1,395 | 1,434 |  |  |
|  | Cumulative % | 0.0 | 0.4 | 0.6 | 0.7 | 0.8 | 0.9 | 1.0 | 1.1 | 1.2 | 1.3 | 1.3 | 1.3 | 1.4 | 1.4 |  |  |
| 2009 | N | 0 | 424 | 128 | 101 | 100 | 95 | 95 | 107 | 66 | 46 | 28 | 47 | 38 |  |  |  |
|  | Cumulative N | 0 | 424 | 552 | 653 | 753 | 848 | 943 | 1,050 | 1,116 | 1,162 | 1,190 | 1,237 | 1,275 |  |  |  |
|  | Cumulative % | 0.0 | 0.4 | 0.6 | 0.7 | 0.8 | 0.8 | 0.9 | 1.0 | 1.1 | 1.2 | 1.2 | 1.2 | 1.3 |  |  |  |
| 2010 | N | 0 | 358 | 98 | 102 | 102 | 117 | 121 | 62 | 47 | 49 | 32 | 32 |  |  |  |  |
|  | Cumulative N | 0 | 358 | 456 | 558 | 660 | 777 | 898 | 960 | 1,007 | 1,056 | 1,088 | 1,120 |  |  |  |  |
|  | Cumulative % | 0.0 | 0.4 | 0.5 | 0.6 | 0.7 | 0.8 | 0.9 | 1.0 | 1.0 | 1.1 | 1.1 | 1.1 |  |  |  |  |
| 2011 | N | 0 | 356 | 130 | 111 | 107 | 116 | 83 | 37 | 43 | 47 | 29 |  |  |  |  |  |
|  | Cumulative N | 0 | 356 | 486 | 597 | 704 | 820 | 903 | 940 | 983 | 1,030 | 1,059 |  |  |  |  |  |
|  | Cumulative % | 0.0 | 0.4 | 0.5 | 0.6 | 0.7 | 0.8 | 0.9 | 1.0 | 1.0 | 1.0 | 1.1 |  |  |  |  |  |
| 2012 | N | 0 | 336 | 134 | 145 | 143 | 80 | 56 | 41 | 40 | 47 |  |  |  |  |  |  |
|  | Cumulative N | 0 | 336 | 470 | 615 | 758 | 838 | 894 | 935 | 975 | 1,022 |  |  |  |  |  |  |
|  | Cumulative % | 0.0 | 0.3 | 0.5 | 0.6 | 0.7 | 0.8 | 0.9 | 0.9 | 1.0 | 1.0 |  |  |  |  |  |  |
| 2013 | N | 0 | 366 | 125 | 140 | 89 | 59 | 46 | 38 | 32 |  |  |  |  |  |  |  |
|  | Cumulative N | 0 | 366 | 491 | 631 | 720 | 779 | 825 | 863 | 895 |  |  |  |  |  |  |  |
|  | Cumulative % | 0.0 | 0.4 | 0.5 | 0.6 | 0.7 | 0.8 | 0.8 | 0.9 | 0.9 |  |  |  |  |  |  |  |
| 2014 | N | 0 | 365 | 158 | 87 | 53 | 45 | 51 | 42 |  |  |  |  |  |  |  |  |
|  | Cumulative N | 0 | 365 | 523 | 610 | 663 | 708 | 759 | 801 |  |  |  |  |  |  |  |  |
|  | Cumulative % | 0.0 | 0.4 | 0.5 | 0.6 | 0.7 | 0.7 | 0.8 | 0.8 |  |  |  |  |  |  |  |  |
| 2015 | N | 0 | 383 | 100 | 72 | 56 | 41 | 53 |  |  |  |  |  |  |  |  |  |
|  | Cumulative N | 0 | 383 | 483 | 555 | 611 | 652 | 705 |  |  |  |  |  |  |  |  |  |
|  | Cumulative % | 0.0 | 0.4 | 0.5 | 0.6 | 0.6 | 0.7 | 0.7 |  |  |  |  |  |  |  |  |  |
| 2016 | N | 0 | 338 | 82 | 57 | 55 | 55 |  |  |  |  |  |  |  |  |  |  |
|  | Cumulative N | 0 | 338 | 420 | 477 | 532 | 587 |  |  |  |  |  |  |  |  |  |  |
|  | Cumulative % | 0.0 | 0.3 | 0.4 | 0.5 | 0.5 | 0.6 |  |  |  |  |  |  |  |  |  |  |
| 2017 | N | 0 | 291 | 83 | 59 | 73 |  |  |  |  |  |  |  |  |  |  |  |
|  | Cumulative N | 0 | 291 | 374 | 433 | 506 |  |  |  |  |  |  |  |  |  |  |  |
|  | Cumulative % | 0.0 | 0.3 | 0.4 | 0.5 | 0.5 |  |  |  |  |  |  |  |  |  |  |  |
| 2018 | N | 0 | 267 | 95 | 80 |  |  |  |  |  |  |  |  |  |  |  |  |
|  | Cumulative N | 0 | 267 | 362 | 442 |  |  |  |  |  |  |  |  |  |  |  |  |
|  | Cumulative % | 0.0 | 0.3 | 0.4 | 0.5 |  |  |  |  |  |  |  |  |  |  |  |  |
| 2019 | N | 0 | 224 | 77 |  |  |  |  |  |  |  |  |  |  |  |  |  |
|  | Cumulative N | 0 | 224 | 301 |  |  |  |  |  |  |  |  |  |  |  |  |  |
|  | Cumulative % | 0.0 | 0.2 | 0.3 |  |  |  |  |  |  |  |  |  |  |  |  |  |
| 2020 | N | 0 | 212 |  |  |  |  |  |  |  |  |  |  |  |  |  |  |
|  | Cumulative N | 0 | 212 |  |  |  |  |  |  |  |  |  |  |  |  |  |  |
|  | Cumulative % | 0.0 | 0.2 |  |  |  |  |  |  |  |  |  |  |  |  |  |  |

**Table E5a.** Number, cumulative number and cumulative incidence of first-time child protection reports by year of age for Aboriginal children born 2006-2020 (child protection service data only)

| Birth-year cohort |  | Age |  |  |  |  |  |  |  |  |  |  |  |  |  |  |  |
| --- | --- | --- | --- | --- | --- | --- | --- | --- | --- | --- | --- | --- | --- | --- | --- | --- | --- |
|  |  | Birth | 1 | 2 | 3 | 4 | 5 | 6 | 7 | 8 | 9 | 10 | 11 | 12 | 13 | 14 | 15 |
| 2006 | N | 720 | 1,297 | 503 | 384 | 230 | 154 | 156 | 120 | 114 | 113 | 116 | 93 | 77 | 81 | 92 | 107 |
|  | Cumulative N | 720 | 2,017 | 2,520 | 2,904 | 3,134 | 3,288 | 3,444 | 3,564 | 3,678 | 3,791 | 3,907 | 4,000 | 4,077 | 4,158 | 4,250 | 4,357 |
|  | Cumulative % | 12.0 | 33.5 | 41.8 | 48.1 | 51.9 | 54.4 | 56.9 | 58.9 | 60.7 | 62.6 | 64.4 | 66.0 | 67.2 | 68.5 | 70.0 | 71.8 |
| 2007 | N | 902 | 1,245 | 555 | 308 | 183 | 167 | 126 | 135 | 128 | 109 | 113 | 83 | 77 | 86 | 111 |  |
|  | Cumulative N | 902 | 2,147 | 2,702 | 3,010 | 3,193 | 3,360 | 3,486 | 3,621 | 3,749 | 3,858 | 3,971 | 4,054 | 4,131 | 4,217 | 4,328 |  |
|  | Cumulative % | 14.4 | 34.4 | 43.3 | 48.2 | 51.0 | 53.7 | 55.6 | 57.8 | 59.8 | 61.5 | 63.2 | 64.5 | 65.7 | 67.1 | 68.8 |  |
| 2008 | N | 1,060 | 1,268 | 420 | 249 | 235 | 181 | 148 | 143 | 121 | 123 | 82 | 97 | 114 | 121 |  |  |
|  | Cumulative N | 1,060 | 2,328 | 2,748 | 2,997 | 3,232 | 3,413 | 3,561 | 3,704 | 3,825 | 3,948 | 4,030 | 4,127 | 4,241 | 4,362 |  |  |
|  | Cumulative % | 17.2 | 37.9 | 44.7 | 48.7 | 52.5 | 55.4 | 57.8 | 60.0 | 62.0 | 63.9 | 65.2 | 66.8 | 68.6 | 70.5 |  |  |
| 2009 | N | 1,161 | 1,024 | 353 | 296 | 184 | 212 | 196 | 165 | 148 | 92 | 110 | 115 | 110 |  |  |  |
|  | Cumulative N | 1,161 | 2,185 | 2,538 | 2,834 | 3,018 | 3,230 | 3,426 | 3,591 | 3,739 | 3,831 | 3,941 | 4,056 | 4,166 |  |  |  |
|  | Cumulative % | 18.8 | 35.5 | 41.2 | 46.0 | 48.9 | 52.3 | 55.5 | 58.1 | 60.5 | 61.9 | 63.7 | 65.5 | 67.3 |  |  |  |
| 2010 | N | 1,013 | 870 | 432 | 354 | 251 | 223 | 195 | 167 | 135 | 112 | 140 | 101 |  |  |  |  |
|  | Cumulative N | 1,013 | 1,883 | 2,315 | 2,669 | 2,920 | 3,143 | 3,338 | 3,505 | 3,640 | 3,752 | 3,892 | 3,993 |  |  |  |  |
|  | Cumulative % | 16.4 | 30.5 | 37.4 | 43.1 | 47.2 | 50.8 | 53.9 | 56.6 | 58.7 | 60.5 | 62.8 | 64.4 |  |  |  |  |
| 2011 | N | 902 | 900 | 502 | 333 | 272 | 216 | 184 | 143 | 141 | 134 | 132 |  |  |  |  |  |
|  | Cumulative N | 902 | 1,802 | 2,304 | 2,637 | 2,909 | 3,125 | 3,309 | 3,452 | 3,593 | 3,727 | 3,859 |  |  |  |  |  |
|  | Cumulative % | 14.5 | 28.9 | 36.9 | 42.2 | 46.6 | 50.0 | 52.9 | 55.2 | 57.4 | 59.6 | 61.7 |  |  |  |  |  |
| 2012 | N | 948 | 938 | 500 | 355 | 288 | 225 | 188 | 169 | 161 | 133 |  |  |  |  |  |  |
|  | Cumulative N | 948 | 1,886 | 2,386 | 2,741 | 3,029 | 3,254 | 3,442 | 3,611 | 3,772 | 3,905 |  |  |  |  |  |  |
|  | Cumulative % | 14.9 | 29.7 | 37.6 | 43.2 | 47.7 | 51.2 | 54.1 | 56.8 | 59.3 | 61.4 |  |  |  |  |  |  |
| 2013 | N | 1,016 | 951 | 480 | 361 | 278 | 188 | 185 | 195 | 135 |  |  |  |  |  |  |  |
|  | Cumulative N | 1,016 | 1,967 | 2,447 | 2,808 | 3,086 | 3,274 | 3,459 | 3,654 | 3,789 |  |  |  |  |  |  |  |
|  | Cumulative % | 15.9 | 30.9 | 38.4 | 44.0 | 48.4 | 51.3 | 54.2 | 57.2 | 59.4 |  |  |  |  |  |  |  |
| 2014 | N | 1,033 | 928 | 483 | 365 | 203 | 177 | 206 | 182 |  |  |  |  |  |  |  |  |
|  | Cumulative N | 1,033 | 1,961 | 2,444 | 2,809 | 3,012 | 3,189 | 3,395 | 3,577 |  |  |  |  |  |  |  |  |
|  | Cumulative % | 16.6 | 31.6 | 39.3 | 45.2 | 48.5 | 51.3 | 54.6 | 57.6 |  |  |  |  |  |  |  |  |
| 2015 | N | 1,067 | 911 | 425 | 303 | 234 | 209 | 211 |  |  |  |  |  |  |  |  |  |
|  | Cumulative N | 1,067 | 1,978 | 2,403 | 2,706 | 2,940 | 3,149 | 3,360 |  |  |  |  |  |  |  |  |  |
|  | Cumulative % | 16.6 | 30.8 | 37.4 | 42.1 | 45.8 | 49.1 | 52.4 |  |  |  |  |  |  |  |  |  |
| 2016 | N | 1,067 | 850 | 409 | 269 | 247 | 190 |  |  |  |  |  |  |  |  |  |  |
|  | Cumulative N | 1,067 | 1,917 | 2,326 | 2,595 | 2,842 | 3,032 |  |  |  |  |  |  |  |  |  |  |
|  | Cumulative % | 16.2 | 29.1 | 35.3 | 39.4 | 43.2 | 46.1 |  |  |  |  |  |  |  |  |  |  |
| 2017 | N | 1,063 | 762 | 356 | 307 | 236 |  |  |  |  |  |  |  |  |  |  |  |
|  | Cumulative N | 1,063 | 1,825 | 2,181 | 2,488 | 2,724 |  |  |  |  |  |  |  |  |  |  |  |
|  | Cumulative % | 16.4 | 28.2 | 33.7 | 38.5 | 42.2 |  |  |  |  |  |  |  |  |  |  |  |
| 2018 | N | 954 | 786 | 433 | 269 |  |  |  |  |  |  |  |  |  |  |  |  |
|  | Cumulative N | 954 | 1,740 | 2,173 | 2,442 |  |  |  |  |  |  |  |  |  |  |  |  |
|  | Cumulative % | 14.4 | 26.2 | 32.8 | 36.8 |  |  |  |  |  |  |  |  |  |  |  |  |
| 2019 | N | 1,055 | 816 | 369 |  |  |  |  |  |  |  |  |  |  |  |  |  |
|  | Cumulative N | 1,055 | 1,871 | 2,240 |  |  |  |  |  |  |  |  |  |  |  |  |  |
|  | Cumulative % | 15.5 | 27.5 | 32.9 |  |  |  |  |  |  |  |  |  |  |  |  |  |
| 2020 | N | 1,155 | 760 |  |  |  |  |  |  |  |  |  |  |  |  |  |  |
|  | Cumulative N | 1,155 | 1,915 |  |  |  |  |  |  |  |  |  |  |  |  |  |  |
|  | Cumulative % | 16.6 | 27.5 |  |  |  |  |  |  |  |  |  |  |  |  |  |  |

**Table E5b.** Number, cumulative number and cumulative incidence of first-time child protection ROSH reports by year of age for Aboriginal children born 2006-2020 (child protection service data only)

| Birth-year cohort |  | Age |  |  |  |  |  |  |  |  |  |  |  |  |  |  |  |
| --- | --- | --- | --- | --- | --- | --- | --- | --- | --- | --- | --- | --- | --- | --- | --- | --- | --- |
|  |  | Birth | 1 | 2 | 3 | 4 | 5 | 6 | 7 | 8 | 9 | 10 | 11 | 12 | 13 | 14 | 15 |
| 2006 | N | 627 | 1,248 | 498 | 362 | 206 | 119 | 125 | 114 | 114 | 121 | 101 | 90 | 82 | 93 | 84 | 109 |
|  | Cumulative N | 627 | 1,875 | 2,373 | 2,735 | 2,941 | 3,060 | 3,185 | 3,299 | 3,413 | 3,534 | 3,635 | 3,725 | 3,807 | 3,900 | 3,984 | 4,093 |
|  | Cumulative % | 10.4 | 31.1 | 39.4 | 45.3 | 48.7 | 50.6 | 52.7 | 54.5 | 56.4 | 58.3 | 60.0 | 61.4 | 62.8 | 64.3 | 65.6 | 67.4 |
| 2007 | N | 860 | 1,147 | 526 | 286 | 151 | 138 | 112 | 126 | 108 | 111 | 108 | 95 | 85 | 98 | 108 |  |
|  | Cumulative N | 860 | 2,007 | 2,533 | 2,819 | 2,970 | 3,108 | 3,220 | 3,346 | 3,454 | 3,565 | 3,673 | 3,768 | 3,853 | 3,951 | 4,059 |  |
|  | Cumulative % | 13.8 | 32.1 | 40.6 | 45.1 | 47.5 | 49.7 | 51.4 | 53.4 | 55.1 | 56.8 | 58.5 | 60.0 | 61.3 | 62.9 | 64.5 |  |
| 2008 | N | 1,005 | 1,178 | 393 | 200 | 185 | 155 | 133 | 133 | 132 | 116 | 83 | 113 | 116 | 119 |  |  |
|  | Cumulative N | 1,005 | 2,183 | 2,576 | 2,776 | 2,961 | 3,116 | 3,249 | 3,382 | 3,514 | 3,630 | 3,713 | 3,826 | 3,942 | 4,061 |  |  |
|  | Cumulative % | 16.3 | 35.5 | 41.9 | 45.1 | 48.1 | 50.6 | 52.7 | 54.8 | 56.9 | 58.8 | 60.1 | 61.9 | 63.8 | 65.7 |  |  |
| 2009 | N | 1,092 | 914 | 284 | 245 | 171 | 196 | 184 | 152 | 158 | 113 | 130 | 125 | 107 |  |  |  |
|  | Cumulative N | 1,092 | 2,006 | 2,290 | 2,535 | 2,706 | 2,902 | 3,086 | 3,238 | 3,396 | 3,509 | 3,639 | 3,764 | 3,871 |  |  |  |
|  | Cumulative % | 17.7 | 32.6 | 37.2 | 41.1 | 43.9 | 47.0 | 49.9 | 52.4 | 54.9 | 56.7 | 58.8 | 60.8 | 62.5 |  |  |  |
| 2010 | N | 900 | 703 | 347 | 320 | 272 | 214 | 197 | 155 | 152 | 135 | 142 | 104 |  |  |  |  |
|  | Cumulative N | 900 | 1,603 | 1,950 | 2,270 | 2,542 | 2,756 | 2,953 | 3,108 | 3,260 | 3,395 | 3,537 | 3,641 |  |  |  |  |
|  | Cumulative % | 14.6 | 25.9 | 31.5 | 36.7 | 41.1 | 44.5 | 47.7 | 50.1 | 52.6 | 54.7 | 57.0 | 58.7 |  |  |  |  |
| 2011 | N | 680 | 692 | 450 | 373 | 280 | 244 | 192 | 152 | 162 | 155 | 134 |  |  |  |  |  |
|  | Cumulative N | 680 | 1,372 | 1,822 | 2,195 | 2,475 | 2,719 | 2,911 | 3,063 | 3,225 | 3,380 | 3,514 |  |  |  |  |  |
|  | Cumulative % | 10.9 | 22.0 | 29.2 | 35.2 | 39.6 | 43.5 | 46.5 | 49.0 | 51.5 | 54.0 | 56.1 |  |  |  |  |  |
| 2012 | N | 704 | 760 | 503 | 364 | 271 | 234 | 201 | 192 | 187 | 139 |  |  |  |  |  |  |
|  | Cumulative N | 704 | 1,464 | 1,967 | 2,331 | 2,602 | 2,836 | 3,037 | 3,229 | 3,416 | 3,555 |  |  |  |  |  |  |
|  | Cumulative % | 11.1 | 23.1 | 31.0 | 36.7 | 40.9 | 44.6 | 47.7 | 50.7 | 53.7 | 55.9 |  |  |  |  |  |  |
| 2013 | N | 784 | 859 | 448 | 346 | 280 | 221 | 209 | 209 | 138 |  |  |  |  |  |  |  |
|  | Cumulative N | 784 | 1,643 | 2,091 | 2,437 | 2,717 | 2,938 | 3,147 | 3,356 | 3,494 |  |  |  |  |  |  |  |
|  | Cumulative % | 12.3 | 25.8 | 32.8 | 38.2 | 42.6 | 46.0 | 49.3 | 52.6 | 54.7 |  |  |  |  |  |  |  |
| 2014 | N | 830 | 766 | 498 | 353 | 231 | 203 | 212 | 172 |  |  |  |  |  |  |  |  |
|  | Cumulative N | 830 | 1,596 | 2,094 | 2,447 | 2,678 | 2,881 | 3,093 | 3,265 |  |  |  |  |  |  |  |  |
|  | Cumulative % | 13.4 | 25.7 | 33.7 | 39.4 | 43.1 | 46.4 | 49.8 | 52.6 |  |  |  |  |  |  |  |  |
| 2015 | N | 866 | 802 | 425 | 291 | 255 | 228 | 218 |  |  |  |  |  |  |  |  |  |
|  | Cumulative N | 866 | 1,668 | 2,093 | 2,384 | 2,639 | 2,867 | 3,085 |  |  |  |  |  |  |  |  |  |
|  | Cumulative % | 13.5 | 26.0 | 32.6 | 37.1 | 41.1 | 44.7 | 48.1 |  |  |  |  |  |  |  |  |  |
| 2016 | N | 892 | 755 | 408 | 292 | 256 | 198 |  |  |  |  |  |  |  |  |  |  |
|  | Cumulative N | 892 | 1,647 | 2,055 | 2,347 | 2,603 | 2,801 |  |  |  |  |  |  |  |  |  |  |
|  | Cumulative % | 13.5 | 25.0 | 31.2 | 35.6 | 39.5 | 42.6 |  |  |  |  |  |  |  |  |  |  |
| 2017 | N | 912 | 717 | 372 | 304 | 230 |  |  |  |  |  |  |  |  |  |  |  |
|  | Cumulative N | 912 | 1,629 | 2,001 | 2,305 | 2,535 |  |  |  |  |  |  |  |  |  |  |  |
|  | Cumulative % | 14.1 | 25.2 | 31.0 | 35.7 | 39.3 |  |  |  |  |  |  |  |  |  |  |  |
| 2018 | N | 829 | 730 | 415 | 294 |  |  |  |  |  |  |  |  |  |  |  |  |
|  | Cumulative N | 829 | 1,559 | 1,974 | 2,268 |  |  |  |  |  |  |  |  |  |  |  |  |
|  | Cumulative % | 12.5 | 23.5 | 29.8 | 34.2 |  |  |  |  |  |  |  |  |  |  |  |  |
| 2019 | N | 937 | 768 | 369 |  |  |  |  |  |  |  |  |  |  |  |  |  |
|  | Cumulative N | 937 | 1,705 | 2,074 |  |  |  |  |  |  |  |  |  |  |  |  |  |
|  | Cumulative % | 13.8 | 25.1 | 30.5 |  |  |  |  |  |  |  |  |  |  |  |  |  |
| 2020 | N | 1,031 | 734 |  |  |  |  |  |  |  |  |  |  |  |  |  |  |
|  | Cumulative N | 1,031 | 1,765 |  |  |  |  |  |  |  |  |  |  |  |  |  |  |
|  | Cumulative % | 14.8 | 25.3 |  |  |  |  |  |  |  |  |  |  |  |  |  |  |

**Table E5c.** Number, cumulative number and cumulative incidence of first-time child protection investigations by year of age for Aboriginal children born 2006-2020 (child protection service data only)

| Birth-year cohort |  | Age |  |  |  |  |  |  |  |  |  |  |  |  |  |  |  |
| --- | --- | --- | --- | --- | --- | --- | --- | --- | --- | --- | --- | --- | --- | --- | --- | --- | --- |
|  |  | Birth | 1 | 2 | 3 | 4 | 5 | 6 | 7 | 8 | 9 | 10 | 11 | 12 | 13 | 14 | 15 |
| 2006 | N | 70 | 541 | 222 | 191 | 139 | 131 | 156 | 147 | 127 | 148 | 131 | 129 | 111 | 114 | 131 | 111 |
|  | Cumulative N | 70 | 611 | 833 | 1,024 | 1,163 | 1,294 | 1,450 | 1,597 | 1,724 | 1,872 | 2,003 | 2,132 | 2,243 | 2,357 | 2,488 | 2,599 |
|  | Cumulative % | 1.2 | 10.1 | 13.8 | 17.0 | 19.2 | 21.4 | 23.9 | 26.3 | 28.3 | 30.7 | 32.9 | 35.0 | 36.8 | 38.6 | 40.7 | 42.5 |
| 2007 | N | 79 | 495 | 228 | 194 | 151 | 188 | 176 | 147 | 135 | 150 | 131 | 118 | 134 | 147 | 113 |  |
|  | Cumulative N | 79 | 574 | 802 | 996 | 1,147 | 1,335 | 1,511 | 1,658 | 1,793 | 1,943 | 2,074 | 2,192 | 2,326 | 2,473 | 2,586 |  |
|  | Cumulative % | 1.3 | 9.2 | 12.8 | 15.9 | 18.3 | 21.3 | 24.0 | 26.3 | 28.4 | 30.8 | 32.8 | 34.7 | 36.8 | 39.1 | 40.8 |  |
| 2008 | N | 76 | 584 | 235 | 168 | 204 | 212 | 167 | 161 | 152 | 140 | 136 | 141 | 120 | 120 |  |  |
|  | Cumulative N | 76 | 660 | 895 | 1,063 | 1,267 | 1,479 | 1,646 | 1,807 | 1,959 | 2,099 | 2,235 | 2,376 | 2,496 | 2,616 |  |  |
|  | Cumulative % | 1.2 | 10.7 | 14.6 | 17.3 | 20.5 | 23.9 | 26.6 | 29.2 | 31.6 | 33.8 | 36.0 | 38.3 | 40.2 | 42.1 |  |  |
| 2009 | N | 106 | 496 | 200 | 235 | 206 | 174 | 189 | 158 | 143 | 140 | 138 | 153 | 142 |  |  |  |
|  | Cumulative N | 106 | 602 | 802 | 1,037 | 1,243 | 1,417 | 1,606 | 1,764 | 1,907 | 2,047 | 2,185 | 2,338 | 2,480 |  |  |  |
|  | Cumulative % | 1.7 | 9.8 | 13.0 | 16.8 | 20.1 | 22.9 | 25.9 | 28.5 | 30.7 | 33.0 | 35.2 | 37.6 | 39.9 |  |  |  |
| 2010 | N | 95 | 518 | 275 | 251 | 202 | 203 | 171 | 158 | 137 | 150 | 160 | 116 |  |  |  |  |
|  | Cumulative N | 95 | 613 | 888 | 1,139 | 1,341 | 1,544 | 1,715 | 1,873 | 2,010 | 2,160 | 2,320 | 2,436 |  |  |  |  |
|  | Cumulative % | 1.5 | 9.9 | 14.4 | 18.4 | 21.7 | 24.9 | 27.7 | 30.2 | 32.4 | 34.8 | 37.3 | 39.2 |  |  |  |  |
| 2011 | N | 112 | 565 | 294 | 219 | 212 | 221 | 190 | 154 | 143 | 165 | 119 |  |  |  |  |  |
|  | Cumulative N | 112 | 677 | 971 | 1,190 | 1,402 | 1,623 | 1,813 | 1,967 | 2,110 | 2,275 | 2,394 |  |  |  |  |  |
|  | Cumulative % | 1.8 | 10.8 | 15.6 | 19.1 | 22.4 | 25.9 | 29.0 | 31.4 | 33.7 | 36.3 | 38.2 |  |  |  |  |  |
| 2012 | N | 199 | 607 | 304 | 263 | 226 | 212 | 188 | 177 | 159 | 147 |  |  |  |  |  |  |
|  | Cumulative N | 199 | 806 | 1,110 | 1,373 | 1,599 | 1,811 | 1,999 | 2,176 | 2,335 | 2,482 |  |  |  |  |  |  |
|  | Cumulative % | 3.1 | 12.7 | 17.5 | 21.6 | 25.1 | 28.5 | 31.4 | 34.2 | 36.7 | 39.0 |  |  |  |  |  |  |
| 2013 | N | 268 | 649 | 309 | 271 | 228 | 175 | 179 | 204 | 146 |  |  |  |  |  |  |  |
|  | Cumulative N | 268 | 917 | 1,226 | 1,497 | 1,725 | 1,900 | 2,079 | 2,283 | 2,429 |  |  |  |  |  |  |  |
|  | Cumulative % | 4.2 | 14.4 | 19.2 | 23.5 | 27.0 | 29.8 | 32.6 | 35.7 | 38.0 |  |  |  |  |  |  |  |
| 2014 | N | 316 | 618 | 353 | 276 | 193 | 182 | 197 | 144 |  |  |  |  |  |  |  |  |
|  | Cumulative N | 316 | 934 | 1,287 | 1,563 | 1,756 | 1,938 | 2,135 | 2,279 |  |  |  |  |  |  |  |  |
|  | Cumulative % | 5.1 | 15.0 | 20.7 | 25.1 | 28.2 | 31.2 | 34.4 | 36.7 |  |  |  |  |  |  |  |  |
| 2015 | N | 362 | 651 | 316 | 227 | 213 | 220 | 191 |  |  |  |  |  |  |  |  |  |
|  | Cumulative N | 362 | 1,013 | 1,329 | 1,556 | 1,769 | 1,989 | 2,180 |  |  |  |  |  |  |  |  |  |
|  | Cumulative % | 5.6 | 15.8 | 20.7 | 24.2 | 27.6 | 31.0 | 34.0 |  |  |  |  |  |  |  |  |  |
| 2016 | N | 438 | 628 | 255 | 228 | 260 | 197 |  |  |  |  |  |  |  |  |  |  |
|  | Cumulative N | 438 | 1,066 | 1,321 | 1,549 | 1,809 | 2,006 |  |  |  |  |  |  |  |  |  |  |
|  | Cumulative % | 6.6 | 16.2 | 20.0 | 23.5 | 27.5 | 30.5 |  |  |  |  |  |  |  |  |  |  |
| 2017 | N | 427 | 595 | 303 | 283 | 233 |  |  |  |  |  |  |  |  |  |  |  |
|  | Cumulative N | 427 | 1,022 | 1,325 | 1,608 | 1,841 |  |  |  |  |  |  |  |  |  |  |  |
|  | Cumulative % | 6.6 | 15.8 | 20.5 | 24.9 | 28.5 |  |  |  |  |  |  |  |  |  |  |  |
| 2018 | N | 363 | 622 | 360 | 269 |  |  |  |  |  |  |  |  |  |  |  |  |
|  | Cumulative N | 363 | 985 | 1,345 | 1,614 |  |  |  |  |  |  |  |  |  |  |  |  |
|  | Cumulative % | 5.5 | 14.8 | 20.3 | 24.4 |  |  |  |  |  |  |  |  |  |  |  |  |
| 2019 | N | 518 | 682 | 326 |  |  |  |  |  |  |  |  |  |  |  |  |  |
|  | Cumulative N | 518 | 1,200 | 1,526 |  |  |  |  |  |  |  |  |  |  |  |  |  |
|  | Cumulative % | 7.6 | 17.6 | 22.5 |  |  |  |  |  |  |  |  |  |  |  |  |  |
| 2020 | N | 615 | 647 |  |  |  |  |  |  |  |  |  |  |  |  |  |  |
|  | Cumulative N | 615 | 1,262 |  |  |  |  |  |  |  |  |  |  |  |  |  |  |
|  | Cumulative % | 8.8 | 18.1 |  |  |  |  |  |  |  |  |  |  |  |  |  |  |

**Table E5d.** Number, cumulative number and cumulative incidence of first-time child protection service-defined substantiations by year of age for Aboriginal children born 2006- 2020 (child protection service data only)

| Birth-year cohort |  | Age |  |  |  |  |  |  |  |  |  |  |  |  |  |  |  |
| --- | --- | --- | --- | --- | --- | --- | --- | --- | --- | --- | --- | --- | --- | --- | --- | --- | --- |
|  |  | Birth | 1 | 2 | 3 | 4 | 5 | 6 | 7 | 8 | 9 | 10 | 11 | 12 | 13 | 14 | 15 |
| 2006 | N | 61 | 512 | 218 | 175 | 133 | 118 | 139 | 133 | 107 | 99 | 112 | 91 | 53 | 64 | 84 | 77 |
|  | Cumulative N | 61 | 573 | 791 | 966 | 1,099 | 1,217 | 1,356 | 1,489 | 1,596 | 1,695 | 1,807 | 1,898 | 1,951 | 2,015 | 2,099 | 2,176 |
|  | Cumulative % | 1.0 | 9.5 | 13.1 | 16.0 | 18.2 | 20.1 | 22.4 | 24.5 | 26.3 | 27.9 | 29.7 | 31.1 | 32.0 | 33.0 | 34.4 | 35.7 |
| 2007 | N | 70 | 469 | 216 | 179 | 138 | 173 | 165 | 116 | 91 | 126 | 106 | 49 | 63 | 85 | 72 |  |
|  | Cumulative N | 70 | 539 | 755 | 934 | 1,072 | 1,245 | 1,410 | 1,526 | 1,617 | 1,743 | 1,849 | 1,898 | 1,961 | 2,046 | 2,118 |  |
|  | Cumulative % | 1.1 | 8.6 | 12.1 | 14.9 | 17.1 | 19.8 | 22.4 | 24.2 | 25.7 | 27.6 | 29.3 | 30.1 | 31.0 | 32.4 | 33.5 |  |
| 2008 | N | 69 | 539 | 222 | 167 | 188 | 198 | 124 | 120 | 123 | 96 | 67 | 57 | 77 | 78 |  |  |
|  | Cumulative N | 69 | 608 | 830 | 997 | 1,185 | 1,383 | 1,507 | 1,627 | 1,750 | 1,846 | 1,913 | 1,970 | 2,047 | 2,125 |  |  |
|  | Cumulative % | 1.1 | 9.9 | 13.5 | 16.2 | 19.2 | 22.4 | 24.4 | 26.3 | 28.2 | 29.8 | 30.9 | 31.8 | 33.0 | 34.2 |  |  |
| 2009 | N | 97 | 460 | 188 | 216 | 196 | 135 | 125 | 116 | 100 | 65 | 56 | 91 | 89 |  |  |  |
|  | Cumulative N | 97 | 557 | 745 | 961 | 1,157 | 1,292 | 1,417 | 1,533 | 1,633 | 1,698 | 1,754 | 1,845 | 1,934 |  |  |  |
|  | Cumulative % | 1.6 | 9.0 | 12.1 | 15.6 | 18.7 | 20.9 | 22.9 | 24.7 | 26.3 | 27.4 | 28.3 | 29.7 | 31.1 |  |  |  |
| 2010 | N | 85 | 461 | 251 | 241 | 171 | 143 | 137 | 124 | 59 | 76 | 89 | 76 |  |  |  |  |
|  | Cumulative N | 85 | 546 | 797 | 1,038 | 1,209 | 1,352 | 1,489 | 1,613 | 1,672 | 1,748 | 1,837 | 1,913 |  |  |  |  |
|  | Cumulative % | 1.4 | 8.8 | 12.9 | 16.8 | 19.5 | 21.8 | 24.0 | 26.0 | 26.9 | 28.2 | 29.6 | 30.8 |  |  |  |  |
| 2011 | N | 100 | 492 | 277 | 180 | 158 | 167 | 135 | 89 | 79 | 103 | 86 |  |  |  |  |  |
|  | Cumulative N | 100 | 592 | 869 | 1,049 | 1,207 | 1,374 | 1,509 | 1,598 | 1,677 | 1,780 | 1,866 |  |  |  |  |  |
|  | Cumulative % | 1.6 | 9.5 | 13.9 | 16.8 | 19.3 | 22.0 | 24.1 | 25.5 | 26.8 | 28.4 | 29.8 |  |  |  |  |  |
| 2012 | N | 175 | 532 | 258 | 198 | 185 | 160 | 82 | 98 | 97 | 100 |  |  |  |  |  |  |
|  | Cumulative N | 175 | 707 | 965 | 1,163 | 1,348 | 1,508 | 1,590 | 1,688 | 1,785 | 1,885 |  |  |  |  |  |  |
|  | Cumulative % | 2.8 | 11.1 | 15.2 | 18.3 | 21.2 | 23.7 | 25.0 | 26.5 | 28.0 | 29.6 |  |  |  |  |  |  |
| 2013 | N | 245 | 517 | 230 | 233 | 177 | 97 | 92 | 132 | 101 |  |  |  |  |  |  |  |
|  | Cumulative N | 245 | 762 | 992 | 1,225 | 1,402 | 1,499 | 1,591 | 1,723 | 1,824 |  |  |  |  |  |  |  |
|  | Cumulative % | 3.8 | 12.0 | 15.6 | 19.2 | 22.0 | 23.5 | 24.9 | 27.0 | 28.6 |  |  |  |  |  |  |  |
| 2014 | N | 252 | 466 | 275 | 212 | 97 | 108 | 127 | 118 |  |  |  |  |  |  |  |  |
|  | Cumulative N | 252 | 718 | 993 | 1,205 | 1,302 | 1,410 | 1,537 | 1,655 |  |  |  |  |  |  |  |  |
|  | Cumulative % | 4.1 | 11.6 | 16.0 | 19.4 | 20.9 | 22.7 | 24.7 | 26.6 |  |  |  |  |  |  |  |  |
| 2015 | N | 278 | 484 | 252 | 130 | 138 | 135 | 138 |  |  |  |  |  |  |  |  |  |
|  | Cumulative N | 278 | 762 | 1,014 | 1,144 | 1,282 | 1,417 | 1,555 |  |  |  |  |  |  |  |  |  |
|  | Cumulative % | 4.3 | 11.9 | 15.8 | 17.8 | 20.0 | 22.1 | 24.3 |  |  |  |  |  |  |  |  |  |
| 2016 | N | 327 | 513 | 124 | 137 | 154 | 146 |  |  |  |  |  |  |  |  |  |  |
|  | Cumulative N | 327 | 840 | 964 | 1,101 | 1,255 | 1,401 |  |  |  |  |  |  |  |  |  |  |
|  | Cumulative % | 5.0 | 12.7 | 14.6 | 16.7 | 19.1 | 21.3 |  |  |  |  |  |  |  |  |  |  |
| 2017 | N | 297 | 336 | 210 | 193 | 173 |  |  |  |  |  |  |  |  |  |  |  |
|  | Cumulative N | 297 | 633 | 843 | 1,036 | 1,209 |  |  |  |  |  |  |  |  |  |  |  |
|  | Cumulative % | 4.6 | 9.8 | 13.0 | 16.0 | 18.7 |  |  |  |  |  |  |  |  |  |  |  |
| 2018 | N | 150 | 350 | 250 | 205 |  |  |  |  |  |  |  |  |  |  |  |  |
|  | Cumulative N | 150 | 500 | 750 | 955 |  |  |  |  |  |  |  |  |  |  |  |  |
|  | Cumulative % | 2.3 | 7.5 | 11.3 | 14.4 |  |  |  |  |  |  |  |  |  |  |  |  |
| 2019 | N | 304 | 451 | 228 |  |  |  |  |  |  |  |  |  |  |  |  |  |
|  | Cumulative N | 304 | 755 | 983 |  |  |  |  |  |  |  |  |  |  |  |  |  |
|  | Cumulative % | 4.5 | 11.1 | 14.5 |  |  |  |  |  |  |  |  |  |  |  |  |  |
| 2020 | N | 377 | 431 |  |  |  |  |  |  |  |  |  |  |  |  |  |  |
|  | Cumulative N | 377 | 808 |  |  |  |  |  |  |  |  |  |  |  |  |  |  |
|  | Cumulative % | 5.4 | 11.6 |  |  |  |  |  |  |  |  |  |  |  |  |  |  |

**Table E5e.** Number, cumulative number and cumulative incidence of first-time OOHC placements by year of age for Aboriginal children born 2006-2020 (child protection service data only)

| Birth-year cohort |  | Age |  |  |  |  |  |  |  |  |  |  |  |  |  |  |  |
| --- | --- | --- | --- | --- | --- | --- | --- | --- | --- | --- | --- | --- | --- | --- | --- | --- | --- |
|  |  | Birth | 1 | 2 | 3 | 4 | 5 | 6 | 7 | 8 | 9 | 10 | 11 | 12 | 13 | 14 | 15 |
| 2006 | N | 0 | 209 | 120 | 112 | 89 | 79 | 64 | 48 | 57 | 58 | 62 | 39 | 25 | 19 | 17 | 15 |
|  | Cumulative N | 0 | 209 | 329 | 441 | 530 | 609 | 673 | 721 | 778 | 836 | 898 | 937 | 962 | 981 | 998 | 1,013 |
|  | Cumulative % | 0.0 | 3.5 | 5.5 | 7.3 | 8.8 | 10.0 | 11.1 | 11.9 | 12.8 | 13.7 | 14.7 | 15.4 | 15.8 | 16.1 | 16.4 | 16.6 |
| 2007 | N | 0 | 232 | 118 | 93 | 77 | 57 | 60 | 68 | 71 | 69 | 39 | 27 | 26 | 23 | 17 |  |
|  | Cumulative N | 0 | 232 | 350 | 443 | 520 | 577 | 637 | 705 | 776 | 845 | 884 | 911 | 937 | 960 | 977 |  |
|  | Cumulative % | 0.0 | 3.7 | 5.6 | 7.1 | 8.3 | 9.2 | 10.1 | 11.2 | 12.3 | 13.4 | 14.0 | 14.4 | 14.8 | 15.2 | 15.5 |  |
| 2008 | N | 0 | 283 | 108 | 77 | 64 | 63 | 79 | 58 | 78 | 39 | 24 | 29 | 21 | 25 |  |  |
|  | Cumulative N | 0 | 283 | 391 | 468 | 532 | 595 | 674 | 732 | 810 | 849 | 873 | 902 | 923 | 948 |  |  |
|  | Cumulative % | 0.0 | 4.6 | 6.4 | 7.6 | 8.6 | 9.6 | 10.9 | 11.8 | 13.1 | 13.7 | 14.1 | 14.5 | 14.9 | 15.3 |  |  |
| 2009 | N | 0 | 268 | 103 | 85 | 84 | 63 | 68 | 66 | 40 | 30 | 20 | 25 | 23 |  |  |  |
|  | Cumulative N | 0 | 268 | 371 | 456 | 540 | 603 | 671 | 737 | 777 | 807 | 827 | 852 | 875 |  |  |  |
|  | Cumulative % | 0.0 | 4.3 | 6.0 | 7.4 | 8.7 | 9.7 | 10.8 | 11.9 | 12.5 | 13.0 | 13.3 | 13.7 | 14.1 |  |  |  |
| 2010 | N | 0 | 283 | 98 | 92 | 61 | 73 | 63 | 50 | 33 | 35 | 25 | 29 |  |  |  |  |
|  | Cumulative N | 0 | 283 | 381 | 473 | 534 | 607 | 670 | 720 | 753 | 788 | 813 | 842 |  |  |  |  |
|  | Cumulative % | 0.0 | 4.6 | 6.2 | 7.6 | 8.6 | 9.8 | 10.8 | 11.6 | 12.1 | 12.7 | 13.1 | 13.6 |  |  |  |  |
| 2011 | N | 0 | 268 | 94 | 82 | 80 | 100 | 55 | 26 | 29 | 28 | 25 |  |  |  |  |  |
|  | Cumulative N | 0 | 268 | 362 | 444 | 524 | 624 | 679 | 705 | 734 | 762 | 787 |  |  |  |  |  |
|  | Cumulative % | 0.0 | 4.3 | 5.8 | 7.1 | 8.4 | 10.0 | 10.8 | 11.3 | 11.7 | 12.2 | 12.6 |  |  |  |  |  |
| 2012 | N | 0 | 248 | 114 | 97 | 96 | 62 | 42 | 34 | 37 | 29 |  |  |  |  |  |  |
|  | Cumulative N | 0 | 248 | 362 | 459 | 555 | 617 | 659 | 693 | 730 | 759 |  |  |  |  |  |  |
|  | Cumulative % | 0.0 | 3.9 | 5.7 | 7.2 | 8.7 | 9.7 | 10.4 | 10.9 | 11.5 | 11.9 |  |  |  |  |  |  |
| 2013 | N | 0 | 256 | 134 | 112 | 81 | 53 | 50 | 52 | 40 |  |  |  |  |  |  |  |
|  | Cumulative N | 0 | 256 | 390 | 502 | 583 | 636 | 686 | 738 | 778 |  |  |  |  |  |  |  |
|  | Cumulative % | 0.0 | 4.0 | 6.1 | 7.9 | 9.1 | 10.0 | 10.7 | 11.6 | 12.2 |  |  |  |  |  |  |  |
| 2014 | N | 0 | 298 | 125 | 104 | 41 | 35 | 50 | 28 |  |  |  |  |  |  |  |  |
|  | Cumulative N | 0 | 298 | 423 | 527 | 568 | 603 | 653 | 681 |  |  |  |  |  |  |  |  |
|  | Cumulative % | 0.0 | 4.8 | 6.8 | 8.5 | 9.1 | 9.7 | 10.5 | 11.0 |  |  |  |  |  |  |  |  |
| 2015 | N | 0 | 311 | 111 | 52 | 50 | 41 | 42 |  |  |  |  |  |  |  |  |  |
|  | Cumulative N | 0 | 311 | 422 | 474 | 524 | 565 | 607 |  |  |  |  |  |  |  |  |  |
|  | Cumulative % | 0.0 | 4.8 | 6.6 | 7.4 | 8.2 | 8.8 | 9.5 |  |  |  |  |  |  |  |  |  |
| 2016 | N | 0 | 309 | 59 | 67 | 61 | 50 |  |  |  |  |  |  |  |  |  |  |
|  | Cumulative N | 0 | 309 | 368 | 435 | 496 | 546 |  |  |  |  |  |  |  |  |  |  |
|  | Cumulative % | 0.0 | 4.7 | 5.6 | 6.6 | 7.5 | 8.3 |  |  |  |  |  |  |  |  |  |  |
| 2017 | N | 0 | 213 | 83 | 60 | 65 |  |  |  |  |  |  |  |  |  |  |  |
|  | Cumulative N | 0 | 213 | 296 | 356 | 421 |  |  |  |  |  |  |  |  |  |  |  |
|  | Cumulative % | 0.0 | 3.3 | 4.6 | 5.5 | 6.5 |  |  |  |  |  |  |  |  |  |  |  |
| 2018 | N | 0 | 208 | 73 | 70 |  |  |  |  |  |  |  |  |  |  |  |  |
|  | Cumulative N | 0 | 208 | 281 | 351 |  |  |  |  |  |  |  |  |  |  |  |  |
|  | Cumulative % | 0.0 | 3.1 | 4.2 | 5.3 |  |  |  |  |  |  |  |  |  |  |  |  |
| 2019 | N | 0 | 207 | 92 |  |  |  |  |  |  |  |  |  |  |  |  |  |
|  | Cumulative N | 0 | 207 | 299 |  |  |  |  |  |  |  |  |  |  |  |  |  |
|  | Cumulative % | 0.0 | 3.0 | 4.4 |  |  |  |  |  |  |  |  |  |  |  |  |  |
| 2020 | N | 0 | 219 |  |  |  |  |  |  |  |  |  |  |  |  |  |  |
|  | Cumulative N | 0 | 219 |  |  |  |  |  |  |  |  |  |  |  |  |  |  |
|  | Cumulative % | 0.0 | 3.1 |  |  |  |  |  |  |  |  |  |  |  |  |  |  |

**Table E6a.** Number, cumulative number and cumulative incidence of first-time child protection reports by year of age for non-Aboriginal children born 2006-2020 (child protection service data only)

| Birth-year cohort |  | Age |  |  |  |  |  |  |  |  |  |  |  |  |  |  |  |
| --- | --- | --- | --- | --- | --- | --- | --- | --- | --- | --- | --- | --- | --- | --- | --- | --- | --- |
|  |  | Birth | 1 | 2 | 3 | 4 | 5 | 6 | 7 | 8 | 9 | 10 | 11 | 12 | 13 | 14 | 15 |
| 2006 | N | 1,550 | 4,896 | 3,106 | 2,944 | 2,257 | 1,594 | 1,669 | 1,624 | 1,610 | 1,569 | 1,606 | 1,667 | 1,680 | 1,796 | 2,118 | 2,479 |
|  | Cumulative N | 1,550 | 6,446 | 9,552 | 12,496 | 14,753 | 16,347 | 18,016 | 19,640 | 21,250 | 22,819 | 24,425 | 26,092 | 27,772 | 29,568 | 31,686 | 34,165 |
|  | Cumulative % | 1.9 | 7.7 | 11.4 | 14.9 | 17.6 | 19.4 | 21.4 | 23.2 | 25.1 | 26.9 | 28.7 | 30.6 | 32.5 | 34.5 | 36.8 | 39.6 |
| 2007 | N | 2,210 | 5,193 | 3,169 | 2,415 | 1,762 | 1,799 | 1,677 | 1,828 | 1,693 | 1,776 | 1,710 | 1,821 | 1,731 | 2,066 | 2,449 |  |
|  | Cumulative N | 2,210 | 7,403 | 10,572 | 12,987 | 14,749 | 16,548 | 18,225 | 20,053 | 21,746 | 23,522 | 25,232 | 27,053 | 28,784 | 30,850 | 33,299 |  |
|  | Cumulative % | 2.5 | 8.4 | 12.0 | 14.8 | 16.8 | 18.8 | 20.7 | 22.8 | 24.7 | 26.6 | 28.5 | 30.5 | 32.3 | 34.5 | 37.2 |  |
| 2008 | N | 2,547 | 5,053 | 2,449 | 1,881 | 1,932 | 1,880 | 1,937 | 1,880 | 1,797 | 1,889 | 1,919 | 1,849 | 2,010 | 2,305 |  |  |
|  | Cumulative N | 2,547 | 7,600 | 10,049 | 11,930 | 13,862 | 15,742 | 17,679 | 19,559 | 21,356 | 23,245 | 25,164 | 27,013 | 29,023 | 31,328 |  |  |
|  | Cumulative % | 2.8 | 8.5 | 11.2 | 13.4 | 15.6 | 17.7 | 19.8 | 21.9 | 23.8 | 25.8 | 27.9 | 29.9 | 32.0 | 34.4 |  |  |
| 2009 | N | 2,687 | 4,024 | 1,983 | 1,967 | 1,956 | 2,003 | 2,044 | 1,940 | 1,926 | 2,050 | 1,856 | 2,122 | 2,095 |  |  |  |
|  | Cumulative N | 2,687 | 6,711 | 8,694 | 10,661 | 12,617 | 14,620 | 16,664 | 18,604 | 20,530 | 22,580 | 24,436 | 26,558 | 28,653 |  |  |  |
|  | Cumulative % | 3.0 | 7.4 | 9.6 | 11.8 | 14.0 | 16.2 | 18.4 | 20.5 | 22.6 | 24.8 | 26.8 | 29.0 | 31.2 |  |  |  |
| 2010 | N | 2,077 | 3,016 | 2,234 | 2,116 | 2,218 | 2,165 | 2,009 | 2,133 | 2,171 | 1,981 | 2,211 | 2,401 |  |  |  |  |
|  | Cumulative N | 2,077 | 5,093 | 7,327 | 9,443 | 11,661 | 13,826 | 15,835 | 17,968 | 20,139 | 22,120 | 24,331 | 26,732 |  |  |  |  |
|  | Cumulative % | 2.3 | 5.6 | 8.1 | 10.4 | 12.8 | 15.1 | 17.3 | 19.5 | 21.8 | 23.9 | 26.2 | 28.7 |  |  |  |  |
| 2011 | N | 1,790 | 3,144 | 2,328 | 2,272 | 2,217 | 2,108 | 2,251 | 2,183 | 2,125 | 2,245 | 2,232 |  |  |  |  |  |
|  | Cumulative N | 1,790 | 4,934 | 7,262 | 9,534 | 11,751 | 13,859 | 16,110 | 18,293 | 20,418 | 22,663 | 24,895 |  |  |  |  |  |
|  | Cumulative % | 2.0 | 5.6 | 8.2 | 10.7 | 13.1 | 15.4 | 17.8 | 20.1 | 22.3 | 24.6 | 27.0 |  |  |  |  |  |
| 2012 | N | 1,878 | 3,314 | 2,416 | 2,265 | 2,192 | 2,240 | 2,382 | 2,302 | 2,294 | 2,387 |  |  |  |  |  |  |
|  | Cumulative N | 1,878 | 5,192 | 7,608 | 9,873 | 12,065 | 14,305 | 16,687 | 18,989 | 21,283 | 23,670 |  |  |  |  |  |  |
|  | Cumulative % | 2.0 | 5.7 | 8.3 | 10.7 | 13.1 | 15.4 | 17.9 | 20.3 | 22.7 | 25.2 |  |  |  |  |  |  |
| 2013 | N | 1,978 | 3,236 | 2,313 | 2,216 | 2,258 | 2,360 | 2,340 | 2,339 | 2,398 |  |  |  |  |  |  |  |
|  | Cumulative N | 1,978 | 5,214 | 7,527 | 9,743 | 12,001 | 14,361 | 16,701 | 19,040 | 21,438 |  |  |  |  |  |  |  |
|  | Cumulative % | 2.1 | 5.6 | 8.1 | 10.5 | 12.9 | 15.3 | 17.8 | 20.2 | 22.6 |  |  |  |  |  |  |  |
| 2014 | N | 2,060 | 3,225 | 2,283 | 2,180 | 2,236 | 2,194 | 2,498 | 2,524 |  |  |  |  |  |  |  |  |
|  | Cumulative N | 2,060 | 5,285 | 7,568 | 9,748 | 11,984 | 14,178 | 16,676 | 19,200 |  |  |  |  |  |  |  |  |
|  | Cumulative % | 2.3 | 5.9 | 8.4 | 10.7 | 13.1 | 15.4 | 18.0 | 20.7 |  |  |  |  |  |  |  |  |
| 2015 | N | 1,958 | 3,230 | 2,312 | 2,226 | 2,151 | 2,360 | 2,572 |  |  |  |  |  |  |  |  |  |
|  | Cumulative N | 1,958 | 5,188 | 7,500 | 9,726 | 11,877 | 14,237 | 16,809 |  |  |  |  |  |  |  |  |  |
|  | Cumulative % | 2.1 | 5.7 | 8.2 | 10.5 | 12.8 | 15.3 | 18.0 |  |  |  |  |  |  |  |  |  |
| 2016 | N | 2,170 | 3,142 | 2,324 | 2,249 | 2,374 | 2,476 |  |  |  |  |  |  |  |  |  |  |
|  | Cumulative N | 2,170 | 5,312 | 7,636 | 9,885 | 12,259 | 14,735 |  |  |  |  |  |  |  |  |  |  |
|  | Cumulative % | 2.3 | 5.6 | 8.0 | 10.4 | 12.8 | 15.4 |  |  |  |  |  |  |  |  |  |  |
| 2017 | N | 2,212 | 3,178 | 2,322 | 2,268 | 2,362 |  |  |  |  |  |  |  |  |  |  |  |
|  | Cumulative N | 2,212 | 5,390 | 7,712 | 9,980 | 12,342 |  |  |  |  |  |  |  |  |  |  |  |
|  | Cumulative % | 2.4 | 5.9 | 8.4 | 10.9 | 13.4 |  |  |  |  |  |  |  |  |  |  |  |
| 2018 | N | 2,066 | 3,545 | 2,475 | 2,302 |  |  |  |  |  |  |  |  |  |  |  |  |
|  | Cumulative N | 2,066 | 5,611 | 8,086 | 10,388 |  |  |  |  |  |  |  |  |  |  |  |  |
|  | Cumulative % | 2.3 | 6.2 | 8.9 | 11.5 |  |  |  |  |  |  |  |  |  |  |  |  |
| 2019 | N | 2,240 | 3,263 | 2,356 |  |  |  |  |  |  |  |  |  |  |  |  |  |
|  | Cumulative N | 2,240 | 5,503 | 7,859 |  |  |  |  |  |  |  |  |  |  |  |  |  |
|  | Cumulative % | 2.4 | 6.0 | 8.6 |  |  |  |  |  |  |  |  |  |  |  |  |  |
| 2020 | N | 2,178 | 3,277 |  |  |  |  |  |  |  |  |  |  |  |  |  |  |
|  | Cumulative N | 2,178 | 5,455 |  |  |  |  |  |  |  |  |  |  |  |  |  |  |
|  | Cumulative % | 2.4 | 6.1 |  |  |  |  |  |  |  |  |  |  |  |  |  |  |

**Table E6b.** Number, cumulative number and cumulative incidence of first-time child protection ROSH reports by year of age for non-Aboriginal children born 2006-2020 (child protection service data only)

| Birth-year cohort |  | Age |  |  |  |  |  |  |  |  |  |  |  |  |  |  |  |
| --- | --- | --- | --- | --- | --- | --- | --- | --- | --- | --- | --- | --- | --- | --- | --- | --- | --- |
|  |  | Birth | 1 | 2 | 3 | 4 | 5 | 6 | 7 | 8 | 9 | 10 | 11 | 12 | 13 | 14 | 15 |
| 2006 | N | 1,311 | 4,141 | 2,578 | 2,552 | 1,834 | 1,042 | 1,009 | 1,092 | 1,176 | 1,126 | 1,153 | 1,300 | 1,381 | 1,488 | 1,797 | 2,149 |
|  | Cumulative N | 1,311 | 5,452 | 8,030 | 10,582 | 12,416 | 13,458 | 14,467 | 15,559 | 16,735 | 17,861 | 19,014 | 20,314 | 21,695 | 23,183 | 24,980 | 27,129 |
|  | Cumulative % | 1.6 | 6.5 | 9.6 | 12.6 | 14.8 | 16.0 | 17.2 | 18.4 | 19.8 | 21.1 | 22.4 | 23.8 | 25.4 | 27.1 | 29.1 | 31.5 |
| 2007 | N | 2,057 | 4,312 | 2,732 | 1,951 | 1,113 | 1,099 | 1,095 | 1,269 | 1,225 | 1,320 | 1,370 | 1,442 | 1,497 | 1,758 | 2,141 |  |
|  | Cumulative N | 2,057 | 6,369 | 9,101 | 11,052 | 12,165 | 13,264 | 14,359 | 15,628 | 16,853 | 18,173 | 19,543 | 20,985 | 22,482 | 24,240 | 26,381 |  |
|  | Cumulative % | 2.3 | 7.2 | 10.3 | 12.6 | 13.8 | 15.1 | 16.3 | 17.8 | 19.1 | 20.6 | 22.1 | 23.6 | 25.2 | 27.1 | 29.4 |  |
| 2008 | N | 2,373 | 4,354 | 1,954 | 1,163 | 1,175 | 1,207 | 1,371 | 1,341 | 1,357 | 1,488 | 1,554 | 1,584 | 1,696 | 2,000 |  |  |
|  | Cumulative N | 2,373 | 6,727 | 8,681 | 9,844 | 11,019 | 12,226 | 13,597 | 14,938 | 16,295 | 17,783 | 19,337 | 20,921 | 22,617 | 24,617 |  |  |
|  | Cumulative % | 2.7 | 7.5 | 9.7 | 11.0 | 12.4 | 13.7 | 15.2 | 16.7 | 18.2 | 19.8 | 21.4 | 23.1 | 24.9 | 27.0 |  |  |
| 2009 | N | 2,532 | 3,257 | 1,271 | 1,216 | 1,259 | 1,394 | 1,426 | 1,450 | 1,516 | 1,682 | 1,582 | 1,853 | 1,805 |  |  |  |
|  | Cumulative N | 2,532 | 5,789 | 7,060 | 8,276 | 9,535 | 10,929 | 12,355 | 13,805 | 15,321 | 17,003 | 18,585 | 20,438 | 22,243 |  |  |  |
|  | Cumulative % | 2.8 | 6.4 | 7.8 | 9.1 | 10.6 | 12.1 | 13.7 | 15.2 | 16.8 | 18.6 | 20.3 | 22.3 | 24.2 |  |  |  |
| 2010 | N | 1,709 | 1,963 | 1,369 | 1,361 | 1,570 | 1,539 | 1,528 | 1,692 | 1,751 | 1,699 | 1,970 | 2,064 |  |  |  |  |
|  | Cumulative N | 1,709 | 3,672 | 5,041 | 6,402 | 7,972 | 9,511 | 11,039 | 12,731 | 14,482 | 16,181 | 18,151 | 20,215 |  |  |  |  |
|  | Cumulative % | 1.9 | 4.0 | 5.5 | 7.0 | 8.8 | 10.4 | 12.0 | 13.8 | 15.7 | 17.4 | 19.5 | 21.6 |  |  |  |  |
| 2011 | N | 1,153 | 2,019 | 1,596 | 1,600 | 1,553 | 1,598 | 1,735 | 1,796 | 1,874 | 1,891 | 1,922 |  |  |  |  |  |
|  | Cumulative N | 1,153 | 3,172 | 4,768 | 6,368 | 7,921 | 9,519 | 11,254 | 13,050 | 14,924 | 16,815 | 18,737 |  |  |  |  |  |
|  | Cumulative % | 1.3 | 3.6 | 5.4 | 7.1 | 8.8 | 10.6 | 12.4 | 14.3 | 16.3 | 18.2 | 20.2 |  |  |  |  |  |
| 2012 | N | 1,204 | 2,086 | 1,731 | 1,625 | 1,654 | 1,728 | 1,882 | 1,931 | 1,999 | 2,043 |  |  |  |  |  |  |
|  | Cumulative N | 1,204 | 3,290 | 5,021 | 6,646 | 8,300 | 10,028 | 11,910 | 13,841 | 15,840 | 17,883 |  |  |  |  |  |  |
|  | Cumulative % | 1.3 | 3.6 | 5.5 | 7.2 | 9.0 | 10.8 | 12.8 | 14.8 | 16.9 | 19.0 |  |  |  |  |  |  |
| 2013 | N | 1,268 | 2,282 | 1,659 | 1,608 | 1,697 | 1,837 | 1,992 | 2,026 | 2,056 |  |  |  |  |  |  |  |
|  | Cumulative N | 1,268 | 3,550 | 5,209 | 6,817 | 8,514 | 10,351 | 12,343 | 14,369 | 16,425 |  |  |  |  |  |  |  |
|  | Cumulative % | 1.4 | 3.8 | 5.6 | 7.3 | 9.1 | 11.0 | 13.1 | 15.2 | 17.3 |  |  |  |  |  |  |  |
| 2014 | N | 1,405 | 2,191 | 1,669 | 1,702 | 1,729 | 1,816 | 2,119 | 2,128 |  |  |  |  |  |  |  |  |
|  | Cumulative N | 1,405 | 3,596 | 5,265 | 6,967 | 8,696 | 10,512 | 12,631 | 14,759 |  |  |  |  |  |  |  |  |
|  | Cumulative % | 1.6 | 4.0 | 5.8 | 7.6 | 9.5 | 11.4 | 13.6 | 15.9 |  |  |  |  |  |  |  |  |
| 2015 | N | 1,351 | 2,350 | 1,770 | 1,702 | 1,773 | 1,995 | 2,188 |  |  |  |  |  |  |  |  |  |
|  | Cumulative N | 1,351 | 3,701 | 5,471 | 7,173 | 8,946 | 10,941 | 13,129 |  |  |  |  |  |  |  |  |  |
|  | Cumulative % | 1.5 | 4.0 | 5.9 | 7.8 | 9.6 | 11.8 | 14.0 |  |  |  |  |  |  |  |  |  |
| 2016 | N | 1,599 | 2,309 | 1,830 | 1,842 | 2,001 | 2,086 |  |  |  |  |  |  |  |  |  |  |
|  | Cumulative N | 1,599 | 3,908 | 5,738 | 7,580 | 9,581 | 11,667 |  |  |  |  |  |  |  |  |  |  |
|  | Cumulative % | 1.7 | 4.1 | 6.0 | 7.9 | 10.0 | 12.2 |  |  |  |  |  |  |  |  |  |  |
| 2017 | N | 1,702 | 2,441 | 1,910 | 1,853 | 2,027 |  |  |  |  |  |  |  |  |  |  |  |
|  | Cumulative N | 1,702 | 4,143 | 6,053 | 7,906 | 9,933 |  |  |  |  |  |  |  |  |  |  |  |
|  | Cumulative % | 1.9 | 4.5 | 6.6 | 8.6 | 10.8 |  |  |  |  |  |  |  |  |  |  |  |
| 2018 | N | 1,618 | 2,768 | 2,066 | 1,929 |  |  |  |  |  |  |  |  |  |  |  |  |
|  | Cumulative N | 1,618 | 4,386 | 6,452 | 8,381 |  |  |  |  |  |  |  |  |  |  |  |  |
|  | Cumulative % | 1.8 | 4.8 | 7.1 | 9.3 |  |  |  |  |  |  |  |  |  |  |  |  |
| 2019 | N | 1,791 | 2,674 | 1,959 |  |  |  |  |  |  |  |  |  |  |  |  |  |
|  | Cumulative N | 1,791 | 4,465 | 6,424 |  |  |  |  |  |  |  |  |  |  |  |  |  |
|  | Cumulative % | 2.0 | 4.9 | 7.0 |  |  |  |  |  |  |  |  |  |  |  |  |  |
| 2020 | N | 1,751 | 2,689 |  |  |  |  |  |  |  |  |  |  |  |  |  |  |
|  | Cumulative N | 1,751 | 4,440 |  |  |  |  |  |  |  |  |  |  |  |  |  |  |
|  | Cumulative % | 2.0 | 5.0 |  |  |  |  |  |  |  |  |  |  |  |  |  |  |

**Table E6c.**
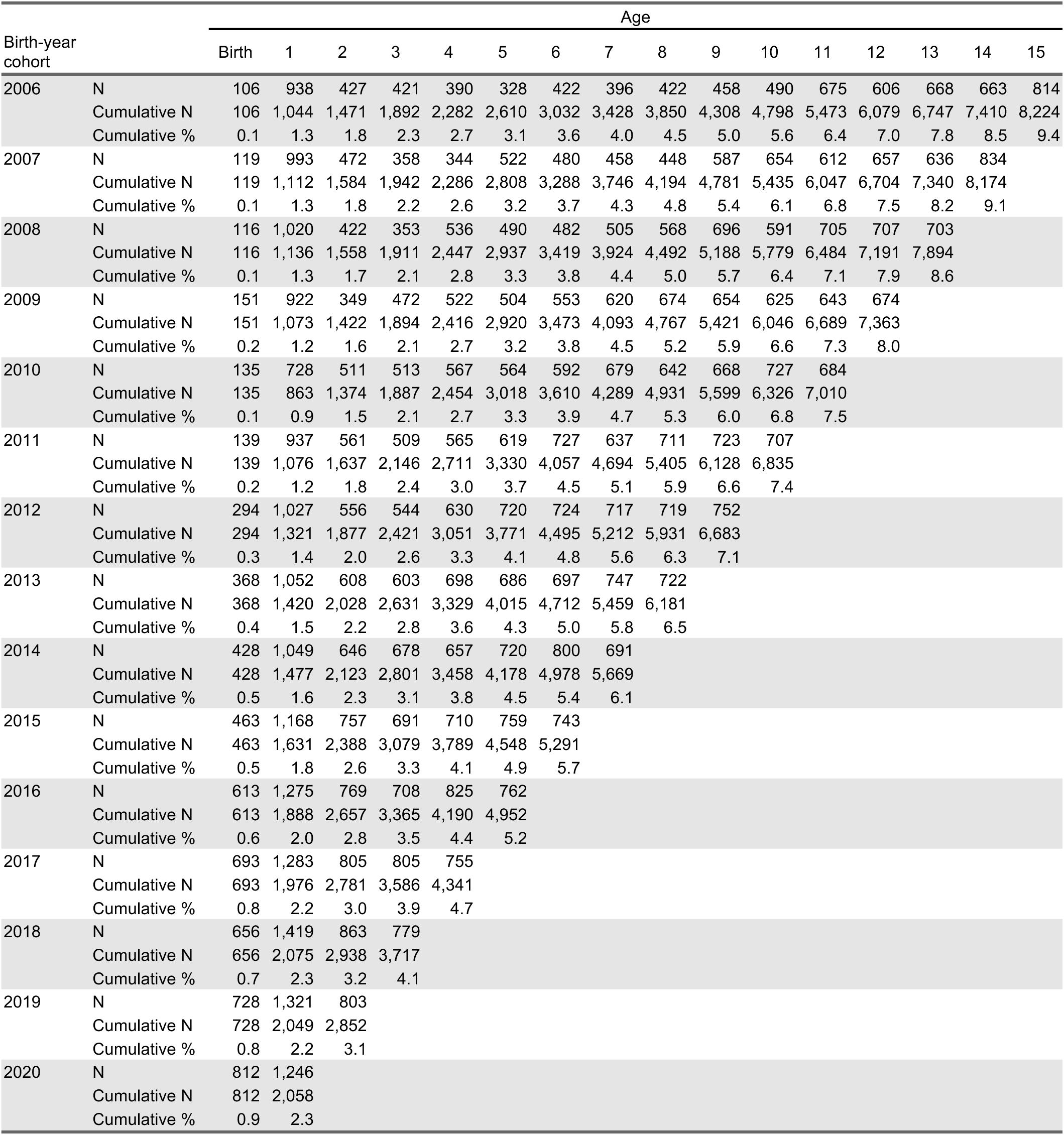
Number, cumulative number and cumulative incidence of first-time child protection investigations by year of age for non-Aboriginal children born 2006-2020 (child protection service data only)

| Birth-year cohort |  | Age |  |  |  |  |  |  |  |  |  |  |  |  |  |  |  |
| --- | --- | --- | --- | --- | --- | --- | --- | --- | --- | --- | --- | --- | --- | --- | --- | --- | --- |
|  |  | Birth | 1 | 2 | 3 | 4 | 5 | 6 | 7 | 8 | 9 | 10 | 11 | 12 | 13 | 14 | 15 |
| 2006 | N | 106 | 938 | 427 | 421 | 390 | 328 | 422 | 396 | 422 | 458 | 490 | 675 | 606 | 668 | 663 | 814 |
|  | Cumulative N | 106 | 1,044 | 1,471 | 1,892 | 2,282 | 2,610 | 3,032 | 3,428 | 3,850 | 4,308 | 4,798 | 5,473 | 6,079 | 6,747 | 7,410 | 8,224 |
|  | Cumulative % | 0.1 | 1.3 | 1.8 | 2.3 | 2.7 | 3.1 | 3.6 | 4.0 | 4.5 | 5.0 | 5.6 | 6.4 | 7.0 | 7.8 | 8.5 | 9.4 |
| 2007 | N | 119 | 993 | 472 | 358 | 344 | 522 | 480 | 458 | 448 | 587 | 654 | 612 | 657 | 636 | 834 |  |
|  | Cumulative N | 119 | 1,112 | 1,584 | 1,942 | 2,286 | 2,808 | 3,288 | 3,746 | 4,194 | 4,781 | 5,435 | 6,047 | 6,704 | 7,340 | 8,174 |  |
|  | Cumulative % | 0.1 | 1.3 | 1.8 | 2.2 | 2.6 | 3.2 | 3.7 | 4.3 | 4.8 | 5.4 | 6.1 | 6.8 | 7.5 | 8.2 | 9.1 |  |
| 2008 | N | 116 | 1,020 | 422 | 353 | 536 | 490 | 482 | 505 | 568 | 696 | 591 | 705 | 707 | 703 |  |  |
|  | Cumulative N | 116 | 1,136 | 1,558 | 1,911 | 2,447 | 2,937 | 3,419 | 3,924 | 4,492 | 5,188 | 5,779 | 6,484 | 7,191 | 7,894 |  |  |
|  | Cumulative % | 0.1 | 1.3 | 1.7 | 2.1 | 2.8 | 3.3 | 3.8 | 4.4 | 5.0 | 5.7 | 6.4 | 7.1 | 7.9 | 8.6 |  |  |
| 2009 | N | 151 | 922 | 349 | 472 | 522 | 504 | 553 | 620 | 674 | 654 | 625 | 643 | 674 |  |  |  |
|  | Cumulative N | 151 | 1,073 | 1,422 | 1,894 | 2,416 | 2,920 | 3,473 | 4,093 | 4,767 | 5,421 | 6,046 | 6,689 | 7,363 |  |  |  |
|  | Cumulative % | 0.2 | 1.2 | 1.6 | 2.1 | 2.7 | 3.2 | 3.8 | 4.5 | 5.2 | 5.9 | 6.6 | 7.3 | 8.0 |  |  |  |
| 2010 | N | 135 | 728 | 511 | 513 | 567 | 564 | 592 | 679 | 642 | 668 | 727 | 684 |  |  |  |  |
|  | Cumulative N | 135 | 863 | 1,374 | 1,887 | 2,454 | 3,018 | 3,610 | 4,289 | 4,931 | 5,599 | 6,326 | 7,010 |  |  |  |  |
|  | Cumulative % | 0.1 | 0.9 | 1.5 | 2.1 | 2.7 | 3.3 | 3.9 | 4.7 | 5.3 | 6.0 | 6.8 | 7.5 |  |  |  |  |
| 2011 | N | 139 | 937 | 561 | 509 | 565 | 619 | 727 | 637 | 711 | 723 | 707 |  |  |  |  |  |
|  | Cumulative N | 139 | 1,076 | 1,637 | 2,146 | 2,711 | 3,330 | 4,057 | 4,694 | 5,405 | 6,128 | 6,835 |  |  |  |  |  |
|  | Cumulative % | 0.2 | 1.2 | 1.8 | 2.4 | 3.0 | 3.7 | 4.5 | 5.1 | 5.9 | 6.6 | 7.4 |  |  |  |  |  |
| 2012 | N | 294 | 1,027 | 556 | 544 | 630 | 720 | 724 | 717 | 719 | 752 |  |  |  |  |  |  |
|  | Cumulative N | 294 | 1,321 | 1,877 | 2,421 | 3,051 | 3,771 | 4,495 | 5,212 | 5,931 | 6,683 |  |  |  |  |  |  |
|  | Cumulative % | 0.3 | 1.4 | 2.0 | 2.6 | 3.3 | 4.1 | 4.8 | 5.6 | 6.3 | 7.1 |  |  |  |  |  |  |
| 2013 | N | 368 | 1,052 | 608 | 603 | 698 | 686 | 697 | 747 | 722 |  |  |  |  |  |  |  |
|  | Cumulative N | 368 | 1,420 | 2,028 | 2,631 | 3,329 | 4,015 | 4,712 | 5,459 | 6,181 |  |  |  |  |  |  |  |
|  | Cumulative % | 0.4 | 1.5 | 2.2 | 2.8 | 3.6 | 4.3 | 5.0 | 5.8 | 6.5 |  |  |  |  |  |  |  |
| 2014 | N | 428 | 1,049 | 646 | 678 | 657 | 720 | 800 | 691 |  |  |  |  |  |  |  |  |
|  | Cumulative N | 428 | 1,477 | 2,123 | 2,801 | 3,458 | 4,178 | 4,978 | 5,669 |  |  |  |  |  |  |  |  |
|  | Cumulative % | 0.5 | 1.6 | 2.3 | 3.1 | 3.8 | 4.5 | 5.4 | 6.1 |  |  |  |  |  |  |  |  |
| 2015 | N | 463 | 1,168 | 757 | 691 | 710 | 759 | 743 |  |  |  |  |  |  |  |  |  |
|  | Cumulative N | 463 | 1,631 | 2,388 | 3,079 | 3,789 | 4,548 | 5,291 |  |  |  |  |  |  |  |  |  |
|  | Cumulative % | 0.5 | 1.8 | 2.6 | 3.3 | 4.1 | 4.9 | 5.7 |  |  |  |  |  |  |  |  |  |
| 2016 | N | 613 | 1,275 | 769 | 708 | 825 | 762 |  |  |  |  |  |  |  |  |  |  |
|  | Cumulative N | 613 | 1,888 | 2,657 | 3,365 | 4,190 | 4,952 |  |  |  |  |  |  |  |  |  |  |
|  | Cumulative % | 0.6 | 2.0 | 2.8 | 3.5 | 4.4 | 5.2 |  |  |  |  |  |  |  |  |  |  |
| 2017 | N | 693 | 1,283 | 805 | 805 | 755 |  |  |  |  |  |  |  |  |  |  |  |
|  | Cumulative N | 693 | 1,976 | 2,781 | 3,586 | 4,341 |  |  |  |  |  |  |  |  |  |  |  |
|  | Cumulative % | 0.8 | 2.2 | 3.0 | 3.9 | 4.7 |  |  |  |  |  |  |  |  |  |  |  |
| 2018 | N | 656 | 1,419 | 863 | 779 |  |  |  |  |  |  |  |  |  |  |  |  |
|  | Cumulative N | 656 | 2,075 | 2,938 | 3,717 |  |  |  |  |  |  |  |  |  |  |  |  |
|  | Cumulative % | 0.7 | 2.3 | 3.2 | 4.1 |  |  |  |  |  |  |  |  |  |  |  |  |
| 2019 | N | 728 | 1,321 | 803 |  |  |  |  |  |  |  |  |  |  |  |  |  |
|  | Cumulative N | 728 | 2,049 | 2,852 |  |  |  |  |  |  |  |  |  |  |  |  |  |
|  | Cumulative % | 0.8 | 2.2 | 3.1 |  |  |  |  |  |  |  |  |  |  |  |  |  |
| 2020 | N | 812 | 1,246 |  |  |  |  |  |  |  |  |  |  |  |  |  |  |
|  | Cumulative N | 812 | 2,058 |  |  |  |  |  |  |  |  |  |  |  |  |  |  |
|  | Cumulative % | 0.9 | 2.3 |  |  |  |  |  |  |  |  |  |  |  |  |  |  |

**Table E6d.** Number, cumulative number and cumulative incidence of first-time child protection-defined substantiations by year of age for non-Aboriginal children born 2006- 2020 (child protection service data only)

| Birth-year cohort |  | Age |  |  |  |  |  |  |  |  |  |  |  |  |  |  |  |
| --- | --- | --- | --- | --- | --- | --- | --- | --- | --- | --- | --- | --- | --- | --- | --- | --- | --- |
|  |  | Birth | 1 | 2 | 3 | 4 | 5 | 6 | 7 | 8 | 9 | 10 | 11 | 12 | 13 | 14 | 15 |
| 2006 | N | 87 | 868 | 390 | 388 | 345 | 283 | 365 | 352 | 303 | 312 | 319 | 429 | 243 | 296 | 329 | 453 |
|  | Cumulative N | 87 | 955 | 1,345 | 1,733 | 2,078 | 2,361 | 2,726 | 3,078 | 3,381 | 3,693 | 4,012 | 4,441 | 4,684 | 4,980 | 5,309 | 5,762 |
|  | Cumulative % | 0.1 | 1.1 | 1.6 | 2.1 | 2.5 | 2.8 | 3.2 | 3.6 | 4.0 | 4.3 | 4.7 | 5.2 | 5.5 | 5.8 | 6.2 | 6.7 |
| 2007 | N | 104 | 910 | 433 | 325 | 296 | 455 | 416 | 353 | 310 | 369 | 419 | 268 | 300 | 313 | 438 |  |
|  | Cumulative N | 104 | 1,014 | 1,447 | 1,772 | 2,068 | 2,523 | 2,939 | 3,292 | 3,602 | 3,971 | 4,390 | 4,658 | 4,958 | 5,271 | 5,709 |  |
|  | Cumulative % | 0.1 | 1.1 | 1.6 | 2.0 | 2.4 | 2.9 | 3.3 | 3.7 | 4.1 | 4.5 | 4.9 | 5.2 | 5.6 | 5.9 | 6.4 |  |
| 2008 | N | 104 | 931 | 388 | 321 | 450 | 426 | 345 | 330 | 399 | 441 | 265 | 342 | 333 | 369 |  |  |
|  | Cumulative N | 104 | 1,035 | 1,423 | 1,744 | 2,194 | 2,620 | 2,965 | 3,295 | 3,694 | 4,135 | 4,400 | 4,742 | 5,075 | 5,444 |  |  |
|  | Cumulative % | 0.1 | 1.2 | 1.6 | 2.0 | 2.5 | 2.9 | 3.3 | 3.7 | 4.1 | 4.6 | 4.9 | 5.2 | 5.6 | 6.0 |  |  |
| 2009 | N | 138 | 833 | 308 | 431 | 448 | 361 | 366 | 400 | 445 | 280 | 273 | 305 | 342 |  |  |  |
|  | Cumulative N | 138 | 971 | 1,279 | 1,710 | 2,158 | 2,519 | 2,885 | 3,285 | 3,730 | 4,010 | 4,283 | 4,588 | 4,930 |  |  |  |
|  | Cumulative % | 0.2 | 1.1 | 1.4 | 1.9 | 2.4 | 2.8 | 3.2 | 3.6 | 4.1 | 4.4 | 4.7 | 5.0 | 5.4 |  |  |  |
| 2010 | N | 124 | 641 | 464 | 442 | 402 | 364 | 384 | 414 | 274 | 298 | 326 | 332 |  |  |  |  |
|  | Cumulative N | 124 | 765 | 1,229 | 1,671 | 2,073 | 2,437 | 2,821 | 3,235 | 3,509 | 3,807 | 4,133 | 4,465 |  |  |  |  |
|  | Cumulative % | 0.1 | 0.8 | 1.4 | 1.8 | 2.3 | 2.7 | 3.1 | 3.5 | 3.8 | 4.1 | 4.5 | 4.8 |  |  |  |  |
| 2011 | N | 119 | 805 | 499 | 383 | 372 | 386 | 462 | 264 | 301 | 338 | 351 |  |  |  |  |  |
|  | Cumulative N | 119 | 924 | 1,423 | 1,806 | 2,178 | 2,564 | 3,026 | 3,290 | 3,591 | 3,929 | 4,280 |  |  |  |  |  |
|  | Cumulative % | 0.1 | 1.0 | 1.6 | 2.0 | 2.4 | 2.8 | 3.3 | 3.6 | 3.9 | 4.3 | 4.7 |  |  |  |  |  |
| 2012 | N | 252 | 882 | 434 | 354 | 425 | 432 | 308 | 318 | 317 | 377 |  |  |  |  |  |  |
|  | Cumulative N | 252 | 1,134 | 1,568 | 1,922 | 2,347 | 2,779 | 3,087 | 3,405 | 3,722 | 4,099 |  |  |  |  |  |  |
|  | Cumulative % | 0.3 | 1.2 | 1.7 | 2.1 | 2.5 | 3.0 | 3.3 | 3.7 | 4.0 | 4.4 |  |  |  |  |  |  |
| 2013 | N | 327 | 828 | 409 | 422 | 456 | 266 | 328 | 351 | 354 |  |  |  |  |  |  |  |
|  | Cumulative N | 327 | 1,155 | 1,564 | 1,986 | 2,442 | 2,708 | 3,036 | 3,387 | 3,741 |  |  |  |  |  |  |  |
|  | Cumulative % | 0.4 | 1.2 | 1.7 | 2.1 | 2.6 | 2.9 | 3.2 | 3.6 | 4.0 |  |  |  |  |  |  |  |
| 2014 | N | 308 | 744 | 484 | 422 | 258 | 304 | 396 | 346 |  |  |  |  |  |  |  |  |
|  | Cumulative N | 308 | 1,052 | 1,536 | 1,958 | 2,216 | 2,520 | 2,916 | 3,262 |  |  |  |  |  |  |  |  |
|  | Cumulative % | 0.3 | 1.2 | 1.7 | 2.2 | 2.4 | 2.7 | 3.2 | 3.5 |  |  |  |  |  |  |  |  |
| 2015 | N | 343 | 842 | 510 | 295 | 328 | 347 | 381 |  |  |  |  |  |  |  |  |  |
|  | Cumulative N | 343 | 1,185 | 1,695 | 1,990 | 2,318 | 2,665 | 3,046 |  |  |  |  |  |  |  |  |  |
|  | Cumulative % | 0.4 | 1.3 | 1.8 | 2.2 | 2.5 | 2.9 | 3.3 |  |  |  |  |  |  |  |  |  |
| 2016 | N | 423 | 866 | 339 | 343 | 365 | 400 |  |  |  |  |  |  |  |  |  |  |
|  | Cumulative N | 423 | 1,289 | 1,628 | 1,971 | 2,336 | 2,736 |  |  |  |  |  |  |  |  |  |  |
|  | Cumulative % | 0.4 | 1.4 | 1.7 | 2.1 | 2.4 | 2.9 |  |  |  |  |  |  |  |  |  |  |
| 2017 | N | 461 | 623 | 396 | 387 | 400 |  |  |  |  |  |  |  |  |  |  |  |
|  | Cumulative N | 461 | 1,084 | 1,480 | 1,867 | 2,267 |  |  |  |  |  |  |  |  |  |  |  |
|  | Cumulative % | 0.5 | 1.2 | 1.6 | 2.0 | 2.5 |  |  |  |  |  |  |  |  |  |  |  |
| 2018 | N | 285 | 690 | 455 | 430 |  |  |  |  |  |  |  |  |  |  |  |  |
|  | Cumulative N | 285 | 975 | 1,430 | 1,860 |  |  |  |  |  |  |  |  |  |  |  |  |
|  | Cumulative % | 0.3 | 1.1 | 1.6 | 2.1 |  |  |  |  |  |  |  |  |  |  |  |  |
| 2019 | N | 416 | 654 | 434 |  |  |  |  |  |  |  |  |  |  |  |  |  |
|  | Cumulative N | 416 | 1,070 | 1,504 |  |  |  |  |  |  |  |  |  |  |  |  |  |
|  | Cumulative % | 0.5 | 1.2 | 1.6 |  |  |  |  |  |  |  |  |  |  |  |  |  |
| 2020 | N | 486 | 660 |  |  |  |  |  |  |  |  |  |  |  |  |  |  |
|  | Cumulative N | 486 | 1,146 |  |  |  |  |  |  |  |  |  |  |  |  |  |  |
|  | Cumulative % | 0.5 | 1.3 |  |  |  |  |  |  |  |  |  |  |  |  |  |  |

**Table E6e.** Number, cumulative number and cumulative incidence of first-time OOHC placements by year of age for non-Aboriginal children born 2006-2020 (child protection service data only)

| Birth-year cohort |  | Age |  |  |  |  |  |  |  |  |  |  |  |  |  |  |  |
| --- | --- | --- | --- | --- | --- | --- | --- | --- | --- | --- | --- | --- | --- | --- | --- | --- | --- |
|  |  | Birth | 1 | 2 | 3 | 4 | 5 | 6 | 7 | 8 | 9 | 10 | 11 | 12 | 13 | 14 | 15 |
| 2006 | N | 0 | 333 | 194 | 158 | 137 | 91 | 90 | 80 | 75 | 88 | 80 | 64 | 31 | 25 | 26 | 25 |
|  | Cumulative N | 0 | 333 | 527 | 685 | 822 | 913 | 1,003 | 1,083 | 1,158 | 1,246 | 1,326 | 1,390 | 1,421 | 1,446 | 1,472 | 1,497 |
|  | Cumulative % | 0.0 | 0.4 | 0.6 | 0.8 | 1.0 | 1.1 | 1.2 | 1.3 | 1.4 | 1.5 | 1.6 | 1.6 | 1.7 | 1.7 | 1.7 | 1.7 |
| 2007 | N | 0 | 449 | 196 | 133 | 125 | 103 | 103 | 84 | 104 | 108 | 68 | 30 | 36 | 30 | 38 |  |
|  | Cumulative N | 0 | 449 | 645 | 778 | 903 | 1,006 | 1,109 | 1,193 | 1,297 | 1,405 | 1,473 | 1,503 | 1,539 | 1,569 | 1,607 |  |
|  | Cumulative % | 0.0 | 0.5 | 0.7 | 0.9 | 1.0 | 1.1 | 1.3 | 1.4 | 1.5 | 1.6 | 1.7 | 1.7 | 1.7 | 1.8 | 1.8 |  |
| 2008 | N | 0 | 446 | 163 | 127 | 121 | 111 | 100 | 94 | 88 | 67 | 49 | 40 | 34 | 39 |  |  |
|  | Cumulative N | 0 | 446 | 609 | 736 | 857 | 968 | 1,068 | 1,162 | 1,250 | 1,317 | 1,366 | 1,406 | 1,440 | 1,479 |  |  |
|  | Cumulative % | 0.0 | 0.5 | 0.7 | 0.8 | 1.0 | 1.1 | 1.2 | 1.3 | 1.4 | 1.5 | 1.5 | 1.6 | 1.6 | 1.6 |  |  |
| 2009 | N | 0 | 438 | 130 | 106 | 101 | 99 | 101 | 109 | 68 | 47 | 28 | 49 | 38 |  |  |  |
|  | Cumulative N | 0 | 438 | 568 | 674 | 775 | 874 | 975 | 1,084 | 1,152 | 1,199 | 1,227 | 1,276 | 1,314 |  |  |  |
|  | Cumulative % | 0.0 | 0.5 | 0.6 | 0.7 | 0.9 | 1.0 | 1.1 | 1.2 | 1.3 | 1.3 | 1.3 | 1.4 | 1.4 |  |  |  |
| 2010 | N | 0 | 375 | 102 | 108 | 110 | 119 | 125 | 64 | 51 | 49 | 32 | 34 |  |  |  |  |
|  | Cumulative N | 0 | 375 | 477 | 585 | 695 | 814 | 939 | 1,003 | 1,054 | 1,103 | 1,135 | 1,169 |  |  |  |  |
|  | Cumulative % | 0.0 | 0.4 | 0.5 | 0.6 | 0.8 | 0.9 | 1.0 | 1.1 | 1.1 | 1.2 | 1.2 | 1.3 |  |  |  |  |
| 2011 | N | 0 | 381 | 134 | 115 | 111 | 124 | 87 | 37 | 44 | 48 | 30 |  |  |  |  |  |
|  | Cumulative N | 0 | 381 | 515 | 630 | 741 | 865 | 952 | 989 | 1,033 | 1,081 | 1,111 |  |  |  |  |  |
|  | Cumulative % | 0.0 | 0.4 | 0.6 | 0.7 | 0.8 | 1.0 | 1.1 | 1.1 | 1.1 | 1.2 | 1.2 |  |  |  |  |  |
| 2012 | N | 0 | 365 | 145 | 155 | 146 | 83 | 59 | 44 | 41 | 49 |  |  |  |  |  |  |
|  | Cumulative N | 0 | 365 | 510 | 665 | 811 | 894 | 953 | 997 | 1,038 | 1,087 |  |  |  |  |  |  |
|  | Cumulative % | 0.0 | 0.4 | 0.6 | 0.7 | 0.9 | 1.0 | 1.0 | 1.1 | 1.1 | 1.2 |  |  |  |  |  |  |
| 2013 | N | 0 | 396 | 133 | 148 | 96 | 60 | 47 | 38 | 32 |  |  |  |  |  |  |  |
|  | Cumulative N | 0 | 396 | 529 | 677 | 773 | 833 | 880 | 918 | 950 |  |  |  |  |  |  |  |
|  | Cumulative % | 0.0 | 0.4 | 0.6 | 0.7 | 0.8 | 0.9 | 0.9 | 1.0 | 1.0 |  |  |  |  |  |  |  |
| 2014 | N | 0 | 390 | 167 | 94 | 58 | 48 | 53 | 43 |  |  |  |  |  |  |  |  |
|  | Cumulative N | 0 | 390 | 557 | 651 | 709 | 757 | 810 | 853 |  |  |  |  |  |  |  |  |
|  | Cumulative % | 0.0 | 0.4 | 0.6 | 0.7 | 0.8 | 0.8 | 0.9 | 0.9 |  |  |  |  |  |  |  |  |
| 2015 | N | 0 | 415 | 103 | 75 | 58 | 45 | 55 |  |  |  |  |  |  |  |  |  |
|  | Cumulative N | 0 | 415 | 518 | 593 | 651 | 696 | 751 |  |  |  |  |  |  |  |  |  |
|  | Cumulative % | 0.0 | 0.5 | 0.6 | 0.6 | 0.7 | 0.8 | 0.8 |  |  |  |  |  |  |  |  |  |
| 2016 | N | 0 | 365 | 88 | 57 | 57 | 60 |  |  |  |  |  |  |  |  |  |  |
|  | Cumulative N | 0 | 365 | 453 | 510 | 567 | 627 |  |  |  |  |  |  |  |  |  |  |
|  | Cumulative % | 0.0 | 0.4 | 0.5 | 0.5 | 0.6 | 0.7 |  |  |  |  |  |  |  |  |  |  |
| 2017 | N | 0 | 310 | 92 | 66 | 74 |  |  |  |  |  |  |  |  |  |  |  |
|  | Cumulative N | 0 | 310 | 402 | 468 | 542 |  |  |  |  |  |  |  |  |  |  |  |
|  | Cumulative % | 0.0 | 0.3 | 0.4 | 0.5 | 0.6 |  |  |  |  |  |  |  |  |  |  |  |
| 2018 | N | 0 | 288 | 98 | 82 |  |  |  |  |  |  |  |  |  |  |  |  |
|  | Cumulative N | 0 | 288 | 386 | 468 |  |  |  |  |  |  |  |  |  |  |  |  |
|  | Cumulative % | 0.0 | 0.3 | 0.4 | 0.5 |  |  |  |  |  |  |  |  |  |  |  |  |
| 2019 | N | 0 | 240 | 81 |  |  |  |  |  |  |  |  |  |  |  |  |  |
|  | Cumulative N | 0 | 240 | 321 |  |  |  |  |  |  |  |  |  |  |  |  |  |
|  | Cumulative % | 0.0 | 0.3 | 0.4 |  |  |  |  |  |  |  |  |  |  |  |  |  |
| 2020 | N | 0 | 232 |  |  |  |  |  |  |  |  |  |  |  |  |  |  |
|  | Cumulative N | 0 | 232 |  |  |  |  |  |  |  |  |  |  |  |  |  |  |
|  | Cumulative % | 0.0 | 0.3 |  |  |  |  |  |  |  |  |  |  |  |  |  |  |

## Footnotes

1 According to the Australian Institute of Aboriginal and Torres Strait Islander Studies, “Indigenous Data Sovereignty is the right of Indigenous peoples to govern the collection, ownership and application of data about Indigenous communities, peoples, lands, and resources. Its enactment mechanism, Indigenous Data Governance, is built around two central premises: the rights of Indigenous nations over data about them, regardless of where it is held and by whom; and the right to the data Indigenous peoples require to support nation rebuilding” (https://aiatsis.gov.au/publication/116530).

2 Noting that ‘restoration’ is the term used by New South Wales Department of Communities and Justice, often referred to as ‘reunification’ in the international literature.

